# Common and rare variant association analyses in Amyotrophic Lateral Sclerosis identify 15 risk loci with distinct genetic architectures and neuron-specific biology

**DOI:** 10.1101/2021.03.12.21253159

**Authors:** Wouter van Rheenen, Rick A.A. van der Spek, Mark K. Bakker, Joke J.F.A. van Vugt, Paul J. Hop, Ramona A.J. Zwamborn, Niek de Klein, Harm-Jan Westra, Olivier B. Bakker, Patrick Deelen, Gemma Shireby, Eilis Hannon, Matthieu Moisse, Denis Baird, Restuadi Restuadi, Egor Dolzhenko, Annelot M. Dekker, Klara Gawor, Henk-Jan Westeneng, Gijs H.P. Tazelaar, Kristel R. van Eijk, Maarten Kooyman, Ross P. Byrne, Mark Doherty, Mark Heverin, Ahmad Al Khleifat, Alfredo Iacoangeli, Aleksey Shatunov, Nicola Ticozzi, Johnathan Cooper-Knock, Bradley N. Smith, Marta Gromicho, Siddharthan Chandran, Suvankar Pal, Karen E. Morrison, Pamela J. Shaw, John Hardy, Richard W. Orrell, Michael Sendtner, Thomas Meyer, Nazli Başak, Anneke J. van der Kooi, Antonia Ratti, Isabella Fogh, Cinzia Gellera, Giuseppe Lauria Pinter, Stefania Corti, Cristina Cereda, Daisy Sproviero, Sandra D’Alfonso, Gianni Sorarù, Gabriele Siciliano, Massimiliano Filosto, Alessandro Padovani, Adriano Chiò, Andrea Calvo, Cristina Moglia, Maura Brunetti, Antonio Canosa, Maurizio Grassano, Ettore Beghi, Elisabetta Pupillo, Giancarlo Logroscino, Beatrice Nefussy, Alma Osmanovic, Angelica Nordin, Yossef Lerner, Michal Zabari, Marc Gotkine, Robert H. Baloh, Shaughn Bell, Patrick Vourc’h, Philippe Corcia, Philippe Couratier, Stéphanie Millecamps, Vincent Meininger, François Salachas, Jesus S. Mora Pardina, Abdelilah Assialioui, Ricardo Rojas-García, Patrick Dion, Jay P. Ross, Albert C. Ludolph, Jochen H. Weishaupt, David Brenner, Axel Freischmidt, Gilbert Bensimon, Alexis Brice, Alexandra Dürr, Christine A.M. Payan, Safa Saker-Delye, Nicholas Wood, Simon Topp, Rosa Rademakers, Lukas Tittmann, Wolfgang Lieb, Andre Franke, Stephan Ripke, Alice Braun, Julia Kraft, David C. Whiteman, Catherine M. Olsen, Andre G. Uitterlinden, Albert Hofman, Marcella Rietschel, Sven Cichon, Markus M. Nöthen, Philippe Amouyel, SLALOM Consortium, PARALS Consortium, SLAGEN Consortium, SLAP Consortium, Bryan Traynor, Adrew B. Singleton, Miguel Mitne Neto, Ruben J. Cauchi, Roel A. Ophoff, Martina Wiedau-Pazos, Catherine Lomen-Hoerth, Vivianna M. van Deerlin, Julian Grosskreutz, Annekathrin Rödiger, Nayana Gaur, Alexander Jörk, Tabea Barthel, Erik Theele, Benjamin Ilse, Beatrice Stubendorff, Otto W. Witte, Robert Steinbach, Christian A. Hübner, Caroline Graff, Lev Brylev, Vera Fominykh, Vera Demeshonok, Anastasia Ataulina, Boris Rogelj, Blaž Koritnik, Janez Zidar, Metka Ravnik-Glavač, Damjan Glavač, Zorica Stević, Vivian Drory, Monica Povedano, Ian P. Blair, Matthew C. Kiernan, Beben Benyamin, Robert D. Henderson, Sarah Furlong, Susan Mathers, Pamela A. McCombe, Merrilee Needham, Shyuan T. Ngo, Garth A. Nicholson, Roger Pamphlett, Dominic B. Rowe, Frederik J. Steyn, Kelly L. Williams, Karen Mather, Perminder S. Sachdev, Anjali K. Henders, Leanne Wallace, Mamede de Carvalho, Susana Pinto, Susanne Petri, Alma Osmanovic, Markus Weber, Guy A. Rouleau, Vincenzo Silani, Charles Curtis, Gerome Breen, Jonathan Glass, Robert H. Brown, John E. Landers, Christopher E. Shaw, Peter M. Andersen, Ewout J.N. Groen, Michael A. van Es, R. Jeroen Pasterkamp, Dongsheng Fan, Fleur C. Garton, Allan F. McRae, George Davey Smith, Tom R. Gaunt, Michael A. Eberle, Jonathan Mill, Russell L. McLaughlin, Orla Hardiman, Kevin P. Kenna, Naomi R. Wray, Ellen Tsai, Heiko Runz, Lude Franke, Ammar Al-Chalabi, Philip Van Damme, Leonard H. van den Berg, Jan H. Veldink

## Abstract

Amyotrophic lateral sclerosis (ALS) is a fatal neurodegenerative disease with a life-time risk of 1 in 350 people and an unmet need for disease-modifying therapies. We conducted a cross-ancestry GWAS in ALS including 29,612 ALS patients and 122,656 controls which identified 15 risk loci in ALS. When combined with 8,953 whole-genome sequenced individuals (6,538 ALS patients, 2,415 controls) and the largest cortex-derived eQTL dataset (MetaBrain), analyses revealed locus-specific genetic architectures in which we prioritized genes either through rare variants, repeat expansions or regulatory effects. ALS associated risk loci were shared with multiple traits within the neurodegenerative spectrum, but with distinct enrichment patterns across brain regions and cell-types. Across environmental and life-style risk factors obtained from literature, Mendelian randomization analyses indicated a causal role for high cholesterol levels. All ALS associated signals combined reveal a role for perturbations in vesicle mediated transport and autophagy, and provide evidence for cell-autonomous disease initiation in glutamatergic neurons.

## Introduction

Amyotrophic lateral sclerosis (ALS) is a fatal neurodegenerative disease affecting 1 in 350 individuals. Due to degeneration of both upper and lower motor neurons patients suffer from progressive paralysis, ultimately leading to respiratory failure within three to five years after disease onset.^1^ In ∼10% of ALS patients there is a clear family history for ALS suggesting a strong genetic predisposition and currently in more than half of these cases a pathogenic mutation can be found.^2^ On the other hand, apparently sporadic ALS is considered a complex trait where heritability is estimated at 40-50%.^3,4^ To date, partially overlapping GWASs have identified up to six genome-wide significant loci, explaining a small proportion of the genetic susceptibility to ALS.^5–10^ Some of these loci found in GWAS harbor rare variants with large effects also present in familial cases (e.g. *C9orf72* and *TBK1*).^11–13^ For other loci, the role of rare variants remains unknown.

While ALS is referred to as a motor neuron disease, cognitive and behavioral changes are observed in up to 50% of the patients, sometimes leading to frontotemporal dementia (FTD). The overlap with FTD is clearly illustrated by the pathogenic hexanucleotide repeat expansion in *C9orf72* which causes familial ALS and/or FTD^11,12^ and the genome-wide genetic correlation between ALS and FTD.^14^ Further expanding the ALS/FTD spectrum, a genetic correlation with progressive supranuclear palsy has been described.^15^ Shared pathogenic mechanisms between ALS and other neurodegenerative diseases, including common diseases such as Alzheimer’s and Parkinson’s disease, can further reveal ALS pathophysiology and inform new therapeutic strategies.

Here, we combine new and existing individual level-genotype data in the largest GWAS of ALS to date. We present a comprehensive screen for pathogenic rare variants and short tandem repeat (STR) expansions as well as regulatory effects observed in brain cortex-derived RNA-seq and methylation datasets to prioritize causal genes within ALS risk loci. Furthermore, we reveal similarities and differences between ALS and other neurodegenerative diseases as well as the biological processes in disease-relevant tissues and cell-types that affect ALS risk.

## Results

### Cross-ancestry meta-analysis reveals 15 risk loci for ALS

To generate the largest genome-wide association study in ALS to date, we merged individual level genotype data from 117 cohorts into 6 strata matched by genotyping platform. A total of 27,205 ALS patients and 110,881 control subjects of European ancestries passed quality control (**Online Methods, Supplementary Table 1-2**). Through meta-analysis of these six strata, we obtained association statistics for 10,461,755 variants down to an observed minor allele-frequency of 0.1% in the Haplotype Reference Consortium resource^16^. We observed moderate inflation of the test statistics (λ_GC_ = 1.12, λ_1000_ = 1.003) and linkage-disequilibrium score regression yielded an intercept of 1.029 (SE = 0.0073), indicating that the majority of inflation is due to the polygenic signal in ALS. The European ancestries analysis identified 12 loci reaching genome-wide significance (P < 5.0 × 10^−8^, **Supplementary Figure 1**). Of these, 8 were present in GWAS of ALS in Asian ancestries^8,10^ and all showed a consistent direction of effects (P_binom_ = 3.9 × 10^−3^). The genetic overlap between ALS risk in European and Asian ancestries resulted in a trans-ancestry genetic correlation of 0.57 (SE = 0.28) for genetic effect and 0.58 (SE = 0.30) for genetic impact, which were not statistically significant different from unity (P = 0.13 and 0.16, respectively). We therefore performed a cross-ancestry meta-analysis which revealed three additional loci, totaling 15 genome-wide significant risk loci for ALS risk (**Figure 1, Table 1, Supplementary Figures 2-16, Supplementary Tables 4-18**). Conditional and joint analysis did not identify secondary signals within these loci.

**Table 1.** Genome-wide significant loci. Details of the top associated SNPs within each genome-wide significant locus. Chr = chromosome, Basepair = position in reference genome GRCh37, A1 = effect allele, A2 = non-effect allele, Freq = frequency of the effect allele in European ancestries GWAS, SE = standard error of effect estimate.

| Chr | Basepair | ID | Prioritized gene | A1 | A2 | Freq | European ancestries |  | Asian ancestries |  | Cross-ancestry |  |
| --- | --- | --- | --- | --- | --- | --- | --- | --- | --- | --- | --- | --- |
|  |  |  |  |  |  |  | Effect (SE) | P | Effect (SE) | P | Effect (SE) | P |
| 9 | 27563868 | rs2453555 | *C9orf72* | A | G | 0.248 | 0.174 (0.013) | $1.0 \times 10^{-43}$ | 0.017 (0.066) | 0.80 | 0.168 (0.012) | $1.5 \times 10^{-41}$ |
| 19 | 17752689 | rs12608932 | *UNC13A* | C | A | 0.347 | 0.125 (0.012) | $8.8 \times 10^{-25}$ | 0.074 (0.038) | 0.053 | 0.120 (0.012) | $3.0 \times 10^{-25}$ |
| 21 | 33039603 | rs80265967 | *SOD1* | C | A | 0.006 | 1.078 (0.124) | $3.5 \times 10^{-18}$ | - | - | 1.078 (0.124) | $3.5 \times 10^{-18}$ |
| 14 | 31045596 | rs229195 | *SCFD1* | A | G | 0.337 | 0.091 (0.012) | $9.2 \times 10^{-15}$ | - | - | 0.091 (0.012) | $9.2 \times 10^{-15}$ |
| 3 | 39508968 | rs631312 | *MOBP/RPSA* | G | A | 0.291 | 0.079 (0.012) | $5.2 \times 10^{-11}$ | 0.084 (0.036) | 0.020 | 0.080 (0.011) | $3.3 \times 10^{-12}$ |
| 6 | 32672641 | rs9275477 | *HLA* | C | A | 0.096 | -0.143 (0.021) | $5.5 \times 10^{-12}$ | -0.110 (0.111) | 0.32 | -0.142 (0.02) | $3.5 \times 10^{-12}$ |
| 12 | 57975700 | rs113247976 | *KIF5A* | T | A | 0.016 | 0.332 (0.049) | $1.4 \times 10^{-11}$ | - | - | 0.332 (0.049) | $1.4 \times 10^{-11}$ |
| 21 | 45753117 | rs75087725 | *CFAP410* | A | C | 0.012 | 0.418 (0.063) | $2.7 \times 10^{-11}$ | - | - | 0.418 (0.063) | $2.7 \times 10^{-11}$ |
| 5 | 150410835 | rs10463311 | *GPX3/TNIP1* | C | T | 0.253 | 0.079 (0.013) | $3.5 \times 10^{-10}$ | 0.042 (0.036) | 0.24 | 0.075 (0.012) | $2.7 \times 10^{-10}$ |
| 20 | 48438761 | rs17785991 | *SLC9A8/SPATA2* | A | T | 0.353 | 0.074 (0.012) | $3.5 \times 10^{-10}$ | 0.045 (0.076) | 0.55 | 0.073 (0.012) | $3.2 \times 10^{-10}$ |
| 12 | 64877053 | rs4075094 | *TBK1* | A | T | 0.112 | -0.098 (0.018) | $1.7 \times 10^{-8}$ | -0.216 (0.090) | 0.017 | -0.103 (0.017) | $2.1 \times 10^{-9}$ |
| 5 | 172354731 | rs517339 | *ERGIC1* | C | T | 0.397 | -0.065 (0.011) | $8.5 \times 10^{-9}$ | -0.067 (0.074) | 0.37 | -0.065 (0.011) | $5.6 \times 10^{-9}$ |
| 4 | 170583157 | rs62333164 | *NEK1* | G | A | 0.335 | 0.063 (0.012) | $7.0 \times 10^{-8}$ | 0.203 (0.070) | $3.8 \times 10^{-3}$ | 0.067 (0.012) | $6.9 \times 10^{-9}$ |
| 13 | 46113984 | rs2985994 | *COG3* | C | T | 0.259 | 0.066 (0.013) | $1.9 \times 10^{-7}$ | 0.100 (0.041) | 0.014 | 0.069 (0.012) | $1.2 \times 10^{-8}$ |
| 7 | 157481780 | rs10280711 | *PTPRN2* | G | C | 0.124 | 0.076 (0.017) | $5.8 \times 10^{-6}$ | 0.132 (0.037) | $2.9 \times 10^{-4}$ | 0.086 (0.015) | $1.8 \times 10^{-8}$ |

**Figure 1.**
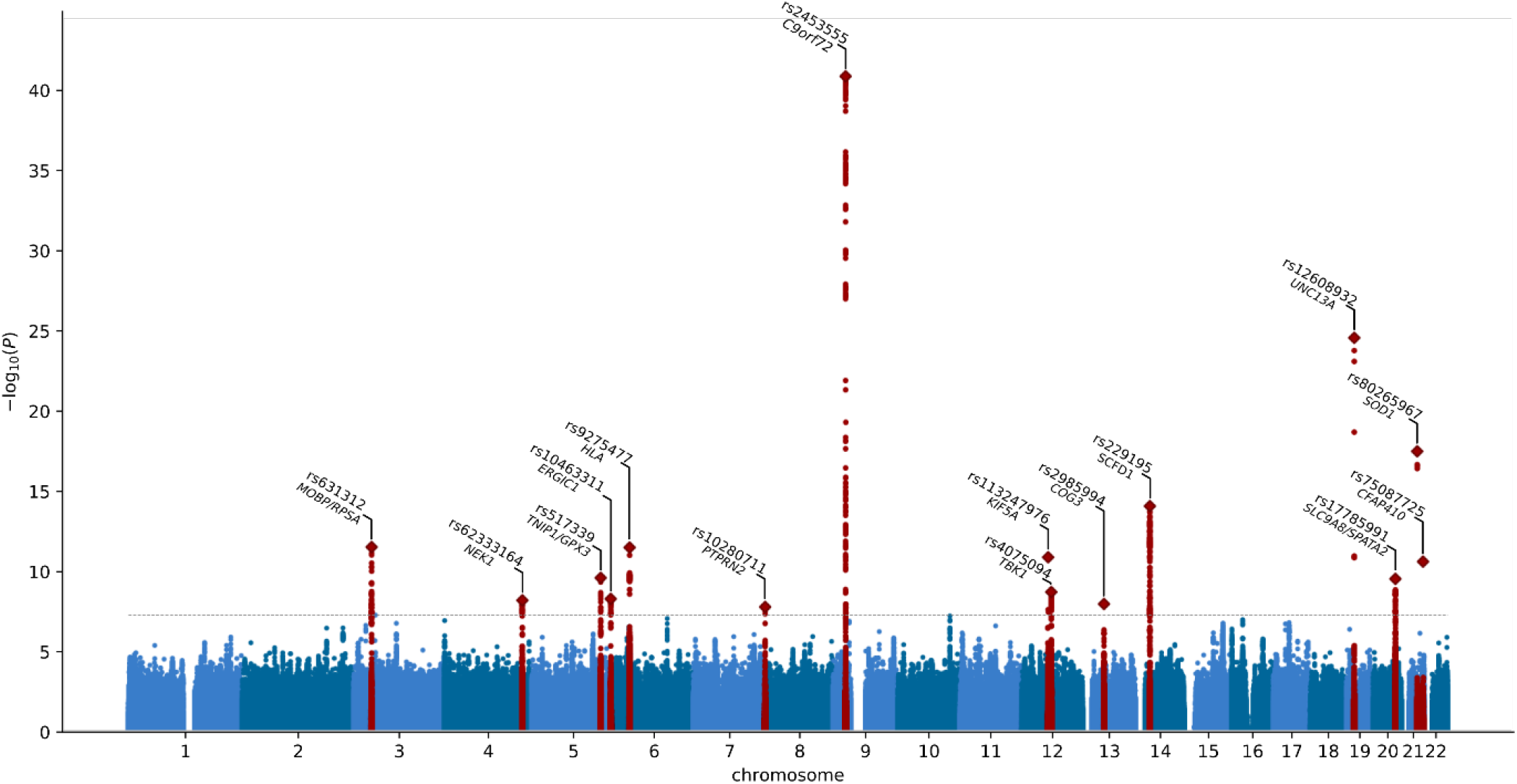
Manhattan plot of cross-ancestry meta-analysis. Horizontal dotted line reflects threshold for calling SNPs genome-wide significant (P = 5 × 10^−8^). Gene labels reflect those prioritized by gene prioritization analysis.

**Figure 2.**
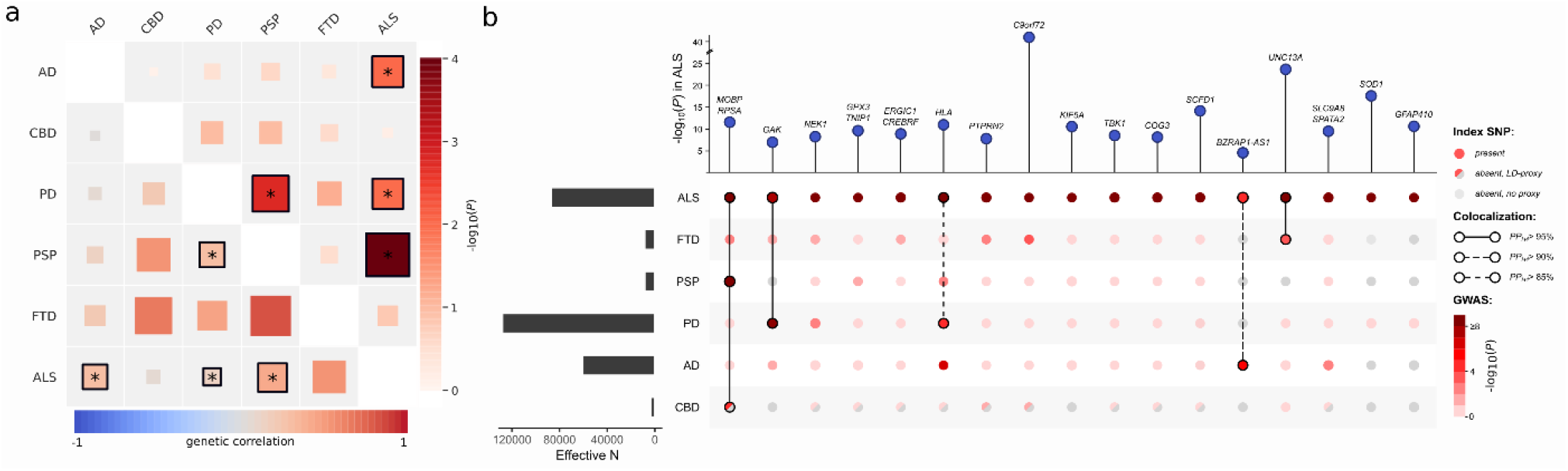
Shared genetic risk among ALS and neurodegenerative diseases. **(a)** Genetic correlation analysis. Genetic correlation was estimated with LD-score regression between each pair of neurodegenerative diseases being ALS, Alzheimer’s disease (AD), corticobasal degeneration (CBD), Parkinson’s disease (PD), progressive supranuclear palsy (PSP), and frontotemporal dementia (FTD). Lower left triangle shows correlation estimate and upper right triangle shows −log_10_ (P-value). Correlations marked with an asterisk were statistically significant P < 0.05. **(b)** SNP associations of ALS lead SNPs or LD-proxies in neurodegenerative diseases. Effective sample size is shown on the left. Posterior probabilities of the same causal SNP affecting two diseases were estimated through colocalization analysis and highlighted as connections.

Of these findings, 8 loci have been reported in previous genome-wide association studies (*C9orf72, UNC13A, SCFD1, MOBP/RPSA, KIF5A, CFAP410, GPX3/TNIP1*, and *TBK1*)^7–9^. The rs80265967 variant corresponds to the p.D90A variant in *SOD1* previously identified in a Finnish ALS cohort enriched for familial ALS^6^. Interestingly, we observed for the first time, a genome-wide significant common variant association signal within the *NEK1* locus, where *NEK1* was previously shown to harbor rare variants associated with ALS ^17^. The recently reported association at the *ACSL5-ZDHHC6* locus^10,18^ did not reach the threshold for genome-wide significance (rs58854276, P_EUR_= 5.4 × 10^−5^, P_ASN_= 4.9 × 10^−7^, P_comb_= 6.5 × 10^−8^, **Supplementary Figure 17, Supplementary Table 19**), despite that our analysis includes all data from the original discovery studies.

### Rare variant association analyses in ALS

To assess a general pattern of underlying architectures that link associated SNPs to causal genes we first tested for annotation specific enrichment using stratified linkage disequilibrium score regression (LDSC). This revealed that 5’ UTR regions as well as coding regions in the genome and those annotated as conserved were most enriched for ALS-associated SNPs (**Supplementary figure 18**). Subsequently we investigated how rare, coding variants contributed to ALS risk generating a whole-genome sequencing dataset of ALS patients (N = 6,538) and controls (N = 2,415). The exome-wide association analysis included transcript-level rare-variant burden testing for different models of allele-frequency thresholds and variant annotations (**Online methods**). This identified *NEK1* as the strongest associated gene (minimal P = 4.9 × 10^−8^ for disruptive and damaging variants at minor allele frequency [MAF] < 0.005), which was the only gene to pass the exome-wide significance thresholds (0.05/17,994 = 2.8 × 10^−6^ and 0.05/58,058 = 8.6 × 10^−7^ for number of genes and protein-coding transcripts, respectively, **Supplementary figures 19-32**). This association is independent from the previously reported increased rare variant burden in familial ALS patients^17^ that were not included in this study.

### Gene prioritization shows locus-specific underlying architectures

To assess whether rare variant associations could drive the common variant signals at the 15 genome-wide significant loci, we combined the common and rare variants analyses to prioritize genes within these loci. The SNP effects on gene expression were assessed through summary-based Mendelian Randomization (SMR) in blood (eQTLGen^19^) and a new brain cortex-derived expression quantitative trait locus (eQTL) dataset (MetaBrain^20^). Similarly, we analyzed methylation-QTL (mQTL) through SMR in blood and brain-derived mQTL datasets^21–23^. Finally, we leveraged the genome-wide signature of ALS associated gene features in a new gene prioritization method to calculate a polygenic priority score (PoPS)^24^. Through these multi-layered gene prioritization strategies we classified each locus into one of four classes of most likely underlying genetic architecture to prioritize the causal gene (**Supplementary figures 33-47**).

First, in three GWAS loci the strongest associated SNP was a low-frequency coding variant which was nominated as the causal variant. This is the case for rs80265967 (*SOD1*, p.D90A, **Supplementary Figure 46**) and rs113247976 (*KIF5A* p.P986L, **Supplementary Figure 40**) which are coding variants in known ALS risk genes. This is also the most likely causal mechanism for rs75087725 (*CFAP410*, formerly *C21orf2*, p.V58L, **Supplementary Figure 46**) as the GWAS variant is coding, no evidence for other mechanisms including repeat expansions, eQTL or mQTL effects is observed within this locus, and *CFAP410* itself is known to directly interact with *NEK1*, another ALS gene.^13,25^ These three loci illustrate the power of large-scale GWAS combined with modern imputation panels to directly identify low-frequency causal variants that confer disease risk.

Second, SNPs can tag a highly pathogenic repeat expansion, as is seen for rs2453555 (*C9orf72*) and the known GGGGCC hexanucleotide repeat in this locus. Conditional analysis revealed no residual signal after conditioning on the repeat expansion which is in LD with the top-SNP (r^2^ = 0.14, |D’ | = 0.99, MAF_SNP_= 0.25, MAF_STR_= 0.047). Besides the repeat expansion, both eQTL and mQTL analysis point to *C9orf72* (**Supplementary Figure 39**). The HEIDI outlier test, however, rejected the null hypothesis that gene expression or methylation mediated the causal effect of the associated SNP (P_HEIDI_eQTL_= 3.7 × 10^−23^ and P_HEIDI_mQTL_= 4.1 × 10^−7^). This is in line with the pathogenic repeat expansion as the causal variant in this locus as and that eQTL and mQTL effects do not mediate a causal effects. Across all other genome-wide significant loci, we found no similar repeat expansions that fully explain the SNP association signal.

Third, in two loci (rs62333164 in *NEK1* and rs4075094 in *TBK1*) common and rare variants converge to the same gene, which are known ALS risk genes^13,17^. For both loci, the rare variant burden association is conditionally independent from the top SNP which was included in the GWAS (**Supplementary figures 34 and 41**). Here, the eQTL and mQTL analyses indicated that the risk-increasing effects of the common variants are mediated through both eQTL and mQTL effects on *NEK1* and *TBK1*. Furthermore, a polymorphic STR downstream of *NEK1* was associated with increased ALS risk (motif = TTTA, threshold = 10 repeat units, expanded allele-frequency = 0.51, P = 5.2 × 10^−5^, FDR = 4.7 × 10^−4^, **Supplementary figure 48**). This polymorphic repeat is in LD with the top associated SNP within this locus (r^2^ = 0.24, |D’ | = 0.70). Within the whole-genome sequencing data, there was no statistically significant association for the top SNP to reliably determine its independent contribution to ALS risk.

Lastly, the fourth group contains remaining loci where there is no direct link to a causal gene through coding variants or repeat expansions. Here, we investigated regulatory effects of the associated SNPs on target genes acting as either eQTL or mQTL. Across all loci, single genes were prioritized by SMR using both mQTL and eQTL for rs2985994 (*COG3* **Supplementary Figure 42**), rs229243 (*SCFD1*, **Supplementary Figure 43**), and rs517339 (*ERGIC1*, **Supplementary Figure 36**). In other loci, both methods prioritized multiple genes, such as rs631312 (*MOBP* and *RPSA*, **Supplementary Figure 33**) and rs10463311 (*GPX3* and *TNIP1*, **Supplementary Figure 35**). Besides the prioritized genes, each of these loci harbor multiple genes that are not prioritized by any method and are therefore less likely to contribute to ALS risk.

### Locus-specific sharing of risk loci between ALS and neurodegenerative diseases

To investigate the pleiotropic properties of ALS-associated loci and shared genetic basis of neurodegeneration, we tested for shared effects among neurodegenerative diseases. We included GWAS from clinically-diagnosed Alzheimer’s disease (AD)^26^, Parkinson’s disease (PD)^27^, frontotemporal dementia (FTD)^28^, progressive supranuclear palsy (PSP)^15^ and corticobasal degeneration (CBD)^29^ to estimate genetic correlations across neurodegenerative diseases. Bivariate LDSC confirmed a statistically significant genetic correlation between ALS and PSP (r_g_= 0.44, SE = 0.11, P = 1.0 × 10^−4^) as previously reported, and also found a significant genetic correlation between ALS and AD (r_g_= 0.31, SE = 0.12, P = 9.6 × 10^−3^) as well as between ALS and PD (r_g_= 0.16, SE = 0.061, P = 0.011, **Figure 2a**). The point estimate for the genetic correlation between ALS and FTD was high (r_g_= 0.59, SE = 0.41, P = 0.15), but not statistically significant due to the limited size of the FTD GWAS. Thus, power to detect a genetic correlation between ALS and FTD using LDSC was limited (**Supplementary Figure 49**).

Patterns of sharing disease-associated genetic variants appeared to be locus specific (**Figure 2b, Supplementary Table 20**). To assess whether two traits shared a common signal indicating shared causal variants, we performed colocalization analyses for all loci meeting P < 5 × 10^−5^ in any of the GWAS on neurodegenerative diseases (N = 161 loci). This revealed a shared signal in the *MOBP/RPSA* between ALS, PSP and CBD, as well as a shared signal in the *UNC13A* locus between ALS and FTD (posterior probability: PP_H4_> 95%, **Supplementary Figure 50**). For the *HLA* locus, there was evidence for a shared causal variant between ALS and PD (PP_H4_= 88%) but no conclusive evidence for ALS and AD (PP_H4_= 51% for a shared causal variant and PP_H3_= 49% for independent signals in both traits).

Furthermore, the colocalization analyses identified two additional shared loci that were not genome-wide significant in the ALS GWAS: between ALS and PD at the *GAK* locus (rs34311866, PP_H4_= 99%) and between ALS and AD at the *BRZAP-AS1* locus (rs2632516, PP_H4_= 90%). Of note, the association at *BZRAP-AS1* was not genome-wide significant in the GWAS of clinically diagnosed AD (P = 3.7 × 10^−7^) either, but was identified in the larger AD-by-proxy GWAS^30^. For FTD subtypes, *C9orf72* showed a co-localization signal for a shared causal variant between ALS and the motor neuron disease subtype of FTD (mndFTD, PP_H4_= 93%, **Supplementary figure 50 and 51**).

### Enrichment of glutamatergic neurons indicate cell-autonomous processes in ALS susceptibility

To find tissues and cell-types which gene expression profiles are enriched for genes within ALS risk loci, we first combined gene-based association statistics calculated using MAGMA^31^ with gene expression patterns from GTEx (v8) in a gene-set enrichment analysis using FUMA.^32^ We observed a significant enrichment in genes expressed in brain tissues, specifically the cerebellum, basal ganglia (caudate nucleus, accumbens, and putamen), and cortex. Whereas this pattern roughly resembles the enrichments observed in PD, it is strikingly different from that observed in AD where blood, lung and spleen were mostly enriched (**Figure 3a**). We subsequently queried single-cell RNA sequencing datasets of human-derived brain samples to further specify brain-specific enriched cell-types using the cell-type analysis module in FUMA^33^. This showed significant enrichment for neurons but not microglia or astrocytes (**Figure 3b**). Further subtyping of these neurons illustrated that genes expressed in glutamatergic neurons were mostly enriched for genes within the ALS-associated risk loci. Again, this contrasted AD which showed specific enrichment of microglia. In single-cell RNA sequencing data obtained from brain tissues in mice, a similar pattern was observed showing neuron-specific enrichment in ALS and PD, but microglia in AD **(Supplementary Figure 52**). Together, this indicates that susceptibility to neurodegeneration in ALS is mainly driven by neuron-specific pathology and not by immune-related tissues and microglia.

**Figure 3.**
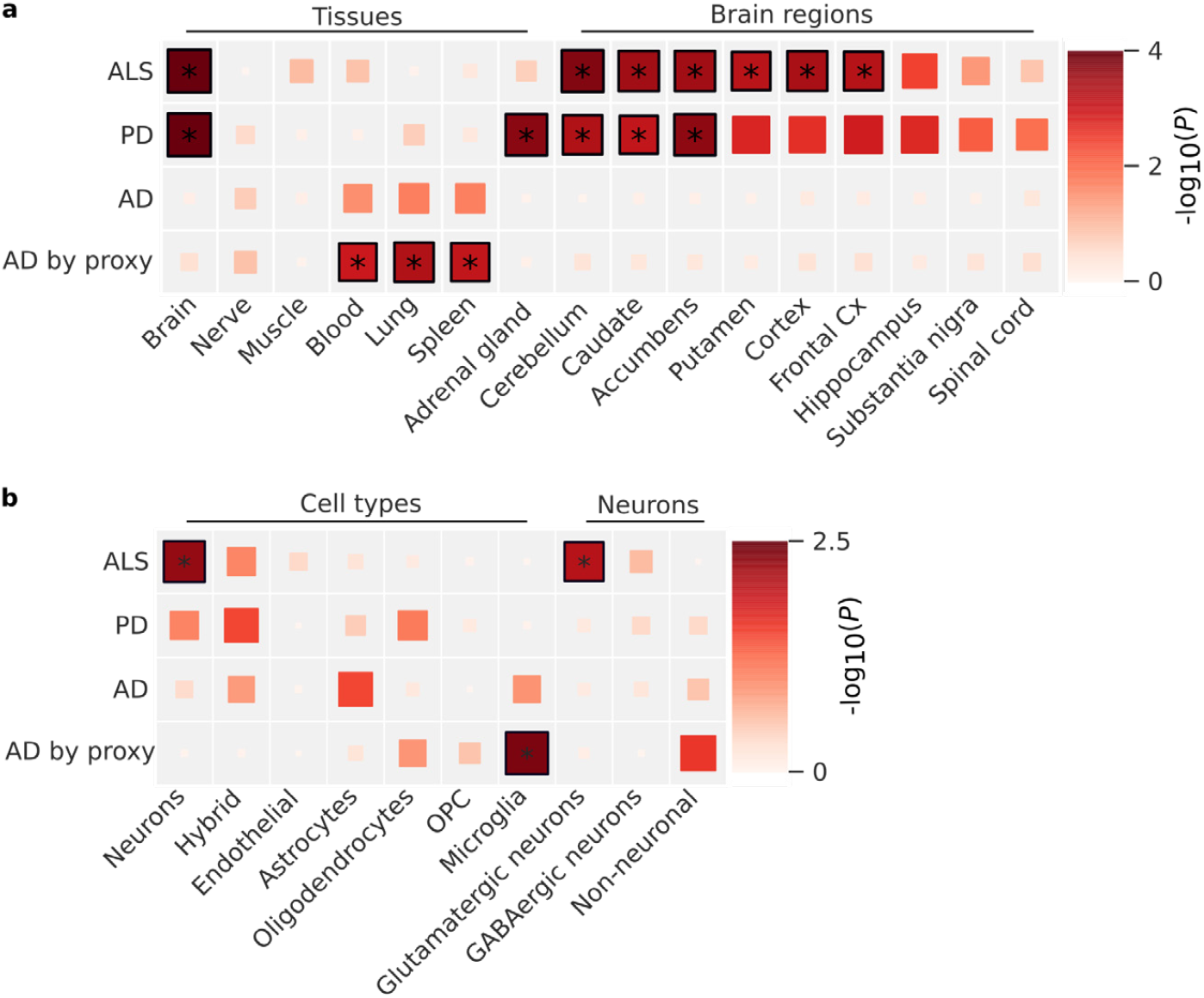
Tissue and cell-type enrichment analysis. **(a)** Enrichment of tissues and brain regions included in the GTEx v8 illustrates a brain-specific enrichment pattern in ALS, similar to Parkinson’s disease but contrasting Alzheimer’s disease. **(b)** Cell-type enrichment analyses indicate neuron-specific enrichment for glutamatergic neurons. No enrichment was found for microglia or other non-neuronal cell-types, contrasting the pattern observed in Alzheimer’s disease. Statistically significant enrichment after correction for multiple testing with a false discovery rate (FDR) < 0.05 are marked with an asterisk. ALS = amyotrophic lateral sclerosis, PD = Parkinson’s disease, AD = Alzheimer’s disease, Cx = cortex, OPC = Oligodendrocyte progenitor cells.

### Brain-specific co-expression networks improve ALS-relevant pathway detection

To assess which processes were mostly enriched in ALS, we performed enrichment analyses that combined gene-based association statistics with gene co-expression patterns obtained from either multi-tissue transcriptome datasets^34^ or RNA-seq data from brain cortex samples (MetaBrain^20^). To validate this approach, we first tested for enrichment of Human Phenotype Ontology (HPO) terms that are linked to well-established disease genes in the Online Mendelian Inheritance in Man (OMIM) and Orphanet catalogues. Using the multi-tissue co-expression matrix, we found no enriched HPO terms after Bonferroni correction for multiple testing. Using the brain-specific co-expression matrix however, we found a strong enrichment of HPO terms that are related to ALS or neurodegenerative diseases in general, including *Cerebral cortical atrophy* (P = 1.8 × 10^−8^), *Abnormal nervous system electrophysiology* (P = 4.1 × 10^−7^) and *Distal amyotrophy* (8.6 × 10^−7^, full-list in **Supplementary table 21**). In general, HPO terms in the neurological branch (*Abnormality of the nervous system*) showed an increase in enrichment statistics in ALS when using the brain-specific co-expression matrix compared to the multi-tissue dataset (**Supplementary Figure 53**), which illustrates the benefit of the brain-specific co-expression matrix for ALS-specific enrichment analyses. Subsequently, we tested for enriched biological processes using Reactome and Gene Ontology terms. Again, using the multi-tissue expression profiles, we found no Reactome annotations to be enriched. Leveraging the brain-specific co-expression networks we identified Vesicle Mediated Transport (“*Membrane Trafficking*” P = 4.2 × 10^−6^, “*Intra-golgi and retrograde Golgi-to-ER trafficking*” P = 1.4 × 10^−5^) and Autophagy (“*Macroautophagy*” P = 3.2 × 10^−5^) as enriched processes after Bonferroni correction for multiple testing (**Supplementary Table 22**). The enriched Gene Ontology terms all related to vesicle mediated transport or autophagy (**Supplementary Table 23** and **24**).

### Cholesterol levels are causally related to ALS

From previous observational case-control studies and our accompanying blood-based methylome-wide study^35^, numerous non-genetic risk factors have been implicated in ALS. Here we studied a selection of those putative risk factors through causal inference in a Mendelian randomization (MR) framework.^36^ We selected 22 risk factors for which robust genetic predictors were available including BMI, smoking, alcohol consumption, physical activity, cholesterol-related traits, cardiovascular diseases and inflammatory markers (**Supplementary Table 25**). These analyses provided the strongest evidence for cholesterol levels causally related to ALS risk (P_WeightedMedian_= 3.2 × 10^−4^, **Figure 4a**, full results in **Supplementary Table 26**). These results were robust to removal of outliers through Radial MR analysis^37^ and we observed no evidence for reverse causality (**Supplementary Table 27 and 28**). Importantly, ascertainment bias can lead to the selection of higher educated control subjects^38^, compared to ALS patients that are mostly ascertained through the clinic. In line with control subjects being longer educated, MR analyses indicate a negative effect for years of schooling on ALS risk (P_IVW_ = 2.0 × 10^−4^, **Figure 4b**). As a result, years of schooling can act as a confounder for the observed risk increasing effect of higher total cholesterol through ascertainment bias. To correct for this potential confounding, we applied multivariate MR analyses including both years of schooling and total cholesterol. The results for total cholesterol were robust in the multivariate analyses, suggesting a causal role for total cholesterol levels on ALS susceptibility (**Supplementary Table 29**).

**Figure 4.**
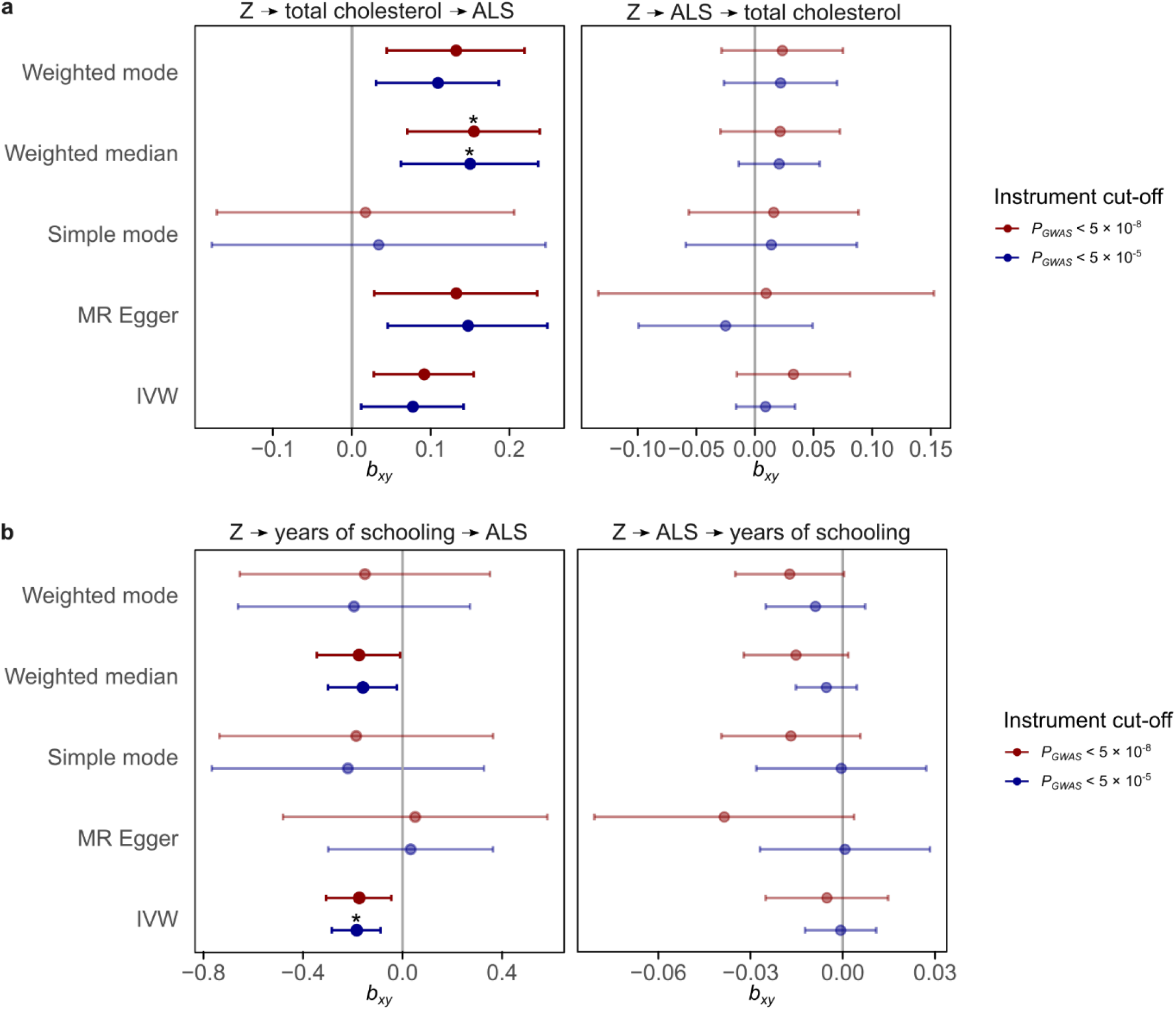
Causal inference of total cholesterol and years of schooling in ALS. **(a)** Mendelian randomization results for ALS and total cholesterol. Results for the five different Mendelian Randomization methods for two different P-value cut-offs for SNP instrument selection. All methods show a consistent positive effect for an increased risk of ALS with higher total cholesterol levels. There is no evidence for reverse causality. **(b)** Mendelian randomization results for ALS and years of schooling. Error-bars reflex 95% confidence intervals. Statistically significant effects that pass Bonferroni correction for multiple testing for all tested traits and MR methods are marked with an asterisk. Z = genetic instrument, MR = Mendelian Randomization, IVW = inverse-variance weighted, *b*_*xy*_ = _estimated_ causal effect for one standard deviation increase in genetically predicted exposure.

## Discussion

In summary, in the largest GWAS on ALS to date including 29,612 ALS patients and 122,656 control subjects, we have identified 15 risk loci contributing to ALS risk. Through in-depth analysis of these loci incorporating rare-variant burden analyses and repeat expansion screens in whole-genome sequencing data, blood and brain-specific eQTL and mQTL analysis we have prioritized genes in 14 of the loci. Across the spectrum of neurodegenerative diseases we identified a genetic correlation between ALS and AD, PD and PSP with locus-specific patterns of shared genetic risk across all neurodegenerative diseases. Colocalization analysis identified two additional loci, *GAK* and *BZRAP1-AS1*, with a high posterior probability of shared causal variants between ALS/PD, and ALS/AD respectively. We found glutamatergic neurons as the most enriched cell type in the brain and brain-specific co-expression network enrichment analyses indicated a role for vesicle-mediated transport and autophagy in ALS. Finally, causal inference of previously described risk factors provides evidence for high total cholesterol levels as a causal risk factor for ALS.

The cross-ancestry comparison illustrated similarities in the genetic risk factors for ALS in European and East Asian ancestries, providing an argument for cross-ancestry studies and to further expand ALS GWAS in non-European populations. Important to note is that 3 loci including those that harbor low-frequency variants (*KIF5A, SOD1*, and *CFAP410*) were not included in the East Asian GWAS due to their low minor allele frequency. Therefore, the shared genetic risk might not extend to rare genetic variation, for which population-specific frequencies have been observed even within Europe.

The multi-layered gene prioritization analyses highlighted four different classes of genome-wide significant loci in ALS. First, the sample size of this GWAS combined with accurate imputation of low-frequency variants directly identified rare coding variants that increase ALS risk. These include the known p.D90A mutation in *SOD1* (MAF = 0.006) as well as rare variants in *KIF5A* (MAF = 0.016) and *CFAP410* (MAF = 0.012) for which, after their identification through GWAS, experimental work confirms their direct role in ALS pathophysiology.^9,25,39^ Second, we confirmed that the pathogenic *C9orf72* repeat expansion is tagged by genome-wide significant GWAS SNPs, and that no residual signal is left by conditioning the SNP on the repeat expansion. Although more repeat expansions are known to affect ALS risk, we found no similar loci where the SNPs tag a highly pathogenic repeat expansion. This suggests that highly pathogenic repeat expansions on a stable haplotype are merely the exception rather than the rule in ALS. Third, common and rare variant association signals can converge on the same gene as is observed for *NEK1* and *TKB1*, consistent with observations for other traits and diseases^40–42^. We show that these signals are conditionally independent and that the common variants act on the same gene through regulatory effects as eQTL or mQTL. In the fourth class, we find evidence for regulatory effects of ALS associated SNPs that act as eQTL or mQTL. These locus-specific architectures illustrate the complexity of ALS associated GWAS loci where not one solution fits all, but instead warrants a multi-layered approach to prioritize genes.

In addition, we find locus-specific patterns of shared effects across neurodegenerative diseases. The *MOBP* locus has previously been identified in PSP and ALS and here we show that indeed both diseases, as well as CBD, are likely to share the same causal variant in this locus. The same is true for *UNC13A* and *C9orf72* with FTD and the motor neuron disease subtype of FTD, respectively. The colocalization analysis with PD identified a shared causal variant in the *GAK* locus, which was not found in the ALS GWAS alone. Furthermore the *BZRAP1-AS1* locus harbors SNPs associated with ALS and AD risk. Although this locus was not significant in either of the GWAS, larger GWAS including AD-by-proxy cases confirmed this as a risk locus for AD. This illustrates the power of cross-disorder analyses to leverage the shared genetic risk of neurodegenerative diseases.

We aimed to clarify the role of neuron-specific pathology in ALS susceptibility as opposed to non-cell autonomous pathology through detailed cell-type enrichment analyses. Previous experiments have illustrated multiple lines of evidence for non-cell autonomous pathology in microglia, astrocytes and oligodendrocytes which ultimately leads to neurodegeneration in ALS.^43–45^ These experiments have shown that non-cell autonomous processes, such as neuro-inflammation, mainly act as modifiers of disease in *SOD1* models of ALS^44,45^. Here, we show that genes within loci associated with ALS susceptibility are specifically expressed in (glutamatergic) neurons. This provides evidence for neuron-specific pathology as a driver of ALS susceptibility, which is in stark contrast to the signal of inflammation associated tissues and cell-types in Alzheimer’s disease^30^. It also shows that disease susceptibility and disease modification can be distinct processes, while both can be targets for potential new treatments in ALS.

The subsequent functional enrichment analyses identified membrane trafficking, Golgi to Endoplasmatic Reticulum (ER) trafficking and autophagy to be enriched for genes within ALS associated loci. These terms and their related Gene Ontology (GO) terms of biological processes are all related to autophagy and degradation of (misfolded) proteins. This corroborates the central hypothesis of impaired protein degradation leading to aberrant protein aggregation in neurons which is the pathological hallmark of ALS. Our results suggest that this is a central mechanism in ALS even in the absence of rare known mutations in genes directly involved in these biological processes such as *TARDBP, FUS, UBQLN2* and *OPTN*^*46*^.

Based on observational studies and MR analyses, conflicting evidence exists for lipid levels including cholesterol as a risk factor for ALS^47–49^. Potential selection bias, reverse causality and the subtype of cholesterol studied challenge the interpretation of these results. Here, we provided support for a causal relationship between high total cholesterol levels and ALS independent of educational attainment and ruling out reverse orientation of the MR effect. The total cholesterol effects were consistent across the different MR methods tested, indicating that this finding is robust to violation of the no horizontal pleiotropy assumption. This is in line with our accompanying study showing methylation changes associated with increased cholesterol levels in ALS^35^. We do not find a clear pattern for either LDL or HDL cholesterol subtypes in relation to ALS risk. Whereas cholesterol levels are closely related to cardiovascular risk, the association between cardiovascular risk and ALS risk remains controversial with conflicting reports.^3,47,50^. Interestingly, recent work has shown that lipid metabolism and autophagy are closely related which brings results of our pathway analyses and Mendelian randomization together.^51^ Both *in vitro* and *in vivo* experiments have shown that autophagy regulates lipid homeostasis through lipolysis and that impaired autophagy increases triglyceride and cholesterol levels. Conversely, high lipid levels were shown to impair autophagy.^51^ Further studies on the effect of high cholesterol levels and protein degradation through autophagy illustrate that high cholesterol levels decrease fusogenic ability of autophagic vesicles through decreased SNARE function^52,53^ and lead to increased protein aggregation due to impaired autophagy in mouse models for Alzheimer’s disease^54^. Therefore, the risk increasing effect of cholesterol on ALS might be mediated through impaired autophagy.

In conclusion, our genome-wide association study identifies 15 risk loci in ALS, and illustrates locus-specific interplay between common and rare genetic variation that helps prioritize genes for future follow-up studies. We show a causal role for cholesterol which can be linked to impaired autophagy as common denominators of neuron-specific pathology that drive ALS susceptibility and serve as potential targets for therapeutic strategies.

## Supporting information

Table with local IRBs

## Data Availability

All summary statistics will be made publicly available in centralized repositories upon publication after peer review.

## Methods

### GWAS

#### Data description

We obtained individual genotype level data for all individuals in the previously published GWAS in ALS in European ancestries^7,9^ and publicly available control datasets including 120,971 controls genotyped on Illumina platforms. Additionally 6,374 cases and 22,526 controls were genotyped on the IlluminaOmniExpress and Illumina GSA array. Details for each cohort are provided in **Supplementary Table 1**. For ALS cases, both cases with and without a family-history for ALS and/or dementia were included. Cases were not pre-screened for specific ALS related mutations. Given the late onset and relatively low life-time risk of ALS, controls were not screened for (subclinical) signs of ALS. A detailed description of the newly genotyped cases and controls is provided in the **Supplementary Information**. All participants gave written informed consent and the relevant local institutional review boards approved this study (**Supplementary Information**). Cases and controls formed cohorts when they were processed in the same lab and were genotyped in the same batch, resulting in 117 independent cohorts.

#### GWAS quality control and imputation

For each cohort, SNPs were first annotated according to dbSNP150 and mapped to the hg19 reference genome. All multi-allelic and palindromic (A/T or C/G) SNPs were excluded. Subsequently, basic quality control was first performed by cohort, excluding extremely low-quality SNPs and genotyped individuals as well as excluding extreme population outliers. Low quality SNPs and genotyped individuals were excluded using PLINK 1.9 (--geno 0.1 and --mind 0.1)^55^. Population structure was assessed by projecting HapMap3 principal components (PCs) using EIGENSOFT^56^ 6.1.4. Extreme outliers from the European ancestries population were removed (> 25 SD on PC1-4). Finally, cohorts were merged into strata based on genotyping platforms to preserve the maximum number of SNPs (**Supplementary Table 2**). Four out of 6 strata were formed by only a single platform. The remaining two strata included multiple platforms with 420,952 and 299,625 overlapping SNPs across platforms in these strata.

After excluding major SNP and sample outliers in cohort QC and merging cohorts into strata, stringent SNP QC was performed per stratum. The following filter criteria were applied: MAF > 0.01, SNP genotyping rate > 0.98, Deviation from Hardy-Weinberg disequilibrium in controls P > 1 × 10^−5^, and haplotype-biased missingness P > 1 × 10^−8^ (PLINK --maf 0.01, --geno 0.02, --hwe 1e-5 midp include-nonctrl, --test-mishap). Then, more stringent QC thresholds were applied to exclude individuals: individual missingness > 0.02, inbreeding coefficient |F| > 0.2, mismatches between genetic and reported gender, and missing phenotypes (PLINK --mind 0.02, --het, --check-sex). Subsequently, SNPs with a differential missingness (-- test-missing midp) P < 1 × 10^−4^ were excluded. Duplicate individuals were removed (PI_HAT > 0.8). Finally, outliers from the European ancestries reference population (projected on HapMap 3: > 10 SD from CEU on PC1-4 and projected on 1000 Genomes: > 4 SD from CEU on PC1-4) and outliers within the stratum itself (> 4SD from stratum mean on PC1-4) were removed (**Supplementary Figures 54-59**).

After removing outliers, principal components were recalculated for each stratum. To assess the result of quality control prior to imputation, genomic inflation factors per stratum were calculated using SAIGE^57^ to run a logistic mixed model regressing SNP genotype on ALS case-control status. SAIGE internally calculates an equivalent of a genetic relationship matrix to correct for relatedness and population structure. Additionally, PC1-20 and genotyping platform were included as covariates.

The number of individuals and SNPs passing quality control for each stratum prior to imputation is described in **Supplementary Table 2**.

#### Post Imputation quality control

Strata were then imputed using the HRC reference panel (r.1.1 2016) on the Michigan Imputation Server^16^. Data was phased using Eagle 2.3. After imputation, one individual of each pair of related samples across strata (PI_HAT > 0.125) was removed whereas related pairs within a stratum were retained since the genetic relationship matrix corrects for relatedness. Post-imputation variant-level quality control included removing all monomorphic SNPs and multi-allelic SNPs from each stratum. SNPs with MAF < 0.1% in the HRC imputation panel were excluded. Subsequently, INFO scores were calculated for each stratum based on dosage information using SNPTEST^58^ v2.5.4-beta3. Within each stratum, SNPs with an INFO-score < 0.6 and those deviating from Hardy-Weinberg equilibrium at P < 1 × 10^−5^ in control subjects were removed. Effective sample size was calculated for each stratum:

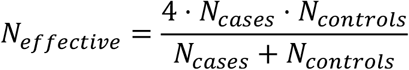

The difference in sample size and number of SNPs for each stratum prior to imputation, resulted in a different set of SNPs passing post-imputation quality control for each stratum. Therefore, only SNPs that were successfully imputed in an effective sample meeting > 50% of the maximum effective sample size were included.

The number of individuals and SNPs passing quality control for each stratum after imputation is described in **Supplementary Table 2**.

#### Association testing and meta-analysis

After quality control, a null logistic mixed model was fitted using SAIGE^57^ 0.29.1 for each stratum with PC1-20 as covariates. The model was fit on a set of high-quality (INFO >0.95), pruned with PLINK 1.9, (--indep-pairwise 50 25 0.1) SNPs in a leave-one-chromosome-out scheme. Subsequently, a SNP-wise logistic mixed model including the saddle point approximation test was performed using genotype dosages with SAIGE. Association statistics for all strata were combined in an inverse variance-weighted fixed effects meta-analysis using METAL^59^.

Genomic inflation factors were calculated per stratum and for the full meta-analysis. To assess any residual confounding due to population stratification and artificial structure in the data we calculated the LD Score regression (LDSC)^60^ intercept using SNP LD-scores calculated in the HapMap3 CEU population.

#### Cross-ancestry analyses

GWAS summary statistics from two Asian ancestry studies were obtained^8,10^. These summary statistics were meta-analyzed with all European ancestry in strata as described above. To assess genetic correlation for ALS in the European and Asian ancestries, we used Popcorn^61^ version 0.9.9. We used population specific LD scores for genetic impact and genetic effect provided with the Popcorn software. The regression model (--use_regression) was used to estimate genetic correlation. We calculated both the correlation of genetic effects (correlation of allelic effect sizes) and genetic impact (correlation of allelic effect size adjusted for difference in allele frequencies).

#### Conditional SNP analysis

Conditional and joint SNP analysis (COJO, GCTA v1.91.1b)^62,63^ was performed to identify potential secondary GWAS signals within a single locus. SNPs with association P ≤ 5 × 10^−8^ were considered. European ancestry controls from the health and retirement study (HRS, cohort 65, **Supplementary Table 1**), included in stratum 4 of this study, were used as LD reference panel.

### Gene prioritization

#### Whole-genome sequencing

##### Sample selection, sequencing and data preparation

ALS cases and controls from Project MinE^64^ were recruited for whole genome sequencing. The participating cohorts are described in the **Supplementary Note**. A full description of Project MinE, the sequencing and quality control pipeline were described previously^65^. In summary, the first batch of 2,250 cases and control samples were sequenced on the Illumina HiSeq 2000 platform. All remaining 7,350 cases and controls were sequenced on the Illumina HiSeq X platform. All samples were sequenced to ∼35X coverage with 100bp reads and ∼25X coverage with 150bp reads for the HiSeq 2000 and HiSeq X respectively. Both sequencing sets used PCR-free library preparation. Samples were also genotyped on the Illumina 2.5M array. Sequencing data was then aligned to GRCh37 using the iSAAC Aligner, and variants called using the iSAAC variant caller; both the aligner and caller are standard to Illumina’s aligning and calling pipeline.

##### Quality control

For variant-level quality control, we set sites with a genotype quality (GQ) < 10 to missing and SNVs and indels with quality (QUAL) scores < 20 and < 30, respectively, were removed. We subsequently performed sample-level quality control. An overview of the number of samples that have been excluded at each of the following QC steps, stratified by country of origin, is included in **Supplementary Table 3**.

We estimated kinship coefficients (i.e., relatedness) using the KING method, as implemented in the SNPRelate package in R. In some instances, cohorts were intentionally enriched for related samples. We identified all pairs of related individuals (kinship > 0.0625).

We calculated the transition-transversion ratio in each sample using SnpSift 4.3p. In WGS data, the expected transition-transversion ratio is ∼2.0. Samples with a Ti/Tv ratio ± 6 SD from the full distribution of samples were removed.

Per sample, we calculated the total number of SNVs and total number of singletons. We removed samples with a total number of SNVs or Singletons > 6 SD from the mean. The transition in sequencing platforms from HiSeq 2000 to HiSeq X (which occurred in parallel with a change in the calling pipeline, to improve indel detection) caused an increase in observed indels per sample. Samples were thus filtered by platform (HiSeq 2000 or HiSeq X) and removed samples with number of indels ± 6 SD from the mean of their respective group.

We calculated average sample depth and again observed noticeable differences between those samples sequenced on the HiSeq 2000 and the HiSeq X, where average depth of coverage was somewhat higher (35X, on average) for samples sequenced on HiSeq 2000 compared to the samples sequenced on the HiSeqX (25X, on average). We removed no samples at this step.

Using the genetically inferred sex based on the number of X and Y chromosome, we tested to see if the inferred genetic sex was concordant with the sex as annotated in the available phenotype information. We excluded samples with mismatching information and samples for which phenotypic information is missing at this time.

We performed the remaining sample QC on high-quality variants: We removed all multi-allelic SNVs, Plink 1.9 (--geno), variants with a missingness > 2% were excluded. We calculated Hardy-Weinberg equilibrium (HWE) in controls only, PLINK 1.9 (--hwe midp), and removed all variants with HWE P < 1 × 10^−5^. We calculated differential missingness, PLINK 1.9 (--test-mishap) between cases and controls and removed variants with P < 1 × 10^−8^. Samples with a missingness > 2%, in SNV and indels, were excluded. Final steps of sample QC was performed on a set of variants with a MAF > 10%, SNP missingness < 0.1%, variants residing outside four complex regions (the major histocompatibility complex (MHC) on chromosome 6; the lactase locus (LCT), on chromosome 2; and inversions on chromosomes 8 and 17); and we excluded the A/T and C/G variants. We used the SNVs to calculate observed and expected autosomal homozygous genotype counts for each sample PLINK 1.9 (--het); samples with |F| > 0.1 were excluded. We excluded duplicate samples; PLINK 1.9 (--genome) with a PIHAT > 0.8, keeping the maximum number of non-duplicated individuals.

Principal component analysis (PCA) implemented in EIGENSOFT was used to visualize potential structure in the data, induced by population stratification or other variables. Projections onto HapMap3 and the 1KG phase3 v5 populations indicated that the samples were primarily of European ancestry, though some were of African or East Asian ancestries, while other samples appeared to be admixed. Outliers from the European population (HapMap3: > 10 SD on PC1-4, 1KG: > 4 SD on PC1-4).

All samples were sent in batches to Illumina for sequencing. To prevent spurious association due to batch specific artifacts, we regressed all variants on a dummy coded variable indicating batch using PLINK 1.9 (--logistic). All variants with an association P < 1 × 10^−10^ in at least 1 batch were excluded.

##### Genic burden association analyses

To aggregate rare variants in a genic burden test framework we used a variety of variant filters to allow for different genetic architectures of ALS associated variants per gene as we and others have used previously^65,66^. In summary, variants were annotated according to allele-frequency threshold (MAF < 0.01 or MAF < 0.005) and predicted variant impact (“missense”, “damaging”, “disruptive”). “Disruptive” variants were those variants classified as frame-shift, splice-site, exon loss, stop gained, start loss and transcription ablation. “Damaging” variants were missense variants predicted to be damaging by seven prediction algorithms (SIFT^67^, Polyphen-2^68^, LRT^69^, MutationTaster2^70^, Mutations Assessor^71^, and PROVEAN^72^). “Missense” variants are those missense variants that did not meet the “damaging” criteria. All combinations of allele frequency threshold and variant annotations were used to test the genic burden on a transcript level in a Firth logistic regression framework where burden was defined as the number of variants per individual. Sex and the first 20 principal components were included as covariates. All ENSEMBL protein coding transcripts for which at least five individuals had a non-zero burden were included in the analysis.

##### Conditional genic burden analysis

We selected for each gene the protein coding transcripts that were strongest associated with ALS across all different combinations of MAF and variant impact thresholds that exhibited the strongest association with ALS. For these transcripts and variants, we applied Firth logistic regression on individuals overlapping the GWAS and WGS dataset (5,158 cases and 2,167 controls). To assess whether the rare variant burden association and the signal from GWAS were conditionally independent we subsequently included the genotype of the top-associated SNP within that locus as covariate.

#### Short tandem repeat screen

For all individuals that were sequenced on the HiSeqX dataset (5,392 cases, 1,795 controls) we screened all loci harboring SNPs associated with ALS meeting genome-wide significance for expansions of known and new short tandem repeats (STRs) using ExpansionHunter^73^ and ExpansionHunter Denovo^74^.

First we used ExpansionHunter (v4.0) to screen for expansions of known STRs located within 1 MB of the top ALS-associated SNP. For this we used the STR catalogue of the ExpansionHunter software which is based on STRs identified from indels in 18 high quality genomes and the gangSTR STR catalogue based on STR annotations in the reference genome^75^. From these catalogues, we excluded all homopolymers. Repeat length was subsequently regressed on case-control status using Firth logistic regression including the first 20 principal components as covariates, recoding the STR size to a biallelic variant using a sliding window over all observed repeat lengths. To correct for multiple testing across all possible thresholds, we applied Benjamini Hochberg correction per STR.

To screen for extremely long STR expansions (similar to the *C9orf72* repeat expansion) at loci that not included in the predefined STR catalogues, we applied ExpansionHunter-Denovo^74^. This method aims to only find STR expansions that exceed the sequencing read-length (> 150 bp) by identifying reads (mapped, mismapped and unmapped) that contain STR motifs, using their mate pairs for *de novo* mapping to the reference genome.

For all STRs we calculated linkage disequilibrium statistics (*r*^*2*^ and |D’ |) between recoded repeat genotypes at the optimal threshold and the top associated GWAS SNP. Subsequently, we conditioned the SNP association on the repeat genotype in a Firth logistic regression.

### Summary-based Mendelian randomization

We used multi-SNP SMR^76,77^ to infer the effect of gene expression variation on ALS using eQTLs (the association of a SNP with expression of a gene) on ALS risk. MetaBrain is a harmonized set of 8,727 RNA-seq samples from 7 regions of the central nervous system from 15 datasets, and we selected eQTLs derived from the cortex region of the brain in samples of European ancestry (MetaBrain Cortex-EUR eQTLs) as our instrument variable^20^. The European-only ALS summary statistics were used as the outcome. To supplement this analysis, we also used eQTLs in blood from the eQTLGen consortium, as this is the largest eQTL resource available. European-ancestry samples in the Health and Retirement study (HRS, cohort 65 of this GWAS) were used as LD reference panel. SNP with MAF ≥ 1% in HRS were included. Further SMR settings were left as default, meaning probes with at least one eQTL with P ≤ 5 × 10^−8^ were included.

We subsequently performed SMR using DNA methylation QTL (mQTL) data and European-only ALS summary statistics. Human prefrontal cortex and whole blood DNA mQTLs were generated as part of ongoing analyses by the Complex Disease Epigenomics Group at the University of Exeter (www.epigenomicslab.com) using the Illumina EPIC HumanMethylation array that quantifies DNAm at >850,000 sites across the genome^21^. The prefrontal cortex mQTL dataset was generated using DNA methylation and SNP data from 522 individuals from the Brains for Dementia Research cohort^22^ and included 4,623,966 cis mQTLs (distance between QTL SNP and DNAm site ≤ 500 kb) between 1,744,102 SNPs and 43,337 DNA methylation sites. The whole blood mQTL dataset was generated using DNAm and SNP data from 2,082 individuals^78^ and included 30,432,023 cis mQTLs between 4,030,902 SNPs and 167,854 DNA methylation sites. mQTLs reaching the significance threshold P ≤ 1 × 10^−10^ were taken forward for SMR analysis as described by Hannon and colleagues^78^. To map CpG sites to their putative target genes we used the expression quantitative trait methylation (eQTM) results from a paired methylation and gene expression (RNA-seq) study in blood^79^. For CpG sites where no eQTM were present in this dataset, we used positional mapping based on the basal regulatory domains and extended regulatory domains as defined in the Genomic Regions Enrichment of Annotations Tool (GREAT)^80^ which is applied in the ‘cpg_to_gene’ function in the CpGtools toolkit^81^.

### Polygenic Priority Score (PoPS)

We used the polygenic priority score (PoPS^24^ v0.1) to rank genes according to the gene features that were enriched in ALS. For this we applied MAGMA in the European ancestries GWAS since it depends on an LD reference panel (1000 Genomes Project, EUR population) to obtain gene-wise association statistics. We used the default 57,543 gene features that were based on expression data, protein-protein interaction networks and pathway membership. Genes were ranked based on the Polygenic Priority Score.

### Cross-trait analyses in neurodegenerative diseases

#### Datasets and data preparation

GWAS summary statistics for clinically-diagnosed Alzheimer’s disease (AD)^26^, Parkinson’s disease (PD)^27^, frontotemporal dementia (FTD)^28^, corticobasal degeneration (CBD)^29^, and progressive supranuclear palsy (PSP)^15^ in European ancestry individuals were obtained. For Alzheimer’s disease we used the clinically diagnosis as case definition to avoid spurious genetic correlations that could have been introduced through the by-proxy design^30^ where by-proxy cases are defined as having a parent with Alzheimer’s disease. Although this is a powerful design for gene discovery and the genetic correlation with clinically diagnosed Alzheimer’s disease is high^82^, mislabeling by-proxy cases when parents suffer from other types of dementia (e.g. Lewy-body dementia, Parkinson’s dementia, FTD, or vascular dementia) can lead to spurious genetic correlations with ALS and other neurodegenerative diseases. For FTD, we primarily used the results of the cross-subtype meta-analysis which includes behavioral variant FTD (bvFTD), semantic dementia (sdFTD), progressive non-fluent aphasia (pnfaFTD) and motor neuron disease FTD (mndFTD). For CBD, allele coding were missing and effect alleles were inferred by matching allele frequencies to those observed in the Haplotype Reference Consortium. SNPs with minor allele frequency > 0.4 were excluded. Since downstream methods rely on LD-scores or population-specific LD patterns, the European ancestry summary statistics from the present study were used for ALS. For sample size parameters, effective sample size was calculated as described previously.

#### Genetic correlation

We first assessed residual confounding through estimating the LD Score regression^60^ intercept using LDSC (v.1.0.0): ALS = 1.03 (SE 0.0073), AD = 1.03 (SE 0.013), PD = 0.98 (SE 0.0065), PSP = 1.05 (SE 0.0076), CBD = 0.98 (SE 0.0073), FTD = 1.00 (SE 0.0071), showing limited inflation of test statistics due to confounding across these studies. Genome-wide genetic correlation between neurodegenerative traits was calculated using LDSC (v1.0.0). Pre-computed LD-scores of European individuals in the 1000 Genomes project for high-quality HapMap3 SNPs were used (eur_w_ld_chr). A free intercept was modelled to allow for potential sample overlap.

#### Colocalization

For each locus (top-SNP +/-100KB) harboring SNPs with an association with any of the neurodegenerative diseases at P < 1 × 10^−5^ we performed colocalization analysis using the ‘coloc’ package in R.^83^ We set the prior probabilities to π _1_ = 1 × 10^−4^, π _2_ = 1 × 10^−4^, π_12_ = 1 × 10^−5^ for a causal variant in trait 1, trait 2 and a shared causal variant between trait 1 and 2 respectively. Using the same parameters, we performed colocalization analysis for ALS and each of the FTD subtypes (bvFTD, sdFTD, pnfaFTD, mndFTD).

### Enrichment analyses

#### LD-score regression annotation-specific enrichment analysis

We used LDSC (v1.0.0) to calculate SNP-based heritability, the LDSC intercept and SNP-based heritability enrichment for partitions of the genome. In all LDSC analyses, summary statistics excluding the HLA region of only European ancestry samples were included. LD scores and partitioned LD scores provided by LDSC were used for genome-wide and genic region-based heritability analyses. The option --overlap-annot was used in the partitioned heritability analysis to allow for overlapping SNP between MAF bins. SNPs with a MAF > 5% were included.

#### Tissue and cell-type enrichment analysis

Tissue and cell-type enrichment analyses were performed using the GWAS summary statistics of the European ancestries meta-analysis and FUMA^32^ software v1.3.6a. FUMA performs a genic aggregation analysis of GWAS association signals to calculate gene-wise association signals using MAGMA v1.6 and subsequent tests whether tissues and cell-types are enriched for expression of these genes. For tissue enrichment analysis we used the GTEx v8 reference set. For cell-type enrichment analyses^33^ we used human-derived single-cell RNA sequencing data on major brain cell-types (GSE67835 without fetal samples^84^), the Allen Brain Atlas Cell-type^85^ for the human-derived major neuronal subtypes and the DropViz^86^ dataset for mouse-derived brain cell-types across all brain regions.

#### Pathway enrichment analysis

We used the Downstreamer software^20^ to identify enriched biological pathways and processes. First, gene-based association statistics are obtained through the PASCAL method^87^ which aggregates SNP association statistics including SNPs up to 10kb up- and downstream of a gene, accounting for linkage disequilibrium using the non-Finish European individuals from the 1000 Genomes Project phase 3 (ref. ^88^) as a reference. In the Downstreamer method, putative core genes are defined as those that are coexpressed with disease-associated genes and can therefore be implicated in disease. Co-expression networks are based on either a large, multi-tissue transcriptome dataset including 56,435 genes and 31,499 individuals, or brain-specific RNA-sequencing data obtained in the MetaBrain resource. The gene-based association statistics, co-expression matrix and gene Z-scores per pathway or HPO term are then combined in a generalized least squares regression model to obtain enrichment statistics.^20^ Enrichment analyses were performed for Reactome, Gene Ontology and Human Phenotype Ontology (HPO) terms using the multi-tissue or brain-specific transcriptome datasets to calculate the co-expression matrix.

The distribution of enrichment Z-score statistics were compared between the analyses using the multi-tissue or the brain-specific co-expression matrices. Using the ‘pyhpo’ module in Python, all HPO terms were assigned to their parent term(s) in the “*Phenotypic abnormality*” (HP:0000118) branch which includes phenotypic abnormalities grouped per organ system.

### Mendelian Randomization

Causal inference through MR analysis was performed for 22 exposures for which large-scale GWAS are available and for which there is prior evidence for an association with ALS. These include 7 behavioral related traits: body mass index (anthropometric)^89^, years of schooling (educational attainment)^90^, alcoholic drinks per week, age of smoking initiation and cigarettes per day from Liu et al.^91^, days per week moderate physical activity and days per week vigorous activity from UK Biobank^92^; 4 blood pressure traits: coronary artery disease^93^, stroke^94^, diastolic blood pressure and systolic blood pressure^95^; 7 immune system traits from Vuckovic et al.^96^ (basophil, eosinophil, lymphocyte, monocyte, neutrophil and while blood cells) and C-reactive protein^97^; and 4 lipid traits from Willer et al.^98^ (HDL cholesterol, LDL cholesterol, total cholesterol and triglycerides). A full description of the included studies is provided in **Supplementary Table 25**. From these GWASs, SNPs to serve as instruments for MR analyses were selected at two different p-value cut-offs (P < 5 × 10^−8^ and P < 5 × 10^−5^) and then LD clumped to obtain independent SNPs. SNP effect estimates on ALS risk were obtained from the European ancestries only GWAS and if needed an LD-proxy was selected (r^2^ > 0.8).

After harmonizing effect-alleles and excluding palindromic SNPs, we performed a series of quality control steps to avoid biased estimates of causal effects, checking for each exposure the (i) instrument coverage (> 85% overlapping SNPs, **Supplementary Table 30**), (ii) instrument strength (F-statistic^36,99,100^ > 10, **Supplementary Table 31**), (iii) distribution and significance of the Wald ratios (visual inspection of volcano plots, **Supplementary Table 32**) and (iv) heterogeneity across the instrument-exposure effects (Q-statistic at P < 0.05 indicating heterogeneity, **Supplementary Table 33**).

We applied 5 different MR methods: Inverse variance weighted (IVW) using the random effects model, MR-Egger, simple mode, weighted median and weighted mode methods. When only a single SNP was available the Wald ratio (WR) test was conducted. MR analysis was conducted in R using the ‘mr()’ function in the ‘TwoSampleMR’ package^101^.

Subsequently, Radial MR analysis was conducted to determine if Wald ratio outliers needed to be removed from the IVW or MR-Egger MR estimates^37^. In addition, we conducted a Q-test to identify outlier SNPs (P < 0.05). These outliers were then removed from the original MR analyses (across all 5 MR methods). The Radial MR analysis was conducted using the RadialMR R package (https://github.com/WSpiller/RadialMR). In order to determine that the MR effects were orientated in the correct direction (from exposure to ALS) we conducted both reverse MR^102^ and Steiger filtering^103^ on our top MR findings.

Finally, we explored whether the MR effects of our total and LDL cholesterol and systolic blood pressure exposures may be confounded by the effect we observed for years of schooling by conducting multivariate MR analysis^104^. Conditional F and Q statistics were calculated using the ‘MVMR’ package^105^ in R.

## Data availability

All summary statistics will be made publicly available in centralized repositories upon publication.

## Consortium members

### SLALOM Consortium

Ettore Beghi^42^, Elisabetta Pupillo^42^, Giancarlo Comi^140^, Nilo Riva^140^, Christian Lunetta^141^, Francesca Gerardi^141^, Maria Sofia Cotelli^142^, Fabrizio Rinaldi^142^, Luca Chiveri^143^, Maria Cristina Guaita^144^, Patrizia Perrone^144^, Mauro Ceroni^145^, Luca Diamanti^145^, Carlo Ferrarese^146^, Lucio Tremolizzo^146^, Maria Luisa Delodovici^147^ & Giorgio Bono^147^

**Affiliations**

42: Laboratory of Neurological Diseases, Department of Neuroscience, Istituto di Ricerche Farmacologiche Mario Negri IRCCS, Milan, Italy.

140: IRCCS San Raffaele Hospital, Milan, Italy.

141: NEMO Clinical Center, Serena Onlus Foundation, Niguarda Ca’ Granda Hospital, Milan, Italy. 142: Civil Hospital of Brescia, Brescia, Italy.

143: Ospedale Valduce, Como, Italy.

144: A.O. Ospedale Civile di Legnano, Legnano, Italy.

145: IRCCS Istituto Neurologico Nazionale “C.Mondino”, Pavia, Italy.

146: A.O. “San Gerardo” di Monza and University of Milano-Bicocca, Italy 147: A.O. “Ospedale di Circolo Fondazione Macchi” di Varese, Varese, Italy.

### PARALS Consortium

Adriano Chiò^40,41^, Andrea Calvo^40,41^, Cristina Moglia^40,41^, Antonio Canosa^40,41,148^, Umberto Manera^40^, Rosario Vasta^40^, Alessandro Bombaci^40^, Maurizio Grassano^40^, Maura Brunetti^40^, Federico Casale^40^, Giuseppe Fuda^40^, Paolina Salamone^40^, Barbara Iazzolino^40^, Laura Peotta^40^, Paolo Cugnasco^40^, Giovanni De Marco^41^, Maria Claudia Torrieri^40^, Francesca Palumbo^40^, Salvatore Gallone^41^, Marco Barberis^149^, Luca Sbaiz^149^, Salvatore Gentile^150^, Alessandro Mauro^40,151^, Letizia Mazzini^152,153^, Fabiola De Marchi^152,153^, Lucia Corrado^154,153^, Sandra D’Alfonso^154,153^, Antonio Bertolotto^155^, Maurizio Gionco^156^, Daniela Leotta^157^, Enrico Odddenino^157^, Daniele Imperiale^158^, Roberto Cavallo^159^, Pietro Pignatta^160^, Marco De Mattei^161^, Claudio Geda^162^, Diego Maria Papurello^163^, Graziano Gusmaroli^164^, Cristoforo Comi^165,166^, Carmelo Labate^167^, Luigi Ruiz^168^, Delfina Ferrandi^169^, Eugenia Rota^170^, Marco Aguggia^171^, Nicoletta Di Vito^171^, Piero Meineri^172^, Paolo Ghiglione^173^, Nicola Launaro^174^, Michele Dotta^175^, Alessia Di Sapio^176^ & Guido Giardini^177^

**Affiliations**

40: “Rita Levi Montalcini” Department of Neuroscience, ALS Centre, University of Torino, Turin, Italy. 41: Neurologia 1, Azienda Ospedaliero Universitaria Città della Salute e della Scienza, Turin, Italy.

148: Azienda Ospedaliero-Universitaria Città della Salute e della Scienza di Torino, Neurology Unit 1U, Turin, Italy.

149: Department of Medical Genetics, Azienda Ospedaliero Universitaria Città della Salute e della Scienza, Turin, Italy.

150: Neurologia 3, Azienda Ospedaliero Universitaria Città della Salute e della Scienza di Torino, Turin, Italy.

151: Istituto Auxologico Italiano, IRCCS, Piancavallo, Italy.

152: Department of Neurology, ‘Amedeo Avogadro’ University of Piemonte Orientale, Novara, Italy. 153: Azienda Ospedaliero Universitaria ‘Maggiore della Carità’, Novara, Italy.

154: Department of Health Sciences, ‘Amedeo Avogadro’ University of Piemonte Orientale, Novara, Italy.

155: Department of Neurology and Multiple Sclerosis Center, Azienda Ospedaliero Universitaria San Luigi, Orbassano, Italy.

156: Department of Neurology, Azienda Ospedaliera ‘Ordine Mauriziano’ di Torino, Turin, Italy. 157: Department of Neurology, Ospedale Martini, ASL Città di Torino, Turin, Italy.

158: Department of Neurology, Ospedale Maria Vittoria, ASL Città di Torino, Turin, Italy.

159: Department of Neurology, Ospedale San Giovanni Bosco, ASL Città di Torino, Turin, Italy. 160: Ospedale Humanitas Gradenigo, Turin, Italy.

161: Department of Neurology, Ospedale ‘Santa Croce’ di Moncalieri, ASL Torino 5, Moncaliari, Italy. 162: Department of Neurology, Ospedale Civile di Ivrea, ASL Torino 4, Ivrea, Italy.

163: Department of Neurology, Presidio Ospedaliero di Ciriè, ASL Torino 4, Ciriè, Italy.

164: Department of Neurology, Ospedale ‘Degli Infermi’ di Biella, ASL Biella, Ponderano, Italy. 165: Department of Neurology, Ospedale ‘Sant’ Andrea’ di Vercelli, ASL Vercelli, Vercelli, Italy.

166: Department of Clinical and Experimental Medicine, ‘Amedeo Avogadro’ University of Piemonte Orientale, Novara, Italy.

167: Department of Neurology, Ospedale Civile ‘Edoardo Agnelli’ di Pinerolo, ALS Torino 2, Pinerolo, Italy.

168: Department of Neurology, Azienda Ospedaliera ‘Santi Antonio e Biagio’ di Alesssandria, Alessandria, Italy.

169: Department of Neurology, Ospedale ‘Santo Spirito’ di Casale Monferrato, ASL Alessandria, Casale Monferrato, Italy.

170: Department of Neurology, Ospedale ‘San Giacomo’ di Novi Ligure, ASL Alesssandria, Novi Ligure, Italy.

171: Department of Neurology, Ospedale ‘Cardinal Massia’ di Asti, ASL Asti, Asti, Italy.

172: Department of Neurology, Azienda Ospedaliera ‘Santa Croce e Carle’ di Cuneo, Cuneo, Italy.

173: Department of Neurology, Ospedale ‘Maggiore Santissima Annuziata’ di Savigliano, ASL Cuneo 1, Savigliano, Italy.

174: Department of Anestesiology, Ospedale ‘Maggiore Santissima Annuziata’ di Savigliano, ASL Cuneo 1, Savigliano, Italy.

175: Department of Neurology, Ospedale ‘Michele e Pietro Ferrero” di Verduno, ASL Cuneo 2, Verduno, Italy.

176: Department of Neurology, Ospedale ‘Regina Montis Regalis’ di Mondovì, ASL Cuneo 1, Italy. 177: Department of Neurology, Ospedale Regionale ‘Umberto Parini’ di Aosta, Aosta, Italy.

### SLAGEN Consortium

Vincenzo Silani^17,18^, Nicola Ticozzi^17,18^, Antonia Ratti^17,30^, Isabella Fogh^14^, Cinzia Tiloca^17^, Silvia Peverelli^17^, Cinzia Gellera^31^, Giuseppe Lauria Pinter^32,33^, Franco Taroni^178^, Viviana Pensato^178^, Barbara Castellotti^178^, Giacomo P. Comi^34,18^, Stefania Corti^34,18^, Roberto Del Bo^34,18^, Cristina Cereda^35^, Mauro Ceroni^179,180^, Stella Gagliardi^35^, Sandra D’ Alfonso^36^, Lucia Corrado^36^, Letizia Mazzini^181^, Gianni Sorarù^37^, Flavia Raggi^37^, Gabriele Siciliano^38^, Costanza Simoncini^38^, Annalisa Lo Gerfo^38^, Massimiliano Filosto^39^, Maurizio Inghilleri^182^ & Alessandra Ferlini^183^,

**Affiliations**

14: Maurice Wohl Clinical Neuroscience Institute, Department of Basic and Clinical Neuroscience, Institute of Psychiatry, Psychology & Neuroscience, King’s College London, London, UK.

17: Department of Neurology-Stroke Unit and Laboratory of Neuroscience, Istituto Auxologico Italiano IRCCS, Milan, Italy.

18: Department of Pathophysiology and Transplantation, “Dino Ferrari” Center, Università degli Studi di Milano, Milan, Italy.

30: Department of Medical Biotechnology and Translational Medicine, Università degli Studi di Milano, Milan, Italy.

31: Unit of Medical Genetics and Neurogenetics, Fondazione IRCCS Istituto Neurologico “Carlo Besta”, Milan, Italy.

32: 3rd Neurology Unit, Motor Neuron Diseases Center, Fondazione IRCCS Istituto Neurologico “Carlo Besta”, MIlan, Italy.

33: ‘L. Sacco’ Department of Biomedical and Clinical Sciences, Università degli Studi di Milano, Milan, Italy.

34: Neurology Unit, IRCCS Foundation Ca’ Granda Ospedale Maggiore Policlinico, Milan, Italy. 35: Genomic and Post-Genomic Center, IRCCS Mondino Foundation, Pavia, Italy.

36: Department of Health Sciences, University of Eastern Piedmont, Novara, Italy. 37: Department of Neurosciences, University of Padova, Padova, Italy.

38: Department of Clinical and Experimental Medicine, University of Pisa, Pisa, Italy.

39: Department of Clinical and Experimental Sciences, University of Brescia, Brescia, Italy.

178: Unit of Genetics of Neurodegenerative and Metabolic Diseases, Fondazione IRCCS Istituto Neurologico ‘Carlo Besta’, Milan, Italy.

179: Unit of General Neurology, IRCCS Mondino Foundation, Pavia, Italy.

180: Department of Brain and Behavioural Sciences, University of Pavia, Pavia, Italy

181: ALS Center, Department of Neurology, Azienda Ospedaliero Universitaria Maggiore della Carità, Novara, Italy

182: Rare Neuromuscular Diseases Centre, Department of Human Neuroscience, Sapienza University, Rome, Italy

183: Unit of Medical Genetics, Department of Medical Science, University of Ferrara, Ferrara, Italy.

### SLAP Consortium

Giancarlo Logroscino^43^, Ettore Beghi^42^, Isabella L. Simone^184^, Bruno Passarella^185^, Vito Guerra^186^, Stefano Zoccolella^187^, Cecilia Nozzoli^185^, Ciro Mundi^188^, Maurizio Leone^189^, Michele Zarrelli^189^, Filippo Tamma^190^, Francesco Valluzzi^191^, Gianluigi Calabrese^192^, Giovanni Boero^193^ & Augusto Rini^185^

**Affiliations**

42: Laboratory of Neurological Diseases, Department of Neuroscience, Istituto di Ricerche Farmacologiche Mario Negri IRCCS, Milan, Italy.

43: Department of Clinical Research in Neurology, University of Bari at “Pia Fondazione Card G. Panico” Hospital, Bari, Italy.

184: Department of Basic Medical Sciences, Neurosciences and Sense Organs, University of Bari, Bari, Italy.

185: Neurological Department, Antonio Perrino’s Hospital, Brindisi, Italy.

186: National Institute of Digestive Diseases. IRCCS S. de Bellis Research Hospital, Castellana Grotte, Italy.

187: ASL Bari, San Paolo Hospital, Milan, Italy.

188: Department of Neuroscience, United Hospital of Foggia, 71100 Foggia, Italy.

189: Unit of Neurology, Department of Medical Sciences, IRCCS Casa Sollievo della Sofferenza, 71013 San Giovanni Rotondo, Italy.

190: Neurology Unit, Miulli Hospital, Acquaviva delle Fonti, BA, Italy. 191: Unit of Neurology, “S. Giacomo” Hospital, Bari, Italy.

192: Department of Neurology, ASL (Local Health Authority) at the “V Fazzi” hospital, 73100 Lecce, Italy.

193: Department of Neurology, ASL (Local Health Authority) at the “SS. Annunziata” hospital, Taranto, Italy.

## Acknowledgments

W.v.R. is supported by funding provided by the Dutch Research Council (NWO) [VENI scheme grant 09150161810018] and Prinses Beatrix Spierfond (neuromuscular fellowship grant W.F19-03). J.J.F.A.v.V. is funded by Projectnumber W.OR20-08 (The “Repeatome” as a basis for new treatments of ALS) of the Prinses Beatrix Spierfonds. K.P.K. is supported by funding provided by the Dutch Research Council (NWO) [VIDI grant 91719350]. G.S. was supported by a PhD studentship from the Alzheimer’s Society. E.H. and J.M. were supported by Medical Research Council (MRC) grant K013807 (awarded to J.M). mQTL SMR data analysis was undertaken using high-performance computing supported by a Medical Research Council (MRC) Clinical Infrastructure award M008924 (awarded to J.M.). French ALS patients of the Pitié-Salpêtrière hospital (Paris) have been collected with ARSla funding support. D.B. and T.R.G. received funding from Biogen and UK Medical Research Council (MRC Epidemiology Unit, MC_UU_00011/1 and MC_UU_00011/4) for this project. G.D.S. works in the Medical Research Council Integrative Epidemiology Unit at the University of Bristol MC_UU_00011/1. D.B., E. Tsai and H.R. are employees of Biogen. J.P.R. is funded by the Canadian Institutes of Health Research (FRN 159279). A.A.K is supported by The Motor Neurone Disease Association (MNDA) and NIHR Maudsley Biomedical Research Centre. R.J.P. is supported by the Gravitation program of the Dutch Ministry of Education, Culture, and Science and the Netherlands Organization for Scientific Research (NWO; BRAINSCAPES). Project MinE Belgium was supported by a grant from IWT (n° 140935), the ALS Liga België, the National Lottery of Belgium and the KU Leuven Opening the Future Fund (awarded to P.V.D.). P.V.D holds a senior clinical investigatorship of FWO-Vlaanderen and is supported by the E. von Behring Chair for Neuromuscular and Neurodegenerative Disorders, the ALS Liga België and the KU Leuven funds “Een Hart voor ALS”, “Laeversfonds voor ALS Onderzoek” and the “Valéry Perrier Race against ALS Fund”. Several authors of this publication are members of the European Reference Network for Rare Neuromuscular Diseases (ERN-NMD). The authors are pleased to acknowledge the contribution of “Live now” Charity Foundation and Moscow ALS palliative care service for supporting patients with ALS and their families. G.A.R is supported by the Canadian Institutes of Health. Research Australia including its Ice Bucket Challenge Grant. We acknowledge funding from the National Health and Medical Research Council (NHMRC) (1078901, 1083187, 1113400, 1095215, 1121962, 1173790, Enabling Grant #402703). The Older Australian Twins Study (OATS, used for controls) acknowledges funding from the NHMRC/Australian Research Council Strategic Award (401162) and NHMRC (1405325, 1024224, 1025243, 1045325, 1085606, 568969, 1093083). OATS was facilitated through access to Twins Research Australia, a national resource supported by a NHMRC Centre of Research Excellence Grant (1079102). The Sydney Memory and Ageing Study (Sydney MAS, used for controls) has been funded by three NHMRC Program Grants (350833, 568969, and 1093083). We also acknowledge the OATS and Sydney MAS research teams: https://cheba.unsw.edu.au/research-projects/sydney-memory-and-ageing-study; https://cheba.unsw.edu.au/project/older-australian-twins-study. D.C.W. is supported by a Research Fellowship [APP1155413] from the National Health and Medical Research Council of Australia (NHMRC). The QSkin Study is supported by Grants [APP1185416, APP1073898, APP1063061] from the National Health and Medical Research Council of Australia (NHMRC). Several authors of this publication are members of the Netherlands Neuromuscular Center (NL-NMD) and the European Reference Network for rare neuromuscular diseases EURO-NMD. PJS is supported as an NIHR Senior Investigator and by the Sheffield NIHR Biomedical Research Centre. This is in part an EU Joint Programme -Neurodegenerative Disease Research (JPND) project. The project is supported through the following funding organisations under the aegis of JPND -www.jpnd.eu (United Kingdom, Medical Research Council (MR/L501529/1; MR/R024804/1) and Economic and Social Research Council (ES/L008238/1)) and through the Motor Neurone Disease Association. This study represents independent research part funded by the National Institute for Health Research (NIHR) Biomedical Research Centre at South London and Maudsley NHS Foundation Trust and King’s College London. A.A-C is supported by an NIHR Senior Investigator Award. Samples used in this research were in part obtained from the UK National DNA Bank for MND Research, funded by the MND Association and the Wellcome Trust. We would like to thank people with MND and their families for their participation in this project. We acknowledge sample management undertaken by Biobanking Solutions funded by the Medical Research Council at the Centre for Integrated Genomic Medical Research, University of Manchester. L.H.v.d.B. reports grants from The Netherlands Organization for Health Research and Development (Vici scheme), grants from The European Community’s Health Seventh Framework Programme (grant agreement n° 259867 (EuroMOTOR)), grants from The Netherlands Organization for Health Research and Development) the STRENGTH project, funded through the EU Joint Programme – Neurodegenerative Disease Research, JPND). This project has received funding from the European Research Council (ERC) under the European Union’s Horizon 2020 research and innovation programme (grant agreement n° 772376 – EScORIAL). The collaboration project is co-funded by the PPP Allowance made available by Health∼Holland, Top Sector Life Sciences & Health, to stimulate public-private partnerships. This study was supported by the ALS Foundation Netherlands

## Author contributions

**Sample ascertainment:** W.v.R, R.A.A.v.d.S, M.M, A.M.D, H-J. Westeneng, G.H.P.T, N.T, J.C-K, B.N.S, M.G, S.C, S.P, K.E.M, P.J.S, J.H, R.W.O, M.S, T.M, N.B, A.J.v.d.K, A.R, C.G, G.L.P, G.P.C, C.C, D.S, S.D’ A, G. Sorarù, G. Siciliano, M.F, A.P, A.C, A. Calvo, C.M, M.B, A. Canosa, M. Grassano, E.B, E.P, G.L, B.N, A.O, A.N, Y.L, M.Z, M. Gotkine, R.H.B, S.B, P.V’h, P.C, P. Couratier, S.M, V.M, F.S, J.S.M.P, A.A, R.R-G, P. Dion, J.P.R, A.C.L, J.H.W, D. Brenner, A.F, G.B, A.B, A.D, C.A.M.P, S.S-D, N.W, S.T, R. Rademakers, A. Braun, J.K, D.C.W, C.M.O, A.G.U, A.H, M.R, S. Cichon, M.M.N, P.A, B.T, A.B.S, M. Mitne Neto, R.J.C, R.A.O, M.W-P, C.L-H, V.M.v.D, J.G, A. Rödiger, N.G, A.J, T.B, E.T, B.I, B.S, O.W.W, R.S, C.A.H, C. Graff, L.B, V.F, V.D, A. Ataulina, B.R, B.K, J.Z, M.R-G, D.G, Z.S, V. Drory, M.P, I.P.B, M.C.K, R.D.H, S. Mathers, P.A.M, M.N, G.A.N, R.P, D.B.R, K.M, P.S.S, M.d.C, S. Pinto, S. Petri, A. Osmanovic, M.W, G.A.R, V.S, J. Glass, R.H. Brown, J.E.L, C.E.S, P.M.A, D.F, F.C.G, A.F.M, R.L.M, O.H, A.A-C, P.V.D, L.H.v.d.B, J.H.V, G.C, N.R, C.L, F.G, M.S.C, F.R, L.C, M.C.G, P.P, M.C, L.D, C.F, L. Tremolizzo, M.L.D, G. Bono, U.M, R.V, A. Bombaci, F.C, G.F, P.S, B. Iazzolino, L.P, P. Cugnasco, G.D.M, M.C.T, F.P, S.G, M. Barberis, L.S, S. Gentile, A.M, L.M, F.D.M, L. Corrado, A. Bertolotto, M. Gionco, D.L, E.O, D.I, R.C, P. Pignatta, M.D.M, C. Geda, D.M.P, G.G, C. Comi, C. Labate, L.R, D. Ferrandi, E.R, M.A, N.D.V, P.M, P.G, N.L, M. Dotta, A.D.S, G. Giardini, C.T, S. Peverelli,, F.T, V.P, B.C, S. Corti, R.D.B, C. Cereda, M. Ceroni, L. Mazzini, F. Raggi, C.S, A.L.G, M.I, A. Ferlini, I.L.S, B.P, V.G, S.Z, C.N, C. Mundi, M.L, M. Zarrelli, F. Tamma, F.V, G. Calabrese, G. Boero & A. Rini **SNP-array genotyping:** W.v.R, R.A.A.v.d.S, A.M.D, A.S, I.F, G.B, A.B, A.D, C.A.M.P, S.S-D, N.W, L.T, W.L, A. Franke, S.R, A. Braun, J.K, D.C.W, C.M.O, A.G.U, A.H, M.R, S. Cichon, M.M.N, P.A, B.T, A.B.S, B.B, S.F, S.T.N, F.J.S, K.L.W, A.K.H, L.W, C. Curtis, G. Breen, D.F, F.C.G, A.F.M, N.R.W, A.A-C, P.V.D, L.H.v.d.B & J.H.V **GWAS quality control:** W.v.R, R.A.A.v.d.S, M.K.B, R.R, R.L.M, N.R.W and J.H.V. **GWAS data analysis:** W.v.R, R.A.A.v.d.S, M.K.B, R.R, R.P.B, M.D, M.H, A.A.K, A.I, A.S, N.T, B.N.S, B.B, D.F, A.F.M, R.L.M, N.R.W and J.H.V. **Whole-genome sequencing:** W.v.R, R.A.A.v.d.S, P.J.H, R.A.J.Z, M.M, A.M.D, G.H.P.T, K.R.v.E, M.K, J.C-K, B.N.S, K.P.K, A.A-C, P.V.D, L.H.v.d.B and J.H.V. **WGS quality-control:** W.v.R, R.A.A.v.d.S, J.J.F.A.v.V, P.J.H, R.A.J.Z, M.M, K.P.K, P.V.D and J.H.V. **WGS rare-variant burden analyses:** W.v.R, R.A.A.v.d.S, P.J.H, R.A.J.Z, K.R.v.E, K.P.K, P.V.D and J.H.V. **WGS STR-analyses:** W.v.R, J.J.F.A.v.V, R.A.J.Z, E.D, M.A.E and J.H.V. **eQTL analyses:** W.v.R, R.A.A.v.d.S, M.K.B, N.d.K, H-J.W, O.B.B, P.D, J.M, L.F and J.H.V. **mQTL analyses:** W.v.R, M.K.B, P.J.H, R.A.J.Z, G.S, E.H, A.M.D and J.H.V. **Cross-disorder analyses:** W.v.R, R.A.A.v.d.S, M.K.B, N.d.K, H-J.W, O.B.B, P.D, E.J.N.G, M.A.v.E, R.J.P, A.F.M, N.R.W, E. Tsai, H.R, L.F and J.H.V. **MR analyses:** W.v.R, R.A.A.v.d.S, M.K.B, D.B, H.J.W, G.D.S, T.R.G, E. Tsai, H.R, and J.H.V. **Writing of the manuscript:** W.v.R, M.K.B, D.B, J.M, E. Tsai and J.H.V. **Revising the manuscript:** W.v.R, R.A.A.v.d.S, M.K.B, J.J.F.A.v.V, G.S, E.H, D.B, R.R, E.D, H.J.W, G.H.P.T, K.R.v.E, E.J.N.G, M.A.v.E, R.J.P, G.D.S, T.R.G, R.L.M, K.P.K, N.R.W, E. Tsai, H.R, L.F, L.H.v.d.B and J.H.V. **Acquired funding and supervised the study:** L.H.v.d.B and J.H.V.

## Competing Interests

J.H.V has sponsored research agreements with Biogen Idec. L.H.v.d.B receives personal fees from Cytokinetics, outside the submitted work. A.A-C. has served on scientific advisory boards for Mitsubishi Tanabe Pharma, OrionPharma, Biogen Idec, Lilly, GSK, Apellis, Amylyx, and Wave Therapeutics. A.C. serves on scientific advisory boards for Mitsubishi Tanabe, Roche, Biogen, Denali, and Cytokinetics.

## Notes

### Competing Interest Statement

Jan Veldink has sponsored research agreements with Biogen Idec. Leonard van den Berg receives personal fees from Cytokinetics, outside the submitted work. Ammar Al-Chalabi has served on scientific advisory boards for Mitsubishi Tanabe Pharma, OrionPharma, Biogen Idec, Lilly, GSK, Apellis, Amylyx, and Wave Therapeutics. Adriano Chio serves on scientific advisory boards for Mitsubishi Tanabe, Roche, Biogen, Denali, and Cytokinetics.

### Author Declarations

Please see attached .tsv file as requested

