## Supplementary information for "Common and rare variant association analyses in Amyotrophic Lateral Sclerosis identify 15 risk loci with distinct genetic architectures and neuron-specific biology"

#### - Supplementary Data -

**Supplementary Information** 1
Description of newly genotyped GWAS cohorts 2
Cohort 66 cKingsOE - Illumina OmniExpress 2
Cohort 104 cKingsGSA - Illumina GSA 3
Cohort 107 cAUS - Illumina Exome 4
Cohort 117 Qskin - Illumina GSA 4
Cohort 118 Sripke - Illumina GSA 4
Cohort 119 and 120 cKingsGSA2 and cErasmusGSA - Illumina GSA 5
Description of cohorts in Project MinE whole-genome sequencing. 7
Consortium members 9

**Supplementary Figures** 10

**Supplementary Tables** 59

**References** 75

### Supplementary Information

#### Description of newly genotyped GWAS cohorts

##### Cohort 66 cKingsOE - Illumina OmniExpress

**Germany (Grosskreutz).** ALS patients and controls were recruited at the Thuringian Neuromuscular Center, Jena University Hospital, Germany. Written informed consent was obtained. ALS patients were diagnosed according to the El-Escorial criteria of definite, probable or laboratory-supported probable ALS. Control subjects were population based controls matched for sex and age. All participants gave written informed consent and the Local ethics committee of the Medical Faculty of Friedrich Schiller University Jena approved this protocol.

**Italy - Milan (Beghi).** ALS patients enrolled were residents of the Lombardy Region, Northern Italy, and were primarily diagnosed in or referred to secondary and tertiary ALS centers included in the population based registry (SLALOM). Patients were not pre-screened for any mutations related to ALS. Control subjects were selected from the lists of the general practitioners of the companion cases and were matched for sex, age and geographic region. The structure of the SLALOM Registry has been described previously[1]. All participant gave written informed consent and the Ethical Committee of Città della Salute Hospital approved this protocol.

**Italy - Turin (Chio).** ALS patients were included in the Piemonte and Valle d'Aosta ALS Register (PARALS), north western Italy. Patients were not pre-screened for any mutations related to ALS. Control subjects were population based controls matched for sex, age and geographic region within Piemonte and Valle d'Aosta. The PARALS has been described in detail previously[2]. All participant gave written informed consent and the Ethical Committee of Città della Salute Hospital approved this protocol.

**Italy - Bari (Logroscino).** ALS patients were enrolled as part of the SLAP registry, a population-based registry including all ALS patients who were residents of Puglia region, SouthEast Italy. All ALS cases were diagnosed or referred to a network of diagnostic facilities including all secondary and tertiary ALS centers present in the geographic area. The base population was the population of Puglia in the same period according to the national census. Patients were not pre-screened for any mutations related to ALS. Control subjects were selected from the lists of the general practitioners (GPs) of the cases and were matched for sex, age, sex, and place of residence. The SLAP Registry has been described in detail previously[3]. All individuals gave written informed consent and the Institutional Review Board of the Azienda Sanitaria Locale, Lecce approved this protocol.

**Sweden (Graff)** This cohort included patients with ALS recruited at the Memory Clinic, Karolinska University Hospital, Huddinge, Sweden, after 1992. Patients were diagnosed with ALS according to the revised El-Escorial criteria for ALS. All individuals gave written informed consent and the Ethics Committee Stockholm approved this protocol.

**The Netherlands (Veldink, van den Berg)**. ALS patients were diagnosed with ALS at the tertiary referral clinic for motor neuron disease at the University Medical Center Utrecht (Dutch ALS Center) or were included in the Prospective ALS Study in The Netherlands. Patients were not pre-screened for any mutations related to ALS. Control subjects were population based controls matched for sex, age and geographic region within the Netherlands. The Prospective ALS study in The Netherlands has been described in detail previously[4]. All individuals gave written informed consent and the University Medical Center Utrecht Medical Ethics Committee, Utrecht approved this protocol.

#### Cohort 104 cKingsGSA - Illumina GSA

**Canada (Rouleau)**. ALS patients and unaffected controls were recruited from neurological clinics across Québec, Canada. Control subjects were recruited from various studies, either as the unaffected parents of psychiatric probands, unaffected relatives of probands of non-ALS neurological diseases, and unaffected individuals recruited specifically as controls. Samples were included without selection for variant carrier status, age, reported sex, or reported ethnicity. All individuals signed informed consent according to the relevant institutional ethics protocol. All individuals gave written informed consent and the Review Ethics Board Office at McGill University, Montreal approved this protocol.

**France - Paris (Millecamps, Meininger, Salachas)**. ALS patients were diagnosed with probable or definite ALS according to the revised El Escorial criteria between 1996 and 2004 at the national ALS reference center of the Pitié-Salpêtrière hospital (Paris). These patients were cases with sporadic occurrence of the disease (with no evident familial history). Control subjects were individuals of European ancestries with a French background (healthy spouses and husbands) matched for age and sex collected at the same period. All individuals gave written informed consent and the Medical Research Ethics Committee of “Assistance Publique-Hôpitaux de Paris” approved this protocol.

**France - Tours (Vourc’h, Corcia, Couratier).** ALS patients were diagnosed with probable or definite ALS according to the El Escorial Criteria by neurologists specialized in motor neuron diseases at the Reference centers for ALS of the University Hospitals of Limoges and Tours (LITORALS federation), members of the French FILSLAN networks. All individuals gave written informed consent and the ethics committee of Tours Hospital and Limoges University Hospital this protocol.

**Slovenia (Koritnik, Rogelj, Zidar, Ravnik-Glavač, Glavač).** ALS patients were diagnosed at the tertiary Ljubljana ALS Centre which takes care of the majority of Slovenian ALS patients. Control samples were healthy individuals matched for gender and age and were unrelated to ALS patients. More details were described previously[5]. All individuals gave written informed consent and the National Medical Ethics Committee of Republic of Slovenia approved this protocol.

**The Netherlands (Veldink, van den Berg).** ALS patients were diagnosed with ALS at the tertiary referral clinic for motor neuron disease at the University Medical Center Utrecht (Dutch ALS Center) or were included in the Prospective ALS Study in The Netherlands. Patients were not pre-screened for any mutations related to ALS. Control subjects were population based controls matched for sex, age and geographic region within the Netherlands. The Prospective ALS study in The Netherlands has been described in detail previously[4]. All individuals gave written informed consent and the University Medical Center Utrecht Medical Ethics Committee, Utrecht approved this protocol.

#### Cohort 107 cAUS - Illumina Exome

**Australia (Wray, Blair, Kiernan)**. This cohort includes the University of Sydney's Australian Motor Neuron Disease DNA Bank (MND Bank) cohort recruited April 2000 to June 2011 (493 cases, 497 controls), with study protocol approved by the Sydney South West Area Health Service Human Research Ethics Committee (HREC). Cases were recruited from around Australia via state-based MND associations with diagnosis verified by a neurologist. The remainder of the cases (N=467) were recruited from clinics across Australia between 2015 and 2017 under HREC approvals from Royal Brisbane and Women's Hospital (RBWH; N=220), Macquarie University Multidisciplinary Motor Neurone Disease Clinic (N=205), Calvary Health Care Bethlehem in Melbourne (N=32), Fiona Stanley Hospital in Perth (N=10), and from 2016 under HREC approvals at each site for the sporadic ALS Australia Systems Genomics Consortium (SALSA-SGC). The ALS cases were diagnosed with definite or probable ALS according to the revised El Escorial criteria[6]. Some controls were recruited as either partners or friends of patients, healthy individuals free of neuromuscular diseases (N=166; N=82 from Macquarie, N=84 from RBWH). Additional controls were included from the Older Australian Twin Study (N=89, selected as single from a twin pair) from QIMR Berghofer Medical Research Institute, University of New South Wales and the University of Melbourne, and from the University of New South Wales Sydney Memory & Ageing Study (N=91), all approved by their respective HRECs. From this cohort N=846 cases and N=665 controls have data included in the dbGAP upload phs002068.v1.p1. All individuals gave written informed consent and the Sydney South West Area Health Service Human Research Ethics Committee (HREC) and HREC at the different sites: University of Sydney, Western Sydney Local Health District, Royal Brisbane and Women Hospital Metro North, South Metropolitan Health Service, Macquarie University, QIMR Berghofer Medical Research Institute, University of New South Wales and the University of Melbourne approved this protocol.

#### Cohort 117 Qskin - Illumina GSA

**Australia (Whiteman, Olsen).** The QSkin Sun and Health Study is a cohort of men and women aged 40–69 years randomly sampled from the population of Queensland, Australia in 2011 (ref[7,8]). The cohort was established to study the development of skin cancer and melanoma; the baseline survey collected demographic and health information. The study sample for these analyses were a subset of the cohort who provided a saliva sample; participants were not screened for ALS. All individuals gave written informed consent and the Human Research Ethics Committee at the QIMR Berghofer Medical Research Institute approved this protocol.

#### Cohort 118 Sripke - Illumina GSA

**Germany (Ripke).** Controls were collected as part of The Berlin Psychosis Study (BePS). This is a case-control sample initiated in greater Berlin aiming to facilitate the discovery of novel genetic variants associated with schizophrenia. The current sample consists of control samples of European ancestries. Control subjects were recruited into the study via local advertisement and participant databases. Individuals were excluded if they had ever been diagnosed with schizophrenia, schizoaffective disorder or bipolar disorder. All individuals gave written informed consent and the Charité Universitätsmedizin, Berlin Medical Ethics Committee approved this protocol.

#### Cohort 119 and 120 cKingsGSA2 and cErasmusGSA - Illumina GSA

The following cases and controls were randomized over these two cohorts.

**Canada (Rouleau).** ALS patients and unaffected controls were recruited from neurological clinics across Québec, Canada. Control subjects were recruited from various studies, either as the unaffected parents of psychiatric probands (n = 334), unaffected relatives of probands of non-ALS neurological diseases (n = 245), and unaffected individuals recruited specifically as controls (n = 164). Samples were included without selection for variant carrier status, age, reported sex, or reported ethnicity. All individuals gave written informed consent and the Review Ethics Board Office at McGill University, Montreal approved this protocol.

**France (Corcia, Vourc'h, Couratier).** ALS patients were diagnosed with probable or definite ALS according to the El Escorial Criteria by neurologists specialized in motor neuron diseases at the Reference centers for ALS of the University Hospitals of Limoges and Tours (LITORALS federation), members of the French FILSLAN networks. All individuals gave written informed consent and the ethics committee of Tours Hospital and Limoges University Hospital this protocol.

**Germany (Petri).** Blood was collected from sporadic and familial ALS cases at the ALS/MND Clinic of the Department of Neurology of Hannover Medical School, Germany. Each patient provided informed consent for participation in the study. All patients were examined at least once by a neurologist specialized in ALS. Extensive clinical workup including magnetic resonance imaging (MRI), cerebral spinal fluid analysis, electromyography (EMG), and nerve conduction studies (NCS) was performed to exclude ALS-mimicking conditions. Longitudinal information over a number of years was available for most individuals as described in ref [9,10]. All individuals gave written informed consent and the Medical Ethics Committee of Hannover Medical School approved this protocol.

**Ireland (McLaughlin, Hardiman).** Cases were diagnosed with probable or definite ALS according to the 1994 El-Escorial Criteria by neurologists specialized in motor neurone diseases at Beaumont Hospital in Dublin. Patients were referred from all regions in Ireland and were part of an ongoing population-based prospective ALS registry. Control samples were healthy individuals matched for geography, sex and age. They were either spouses, those accompanying patients to the ALS clinic or community-derived volunteers. All individuals reported Irish ancestry for at least three generations. All individuals gave written informed consent and the Beaumont Hospital Research & Ethics Committee, Dublin approved this protocol.

**Italy - Bari (Logroscino).** ALS patients were enrolled as part of the SLAP registry, a population-based registry including all ALS patients who were residents of Puglia region, SouthEast Italy. All ALS cases were diagnosed or referred to a network of diagnostic facilities including all secondary and tertiary ALS centers present in the geographic area. The base population was the population of Puglia in the same period according to the national census. Patients were not pre-screened for any mutations related to ALS. Control subjects were selected from the lists of the general practitioners (GPs) of the cases and were matched for sex, age, sex, and place of residence. The SLAP Registry has been described in detail previously[3]. All individuals gave written informed consent and the Institutional Review Board of the Azienda Sanitaria Locale, Lecce approved this protocol.

**Italy - Milan (Silani, Ticozzi).** Patients were diagnosed with ALS according to the El Escorial revised criteria by the SLAGEN Consortium, which includes Italian referral centers for motor neuron diseases. Patients were not pre-screened for any ALS-associated mutations. Control subjects were population-based controls matched for sex, age and geographic region within Italy. All individuals gave written informed consent and the Ethics Committee of the IRCCS Istituto Auxologico Italiano, Milan approved this protocol.

**The Netherlands (Veldink, van den Berg).** ALS patients were diagnosed with ALS at the tertiary referral clinic for motor neuron disease at the University Medical Center Utrecht (Dutch ALS Center) or were included in the Prospective ALS Study in The Netherlands. Patients were not pre-screened for any mutations related to ALS. Control subjects were population based controls matched for sex, age and geographic region within the Netherlands. The Prospective ALS study in The Netherlands has been described in detail previously[4]. All individuals gave written informed consent and the University Medical Center Utrecht Medical Ethics Committee, Utrecht approved this protocol.

**Russia (Brylev).** Patients were diagnosed with probable or definite ALS according to the 1994 El-Escorial Criteria by neurologists specialized in motor neuron diseases in Moscow ALS center and St-Petersburg ALS-care service. Patients were referred from all regions of Russia, and were not pre-screened for any mutations related to ALS. Control subjects were healthy individuals matched for sex and age, some of them spouses or those accompanying patients to the ALS clinic. All individuals gave written informed consent and the Local ethical committee of Buyanov city hospital, Moscow approved this protocol.

**Serbia (Stevic).** All samples were taken from patients seen at the Clinic of Neurology, School of Medicine, University of Belgrade, Serbia. The Clinic of Neurology is a tertiary center caring for a large number of ALS patients in Serbia. Diagnoses were made by neurologists specialized in neuromuscular diseases and motor neuron diseases. After informed consent, full demographic and clinical information was entered into the clinic database. All individuals gave written informed consent and the Ethics Committee of the School of Medicine at the University of Belgrade approved this protocol.

**UK (P. Shaw, Cooper-Knock).** All patients were reviewed by a senior consultant Neurologist and diagnosed with definite or probable ALS, as defined by the El Escorial criteria[6]. A detailed family history was taken from each patient. Population, age and sex matched control subjects were recruited from partners or unrelated carers of patients with ALS. The South Sheffield Research Ethics Committee approved the study, and informed consent was obtained for all samples. All individuals gave written informed consent and the Yorkshire and the Humber - Sheffield Research Ethics Committee approved this protocol.

**US - Cedar-Sinai (Baloh).** Cases were identified at the Cedar-Sinai ALS Clinic in Los Angeles. All cases met El Escorial criteria for definite or probable ALS. All individuals gave written informed consent and the Institutional review board of Cedars-Sinai, Los Angeles approved this protocol.

**US - UCLA (Ophoff).** Cases were identified at the ALS clinical centers of University of California Los Angeles and University of California San Francisco. Patients fulfilled the El-Escorial Criteria for definite or probable ALS. Control participants were population-based individuals. All individuals gave written informed consent and the Institutional review board of the University of California at Los Angeles approved this protocol.

#### Description of cohorts in Project MinE whole-genome sequencing.

**The Netherlands (Veldink, van den Berg)**. ALS patients were diagnosed with ALS at the tertiary referral clinic for motor neuron disease at the University Medical Center Utrecht (Dutch ALS Center) or were included in the Prospective ALS Study in The Netherlands. Patients were not pre-screened for any mutations related to ALS. Control subjects were population based controls matched for sex, age and geographic region within the Netherlands. The Prospective ALS study in The Netherlands has been described in detail previously[4]. All individuals gave written informed consent and the University Medical Center Utrecht Medical Ethics Committee, Utrecht approved this protocol.

**UK MNDA Biobank (C. Shaw, P. Shaw, Al-Chalabi, Morrison)**. Cases were diagnosed with ALS in one of 20 UK hospitals by neurologists specialized in motor neuron diseases. Patients had no family history for ALS. All participated in the UK National Biobank for Motor Neuron Disease Research. Patients had no family history for ALS and were of self-reported European descent. All individuals gave written informed consent and the Trent University Medical Ethics Committee approved this protocol.

**Turkey (Basak)**. ALS patients were recruited from hospitals across Turkey between 2002 and 2019. DNA samples were collected at the Boğaziçi University. A full description is provided in reference ref [11]. All individuals gave written informed consent and the Ethics Committee on Research with Human Participants (INAREK) at Bogazici University, Istanbul approved this protocol.

**Belgium (van Damme).** Patients were diagnosed with ALS at the tertiary referral clinic for motor neuron diseases at the University Hospitals in Leuven. Patients were pre-screened for mutations in *C9orf72*, *SOD1*, *TARDBP* and *FUS*, but were not always excluded in case of pathogenic mutation[12]. Control subjects were often spouses of patients, supplemented with age- and sex-matched controls from other local studies[13]. All individuals gave written informed consent and the Ethical Committee of University Hospital Leuven approved this protocol.

**Ireland (Hardiman, McLaughlin).** Cases were diagnosed with probable or definite ALS according to the 1994 El-Escorial Criteria by neurologists specialized in motor neurone diseases at Beaumont Hospital in Dublin. Patients were referred from all regions in Ireland and were part of an ongoing population-based prospective ALS registry. Patients were selected for sequencing such that all areas of Ireland were adequately represented. Control samples were neurologically healthy volunteers matched for geography, sex and age sampled from the community. All individuals reported Irish ancestry for at least three generations. All individuals gave written informed consent and the Beaumont Hospital Research & Ethics Committee, Dublin approved this protocol.

**Spain (Mora Pardina, Povedano).** ALS patients diagnosed with definite or probable ALS according to the El Escorial criteria. Patients were seen by neurologists and neurophysiologists at the tertiary referral centers: the Bellvitge hospital and Carlos III hospital for Catalonia and Madrid respectively. Controls were healthy individuals, without familial history of ALS, matched for age and sex. All individuals gave written informed consent and the Bellvitge University Hospital Ethics Committee, Barcelona and "Comité de Ética de la Investigación del Hospital Carlos III", Madrid approved this protocol.

**United States (Landers, Glass).** All samples were taken from patients seen at the Emory ALS Center in Atlanta, Georgia, USA. The Emory Center is a tertiary care ALS clinic caring for a large proportion of patients in Georgia and surrounding states[14]. Diagnoses were made by neurologists specialized in neuromuscular diseases and motor neuron diseases. After informed consent, full demographic and clinical information was stored into the clinic database. DNA was collected and stored. All individuals gave written informed consent and the Committee for the Protection of Human Subjects in Research of the University of Massachusetts Medical School, Worcester approved this protocol.

**France (Corcia, Couratier, Vourc'h).** ALS patients were diagnosed with probable or definite ALS according to the El Escorial Criteria by neurologists specialized in motor neuron diseases at the Reference centers for ALS of the University Hospitals of Limoges and Tours (LITORALS federation), members of the French FILSLAN networks. All individuals gave written informed consent and the ethics committee of Tours Hospital and Limoges University Hospital this protocol.

**Sweden (Andersen).** Cases were diagnosed with probable or definite ALS according to the revised El-Escorial Criteria by neurologists specialized in motor neuron diseases. Control individuals were free of any neuromuscular disease and matched for age, gender and ethnicity. Healthy controls were spouses of ALS patients or patients with other neurological diseases. All participants were from Swedish descent all reporting Northern Swedish citizenship for at least three generations. All individuals gave written informed consent and the Regional Ethical Review Board in Umeå approved this protocol.

**Israel (Gotkine, Drory)**. ALS patients were diagnosed with probable or definite ALS according to the El Escorial Criteria and in follow-up at the tertiary referral ALS clinic at the Hadassah University Hospital, Jerusalem or Tel-Aviv Sourasky Medical Center in Tel-Aviv. Patients were not pre-screened for any mutations related to ALS. Patients were referred from all regions in Israel and participated in a prospective ALS database and sample repository. All individuals gave written informed consent and the Hadassah

University Hospital Institutional Review Board, Hadassah and The Institutional Review Board of Tel Aviv Sourasky Medical Center, Tel Aviv and approved this protocol.

**Portugal (deCarvalho, Pinto).** Patients were diagnosed with possible, probable or definite ALS according to the revised El-Escorial criteria by neurologists specialized in motor neuron diseases. Both cases with and without a family history (third degree relatives) were included. Control subjects were spouses or those accompanying patients to the clinic. All individuals gave written informed consent and the Local Research Ethics Committee at the Faculty of Medicine, University of Lisbon, Lisbon approved this protocol.

**Italy (Chio, Ticozzi, Silani).** Patients were diagnosed with ALS according to the El Escorial revised criteria at the ALS tertiary referral center of Istituto Auxologico Italiano IRCCS. All patients had probable or definite familial ALS according to the Byrne criteria for FALS[15]. Patients were pre-screened for mutations in the *SOD1*, *TARDBP*, *FUS* and *C9orf72* genes. All individuals gave written informed consent and the Ethics Committee of the IRCCS Istituto Auxologico Italiano, Milan approved this protocol.

**Switzerland (Weber).** ALS patients were diagnosed at the Muskelzentrum/ALS clinic at the Kantonsspital St. Gallen, a tertiary referral center in Northern Switzerland. Patients fulfilled the El-Escorial Criteria for probable lab supported, probable or definite or ALS. Control subjects were healthy blood donors matched for age and gender. All individuals gave written informed consent and the Kantonale Ethikkomission des Kantons St. Gallen, St. Gallen approved this protocol.

### Supplementary Figures

**Supplementary Figure 1. Manhattan plot in European ancestries GWAS.** Loci containing a genome-wide significant SNP are highlighted in red. SNP IDs are the lead SNPs in each locus.

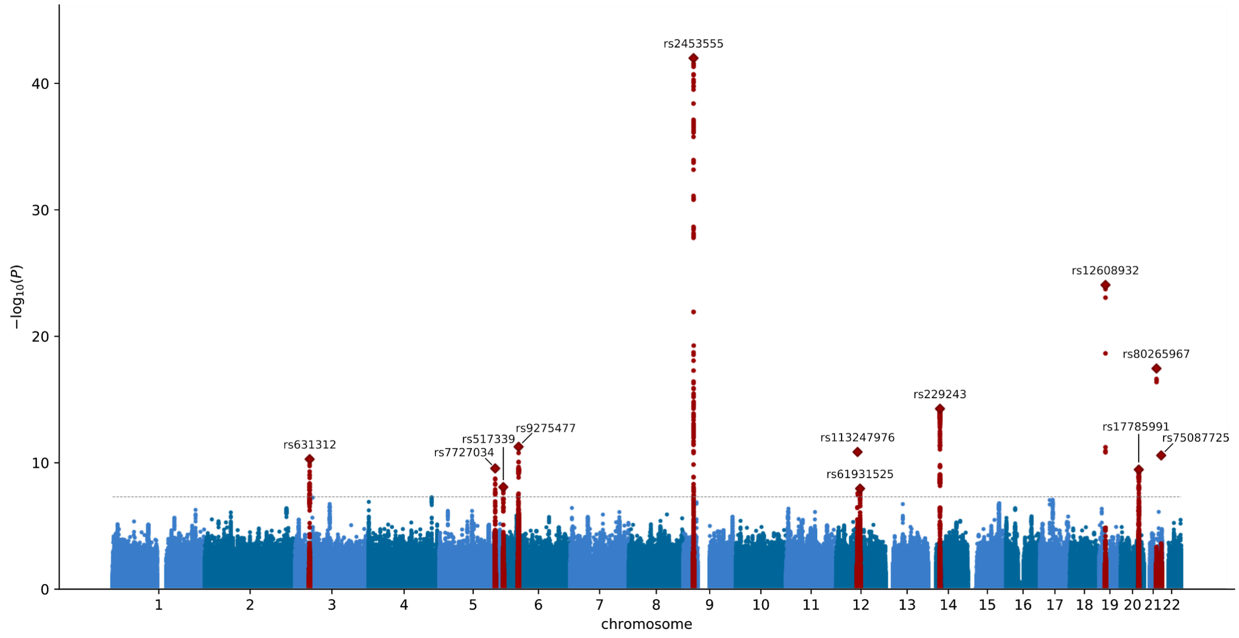


**Supplementary figures 2-16. Forest plots for genome-wide significant loci.** OR = odd-ratio, CI = confidence intervals. Effect allele is shown in the title of the forest plot.

**Supplementary figure 2. Forest plot for rs631312 (G), MOBP/*RPSA***

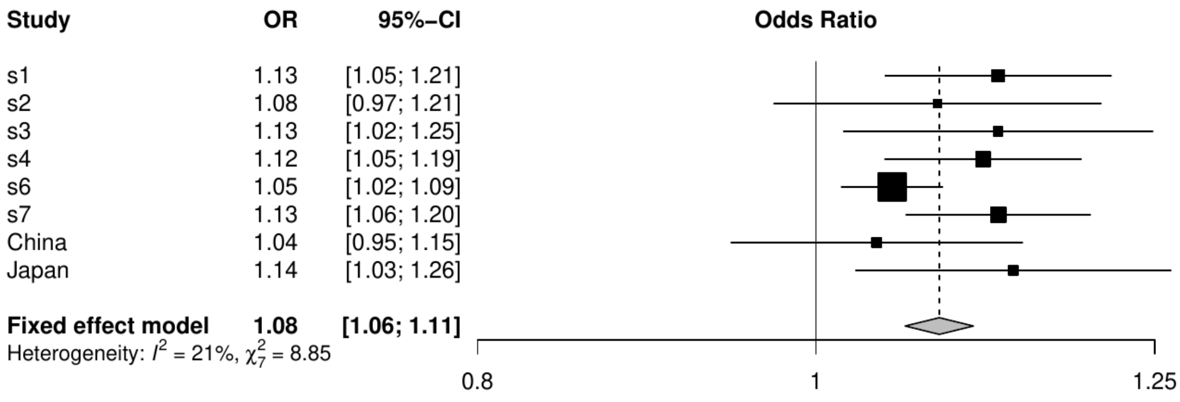

#### Supplementary figure 3. Forest plot for rs62333164 (A), *NEK1*

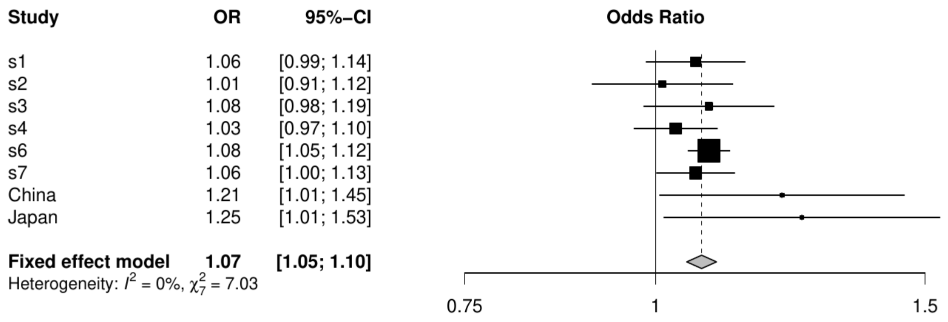


#### Supplementary figure 4. Forest plot for rs10463311 (C), *GPX3/TNIP1*

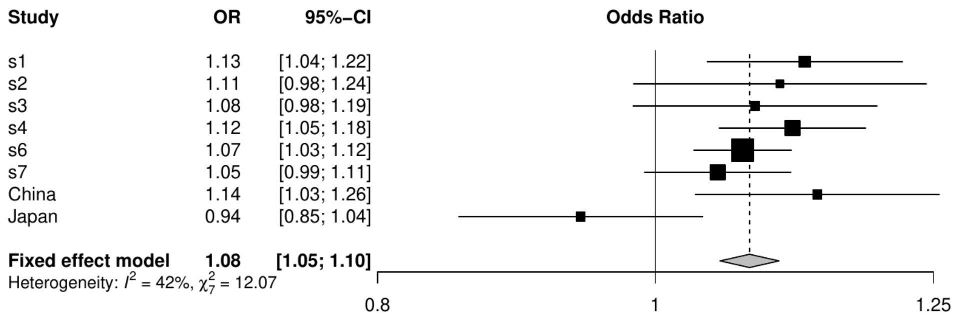


#### Supplementary figure 5. Forest plot for rs517339 (C), *ERGIC1*

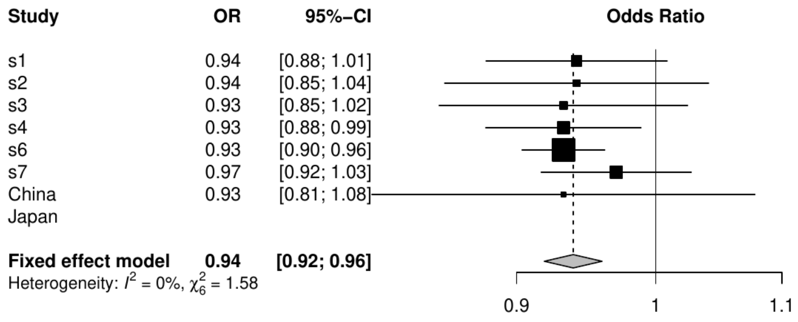

**Supplementary figure 6. Forest plot for rs9275477 (C), *HLA***

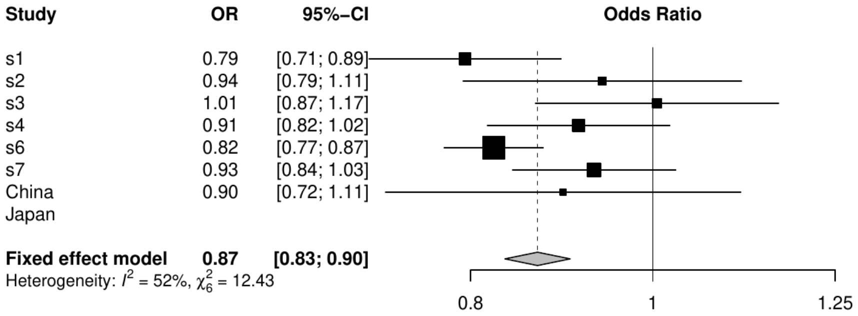


**Supplementary figure 7. Forest plot for rs10280711 (G), *PTPRN2***

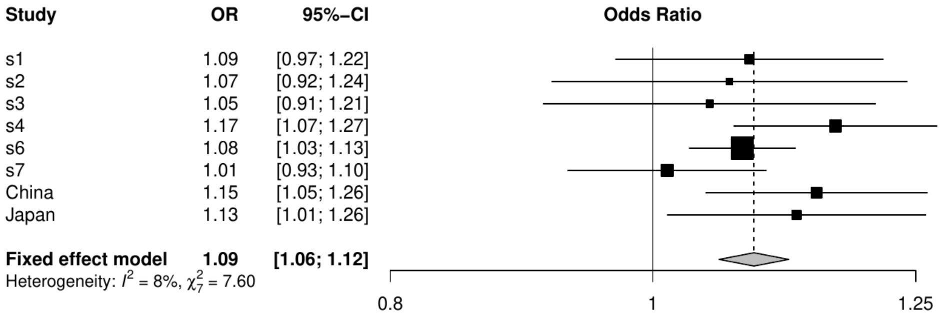


**Supplementary figure 8. Forest plot for rs2453555 (A), *C9orf72***

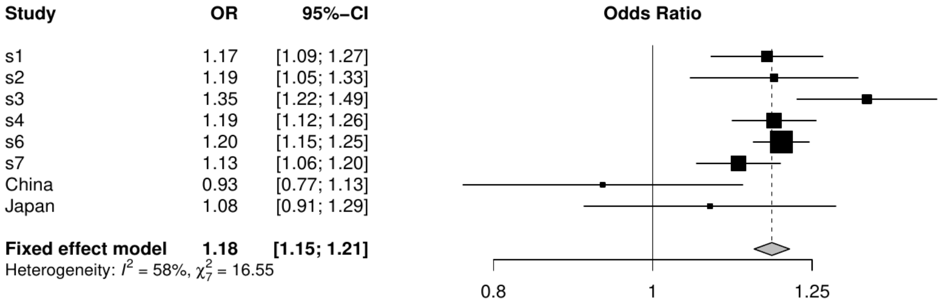

**Supplementary figure 9. Forest plot for rs113247976 (T), *KIF5A***

| Study | OR | 95%–CI |
|---|---|---|
| s1 | 1.30 | [0.95; 1.77] |
| s2 | 1.60 | [1.06; 2.41] |
| s3 | 1.31 | [0.98; 1.76] |
| s4 | 1.14 | [0.85; 1.53] |
| s6 | 1.46 | [1.27; 1.68] |
| s7 | 1.43 | [1.09; 1.89] |
| China | | |
| Japan | | |
| **Fixed effect model** | **1.39** | **[1.26; 1.54]** |

Heterogeneity: $I^2 = 0\%$, $\chi^2_5 = 3.13$

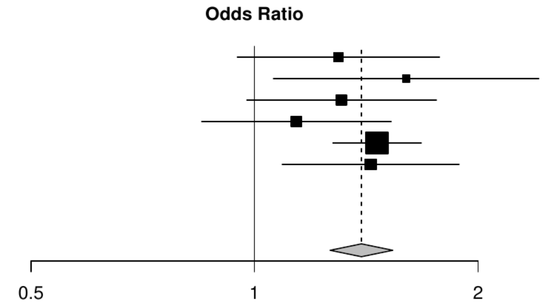


**Supplementary figure 10. Forest plot for rs4075094 (A), *TBK1***

| Study | OR | 95%–CI |
|---|---|---|
| s1 | 0.92 | [0.82; 1.04] |
| s2 | 0.97 | [0.83; 1.14] |
| s3 | 0.85 | [0.74; 0.98] |
| s4 | 0.93 | [0.86; 1.01] |
| s6 | 0.92 | [0.89; 0.96] |
| s7 | 0.83 | [0.76; 0.89] |
| China | 0.76 | [0.61; 0.94] |
| Japan | 0.90 | [0.67; 1.21] |
| **Fixed effect model** | **0.91** | **[0.88; 0.93]** |

Heterogeneity: $I^2 = 36\%$, $\chi^2_7 = 10.91$

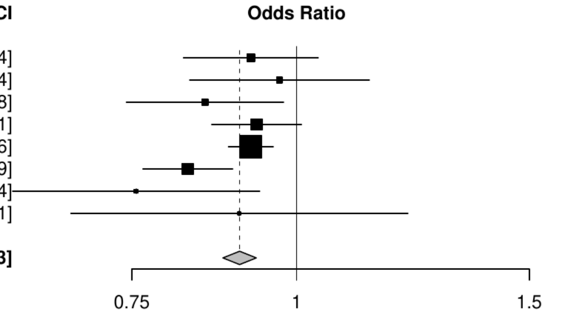


**Supplementary figure 11. Forest plot for rs2985994 (C), COG3**

| Study | OR | 95%–CI |
|---|---|---|
| s1 | 1.12 | [1.03; 1.21] |
| s2 | 1.07 | [0.96; 1.20] |
| s3 | 1.05 | [0.95; 1.17] |
| s4 | 1.06 | [0.99; 1.14] |
| s6 | 1.06 | [1.03; 1.10] |
| s7 | 1.07 | [1.01; 1.14] |
| China | 1.07 | [0.96; 1.19] |
| Japan | 1.15 | [1.02; 1.30] |
| **Fixed effect model** | **1.07** | **[1.05; 1.10]** |

Heterogeneity: $I^2 = 0\%$, $\chi^2_7 = 2.81$

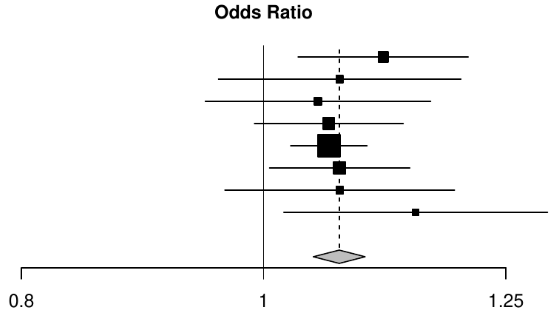

**Supplementary figure 12. Forest plot for rs229195 (A), *SCFD1***

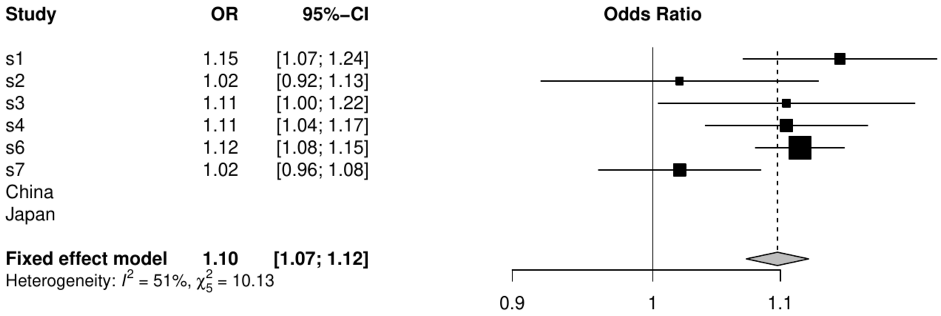


**Supplementary figure 13. Forest plot for rs12608932 (C), *UNC13A***

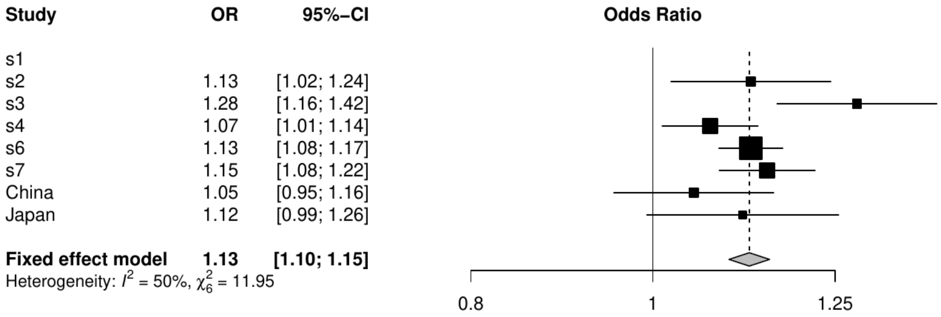


**Supplementary figure 14. Forest plot for rs17785991 (A), *SLC9A8/SPATA2***

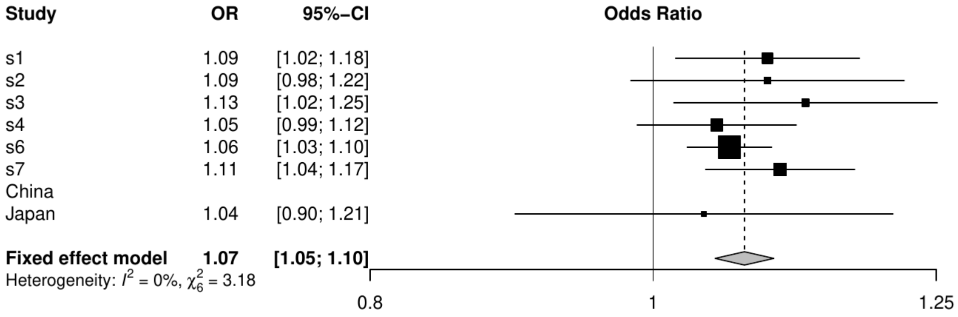

#### Supplementary figure 15. Forest plot for rs80265967 (C), *SOD1*

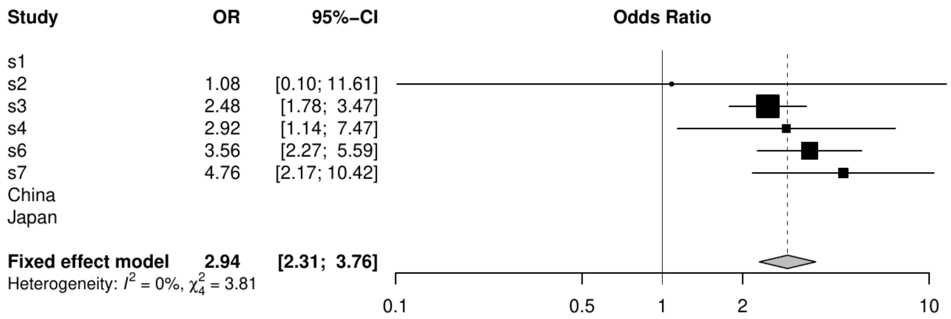


#### Supplementary figure 16. Forest plot for rs75087725 (A), *CFAP410*

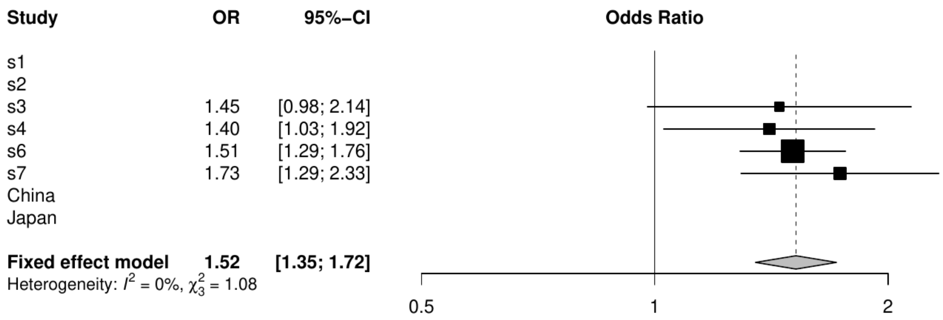


#### Supplementary figure 17. Forest plot for rs58854276 (A), *ACSL5/ZDHHC6*

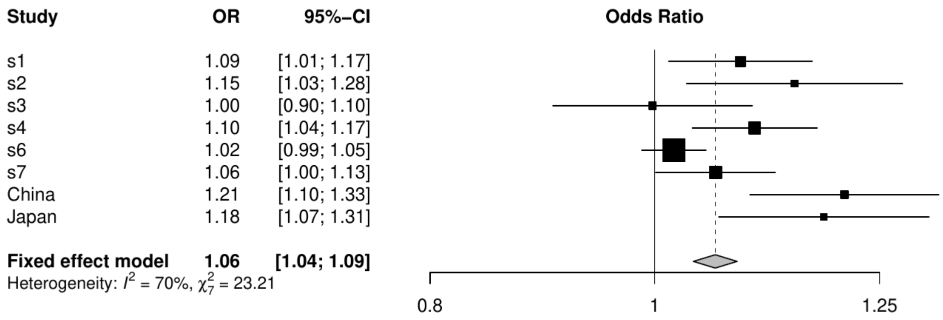

**Supplementary figure 18: Annotation specific enrichment.** Enrichment of SNP-based heritability was calculated with LD-score regression. Grey dashed line represents no enrichment (enrichment = 1). Error bars denote standard error of enrichment estimate. Due to the regression framework in LDSC, enrichment estimates can be < 0 (with large standard errors).

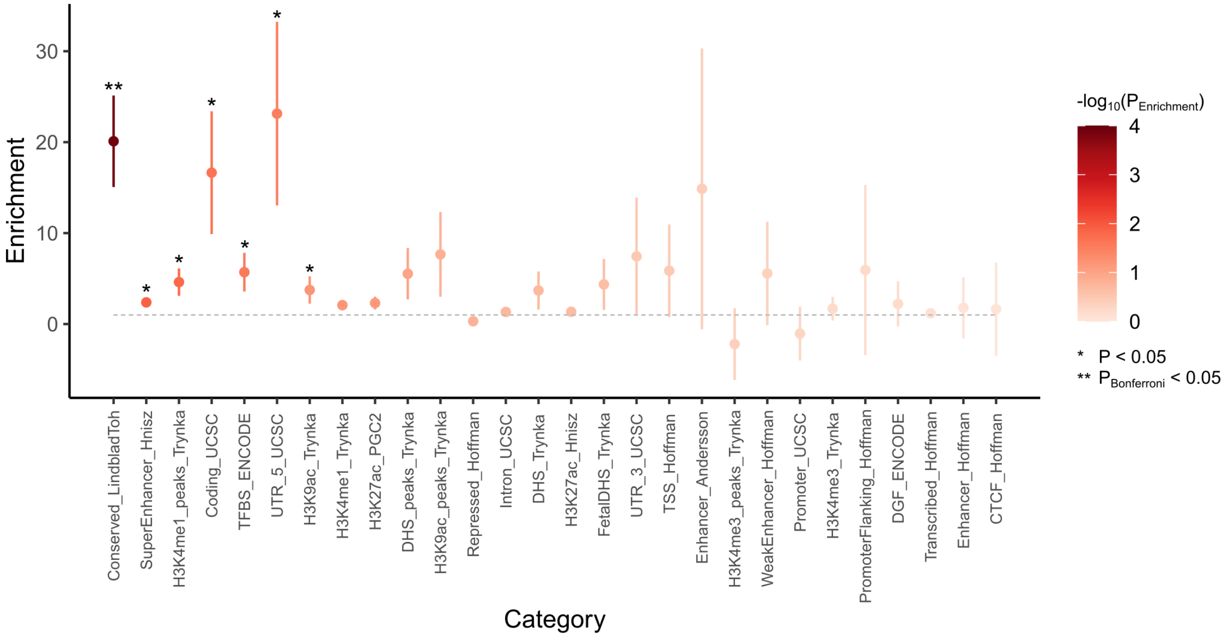


**Supplementary figures 19-32. Rare variant burden analyses.** Burden analysis of each protein coding transcript was performed using Firth logistic regression. Various SNVs sets were used according to MAF and functional consequence. On the left, Manhattan plots of each transcript are shown. Dotted line is the Bonferroni threshold correction for the number of genes tested (0.05/54849). On the right, qq-plots are shown. Shaded areas are 95% confidence intervals of expected $-log_{10}(P)$ under the null hypothesis of no association.

**Supplementary figure 19. Model 1.** MAF < 0.01, disruptive. $\lambda_{GC}$ = 0.944.

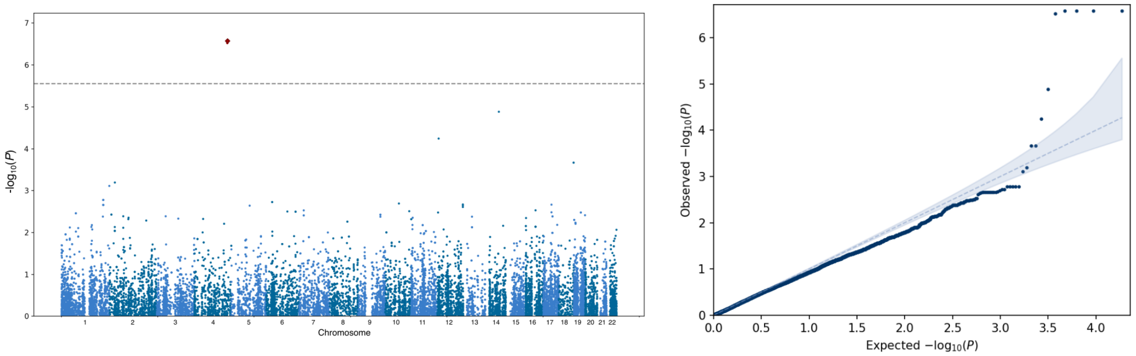

**Supplementary figure 20. Model 2.** MAF < 0.005, disruptive. $\lambda_{GC}$ = 0.940.

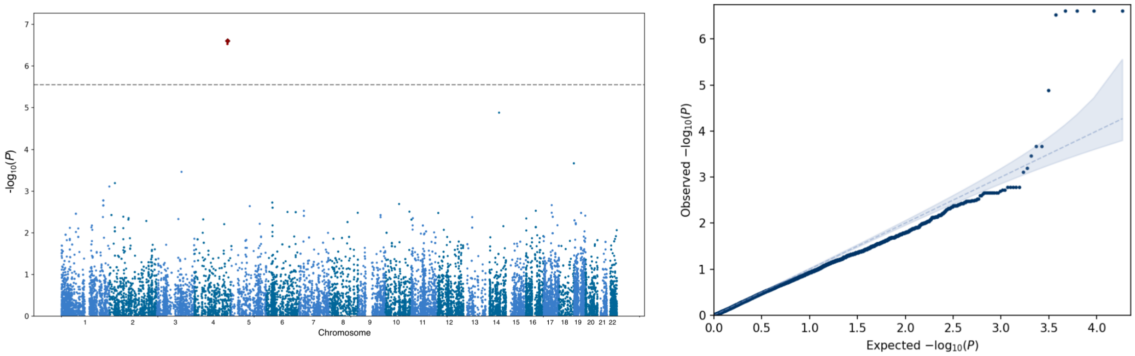


**Supplementary figure 21. Model 3.** MAF < 0.01, damaging. $\lambda_{GC}$ = 0.942.

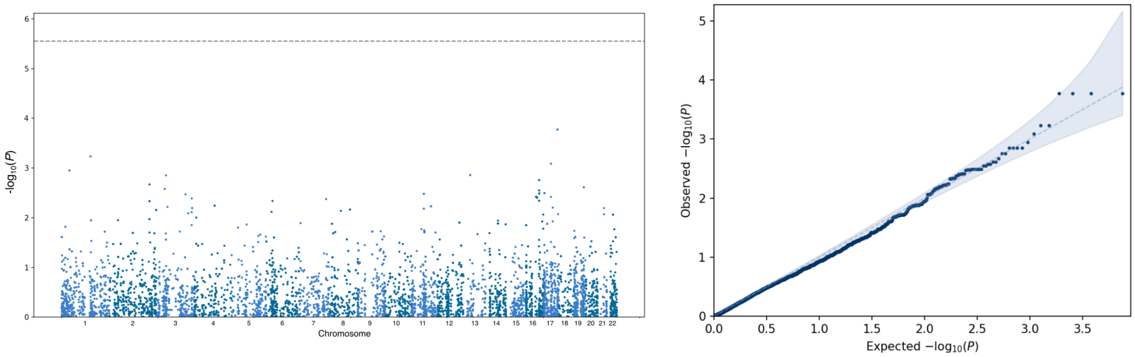


**Supplementary figure 22. Model 4.** MAF < 0.005, damaging. $\lambda_{GC}$ = 0.938.

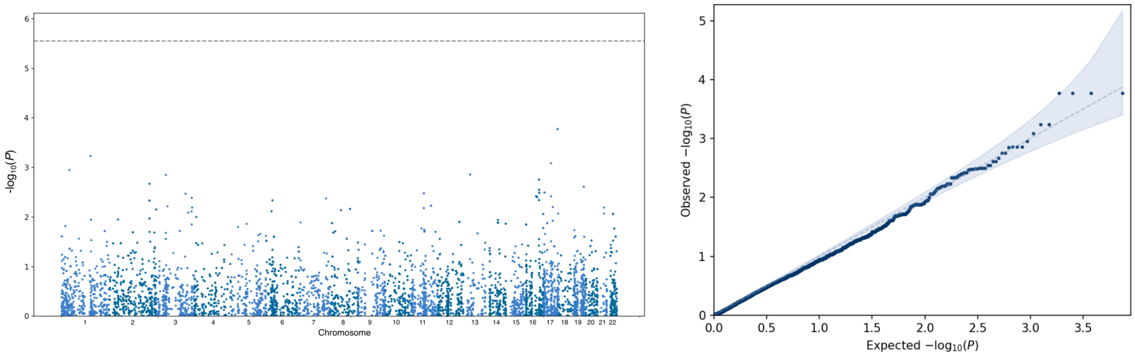

**Supplementary figure 23. Model 5.** MAF < 0.01, missense. $\lambda_{GC}$ = 0.998.

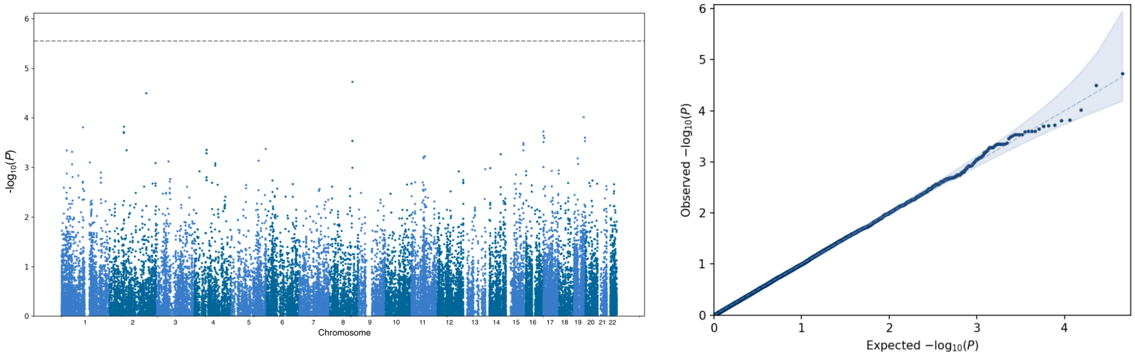


**Supplementary figure 24. Model 6.** MAF < 0.005, missense. $\lambda_{GC}$ = 0.992.

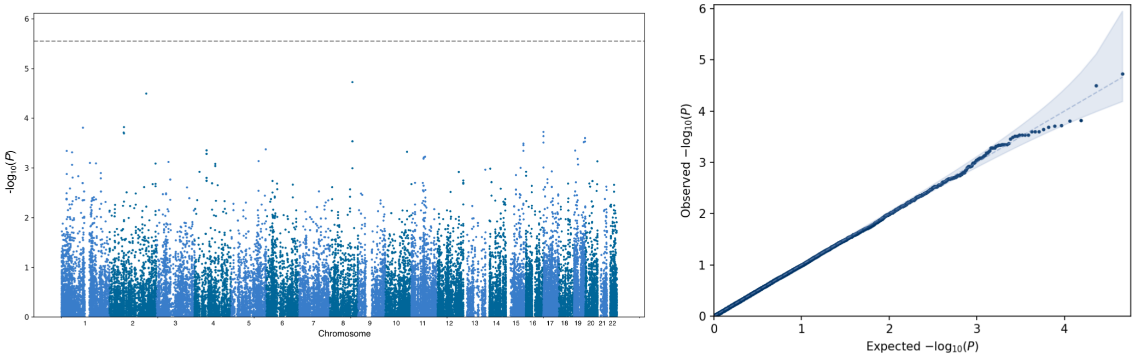


**Supplementary figure 25. Model 7.** MAF < 0.01, synonymous. $\lambda_{GC}$ = 0.977.

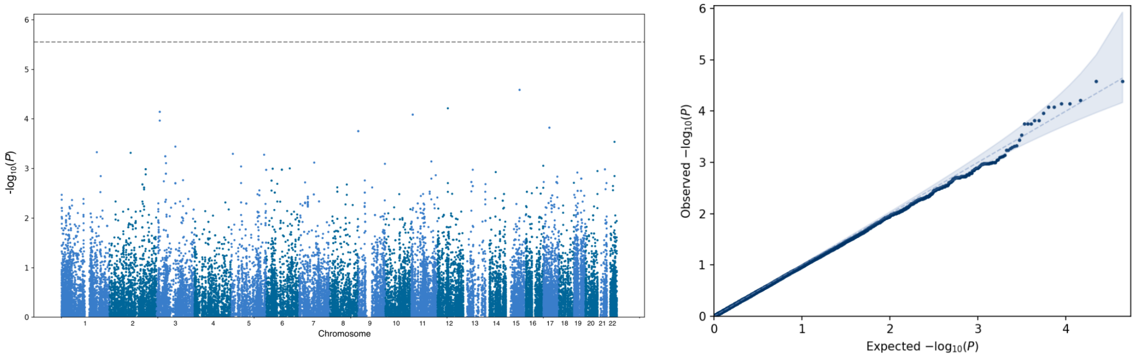

**Supplementary figure 26. Model 8.** MAF < 0.005, synonymous. $\lambda_{GC}$ = 0.989.

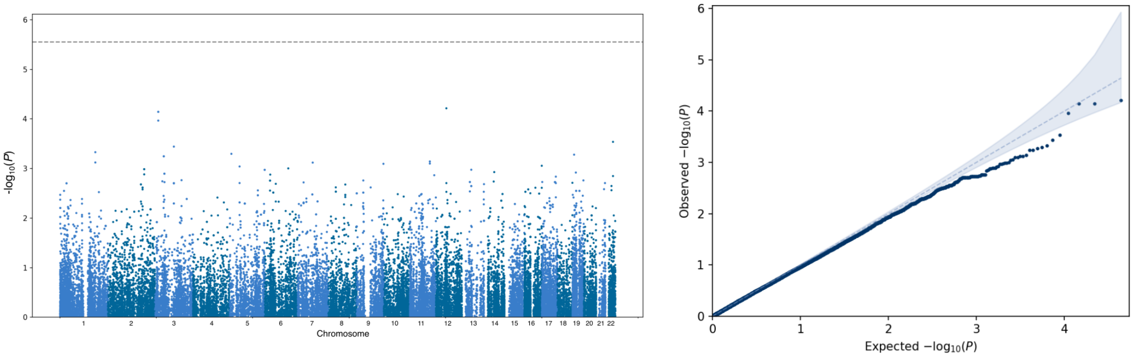


**Supplementary figure 27. Model 9.** MAF < 0.01, non-classified. $\lambda_{GC}$ = 0.976.

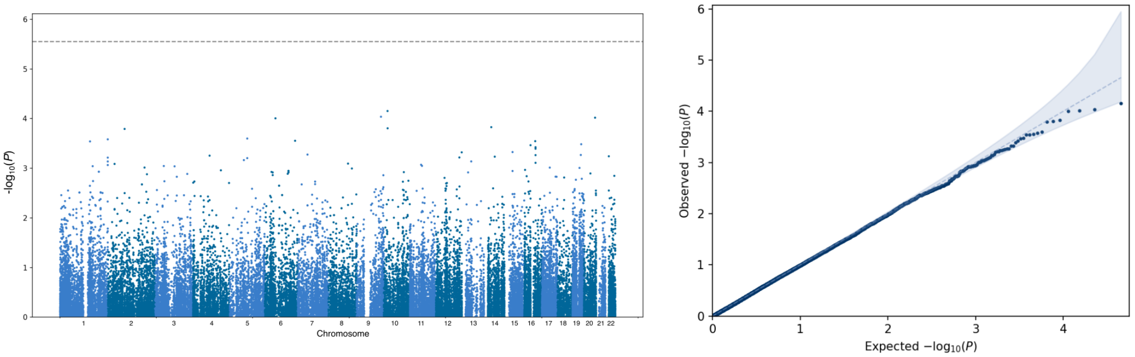


**Supplementary figure 28. Model 10.** MAF < 0.005, non-classified. $\lambda_{GC}$ = 0.976.

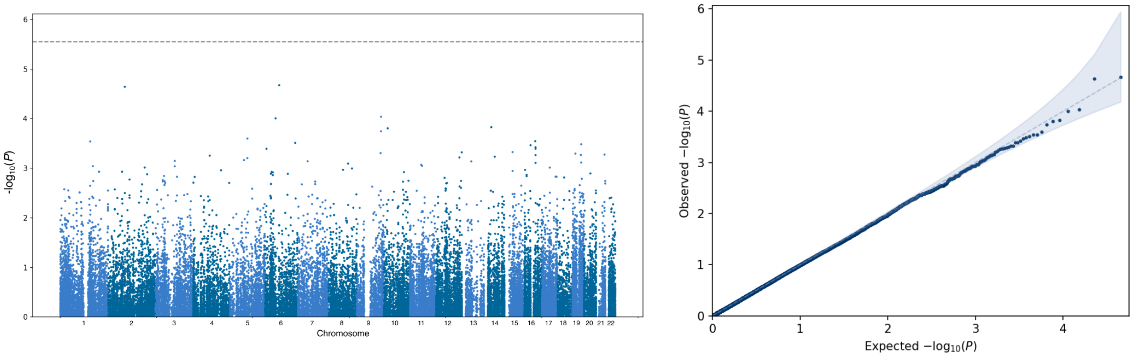

**Supplementary figure 29. Model 11.** MAF < 0.01, disruptive + damaging. $\lambda_{GC}$ = 0.959.

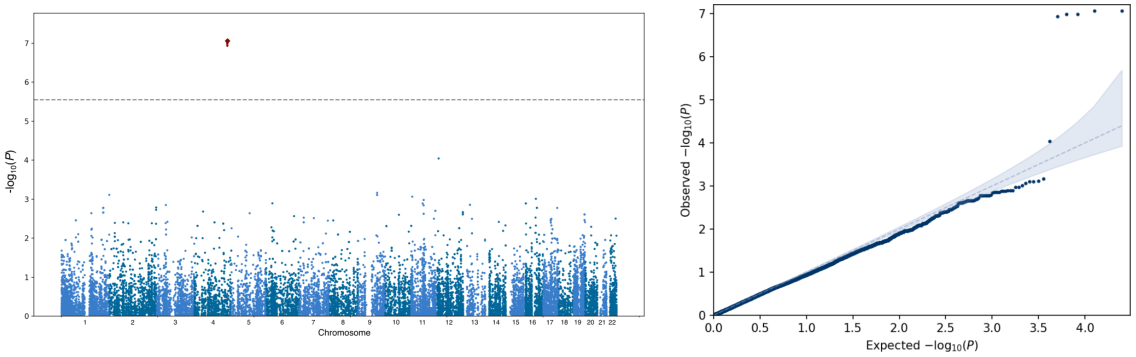


**Supplementary figure 30. Model 12.** MAF < 0.005, disruptive + damaging. $\lambda_{GC}$ = 0.945.

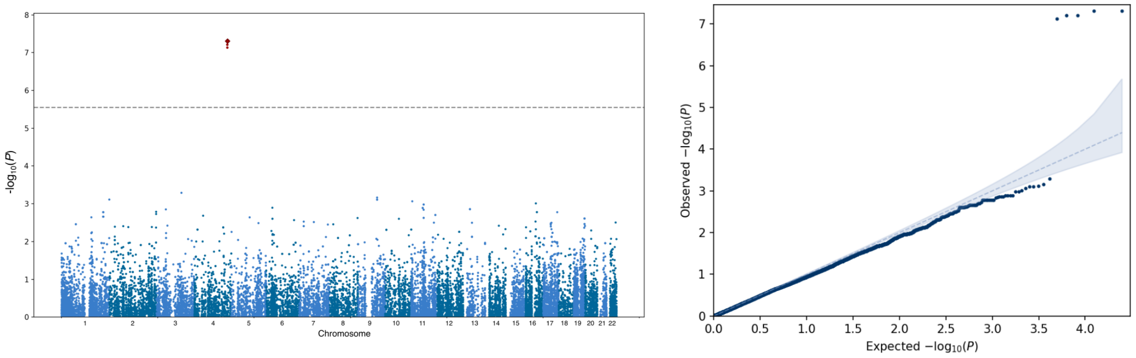


**Supplementary figure 31. Model 13.** MAF < 0.01, disruptive + damaging + missense. $\lambda_{GC}$ = 0.993.

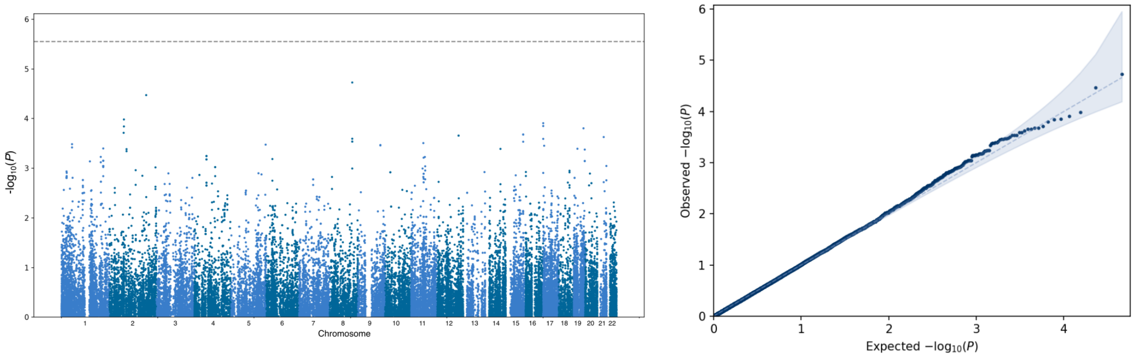

**Supplementary figure 32. Model 14.** MAF < 0.005, disruptive + damaging + missense. $\lambda_{GC}$ = 1.000.

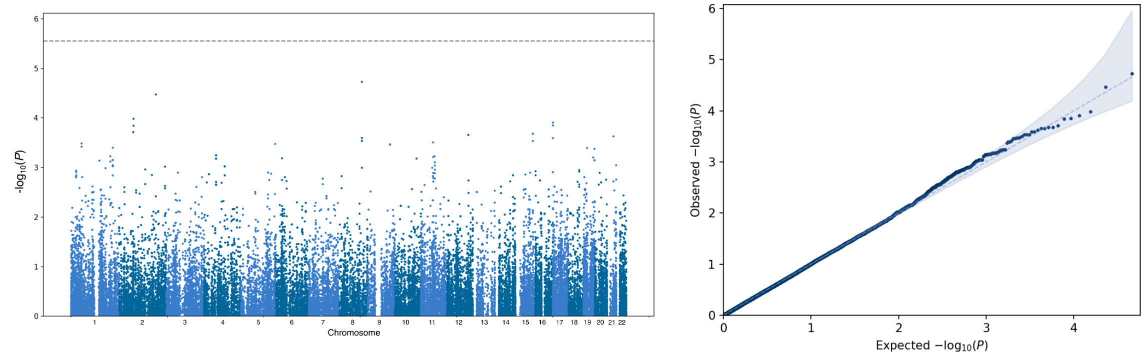


**Supplementary Figures 33-. Track plots for gene prioritization.** Overview of genome-wide significant loci. The locus boundaries are defined as distance to the furthest SNPs with LD $r^2 \geq 0.2$ according to Central European samples from 1000 Genomes phase 3, or 250 Kb on both sides, whichever is greater. The track plots display the following data from top to bottom: 1) karyogram of the chromosome and chromosomal position of the locus. 2) ALS GWAS association statistics of the European + Asian ancestries meta-analysis. Colors illustrate LD with the lead SNP, as calculated in stratum 4. Dotted line is the genome-wide significance threshold of $P = 5 \times 10^{-8}$. 3) GWAS association statistics of other neurodegenerative diseases Alzheimer's disease (AD)[16], corticobasal degeneration (CBD)[17], frontotemporal dementia (FTD)[18], Parkinson's disease (PD)[19] and progressive supranuclear palsy (PSP)[20]. Dotted line: $P = 5 \times 10^{-8}$. 4) Association statistics obtained by Firth logistic regression of repeat size called by ExpansionHunter v4 (ref [21]) or ExpansionHunter Denovo[22]. Dotted line is the Bonferroni multiple testing threshold for the number of repeats tested in the locus. 5) Summary-based Mendelian Randomization (SMR)[23] P-value of methylation quantitative trait loci (mQTL). mQTL effects were obtained from brain tissue and blood. Dotted line is the Bonferroni threshold of number of mQTL effects (brain and blood) tested in the locus. 5) Genetrack obtained from the UCSC browser. 6) Rare variant gene-based burden analysis P-value. Dotted line is the Bonferroni threshold for the number of genes tested within the locus. 7) eQTL-based SMR p-value. eQTLs obtained from blood (eQTLGen) and brain (metaBrain Cortex) were used. Dotted line is the Bonferroni threshold for the number of genes times the number of tissues these genes were tested in. Genes depicted in the shaded area were available in the eQTL dataset, but no eQTLs were strong enough to be included in the SMR analysis. 8) Polygenic priority score (PoPS)[24] quantiles. Quantiles are defined on scores from genome-wide PoPS scores.

**Supplementary figure 33. Trackplot rs631312, *MOBP/RPSA.*** Within this locus no genes were enriched for rare variants that contributed to ALS risk and no repeat expansions associated to ALS risk were found. The ALS-associated SNPs within the rs631312 are predicted to mediate risk of ALS through expression of *CCR8*, *RPSA*, and *MOBP* in brain tissues. Additionally, the three significant CpG sites identified through SMR are predicted to regulate expression of *RPSA* and *MOBP* and *SNORA6A2*. Of *RPSA* and *MOBP* prioritized by both methods, *RPSA* has most gene features in common with the other genes in genome-wide significant loci.

[figure on next page]

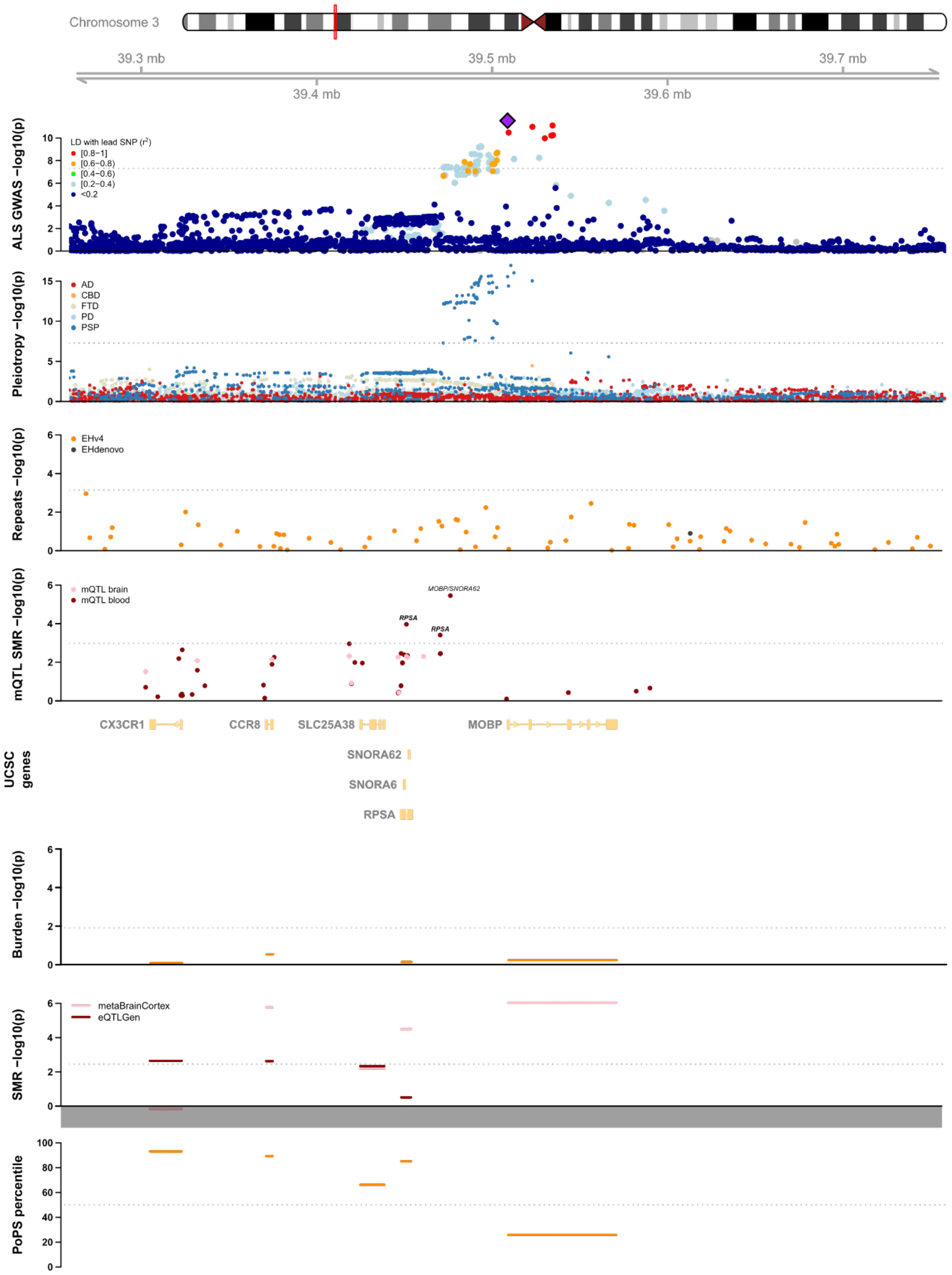
Chromosome 3
39.3 mb
39.5 mb
39.7 mb
39.4 mb
39.6 mb
ALS GWAS −log10(p)
LD with lead SNP (r2)
[0.8−1]
[0.6−0.8)
[0.4−0.6)
[0.2−0.4)
<0.2
Pleiotropy −log10(p)
AD
CBD
FTD
PD
PSP
Repeats −log10(p)
EHv4
EHdenovo
mQTL SMR −log10(p)
mQTL brain
mQTL blood
MOBP/SNORA62
RPSA
RPSA
CX3CR1
CCR8
SLC25A38
MOBP
SNORA62
SNORA6
RPSA
UCSC genes
Burden −log10(p)
SMR −log10(p)
metaBrainCortex
eQTLGen
PoPS percentile

**Supplementary figure 34. Trackplot for rs62333164, *NEK1*.** *NEK1* is a known ALS gene in this locus[25] and we find rare variants in *NEK1* to increase ALS risk. This rare variant burden signal is conditionally independent from the common variant signal ($P_{RVB} = 2.53 \times 10^{-7}$, $P_{RVB|SNP} = 2.32 \times 10^{-7}$). Additionally, an expansion of the tetranucleotide repeat chr4:17030384[TTTA]$_n$ downstream of *NEK1* with more than 10 repeat units is associated with ALS risk (expanded allele-frequency = 0.51, $P_{Repeat} = 5.2 \times 10^{-5}$, FDR = $4.7 \times 10^{-4}$). This repeat expansion is tagged by the top associated GWAS SNP within this locus and therefore does not represent a fully independent signal ($P_{Repeat|SNP} = 0.003$, $r^2 = 0.24$, $|D'| = 0.70$). Besides the rare variant burden signal and repeat expansion, SMR prioritized *NEK1* as the causal gene through both eQTL and mQTL. For both the eQTL and mQTL signals the HEIDI pleiotropy test indicated nominal evidence for pleiotropy ($P < 0.05$, but $P > 0.05$/number of genes).

[figure on next page]

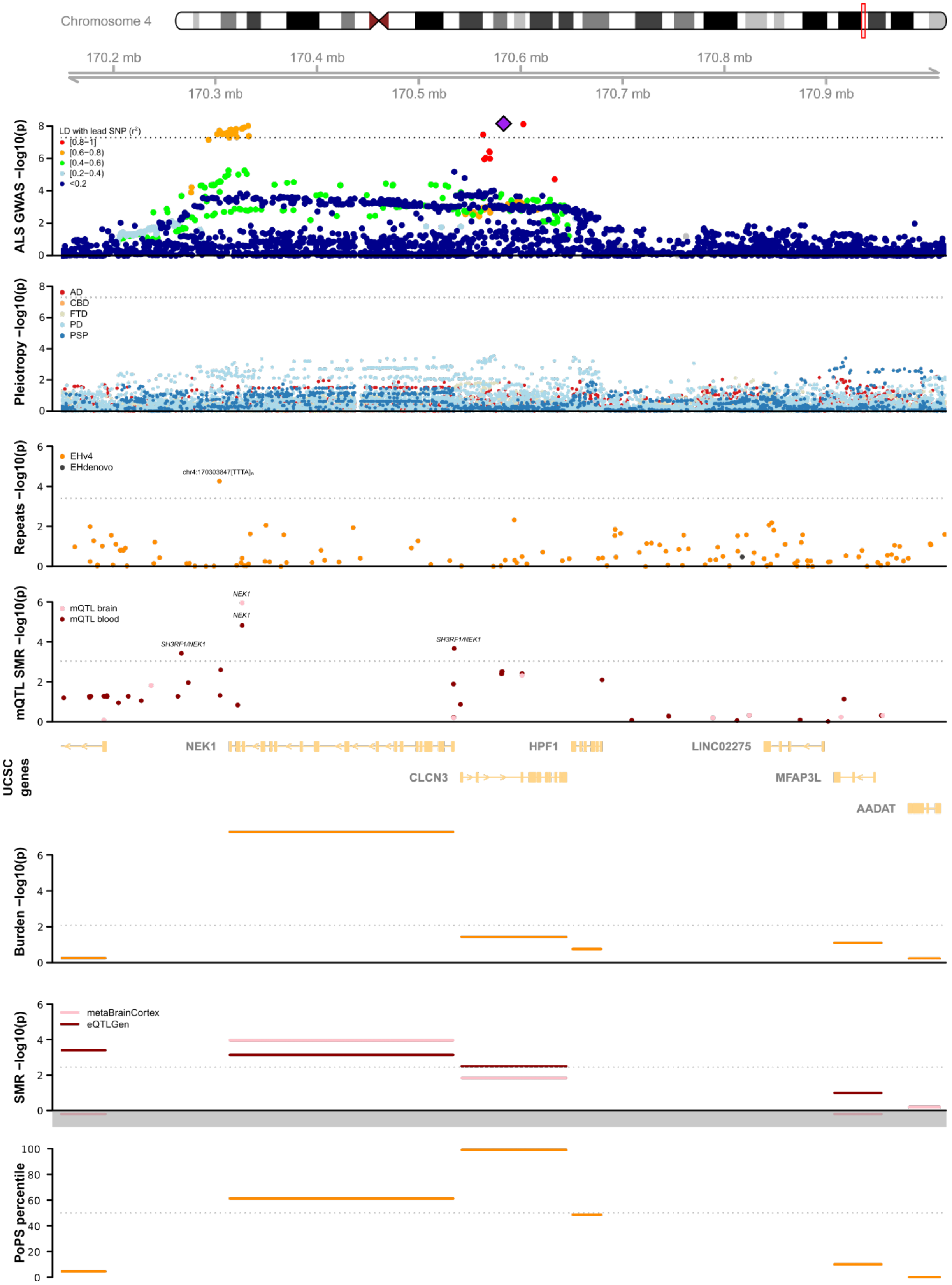

Chromosome 4
170.2 mb
170.4 mb
170.6 mb
170.8 mb
170.3 mb
170.5 mb
170.7 mb
170.9 mb
ALS GWAS −log10(p)
LD with lead SNP ($r^2$)
[0.8−1]
[0.6−0.8)
[0.4−0.6)
[0.2−0.4)
<0.2
Pleiotropy −log10(p)
AD
CBD
FTD
PD
PSP
Repeats −log10(p)
EHv4
EHdenovo
chr4:170303847[TTTA]n
mQTL SMR −log10(p)
mQTL brain
mQTL blood
NEK1
NEK1
SH3RF1/NEK1
SH3RF1/NEK1
UCSC genes
NEK1
HPF1
LINC02275
CLCN3
MFAP3L
AADAT
Burden −log10(p)
SMR −log10(p)
metaBrainCortex
eQTLGen
PoPS percentile

**Supplementary figure 35. Trackplot for rs7727034, *GPX3/TNIP1.*** In the rs7727034 locus, several mQTL effects were found which, based on annotation, were predicted to affect both *TNIP1* and *GPX3*, blood prioritized both *TNIP1* and *GPX3* as well. Brain-specific eQTL, however, prioritized *GPX3* over *TNIP1*. None of the genes within this locus harbored rare variants that were associated with ALS risk, neither where any of the repeats within this locus.
[figure on next page]

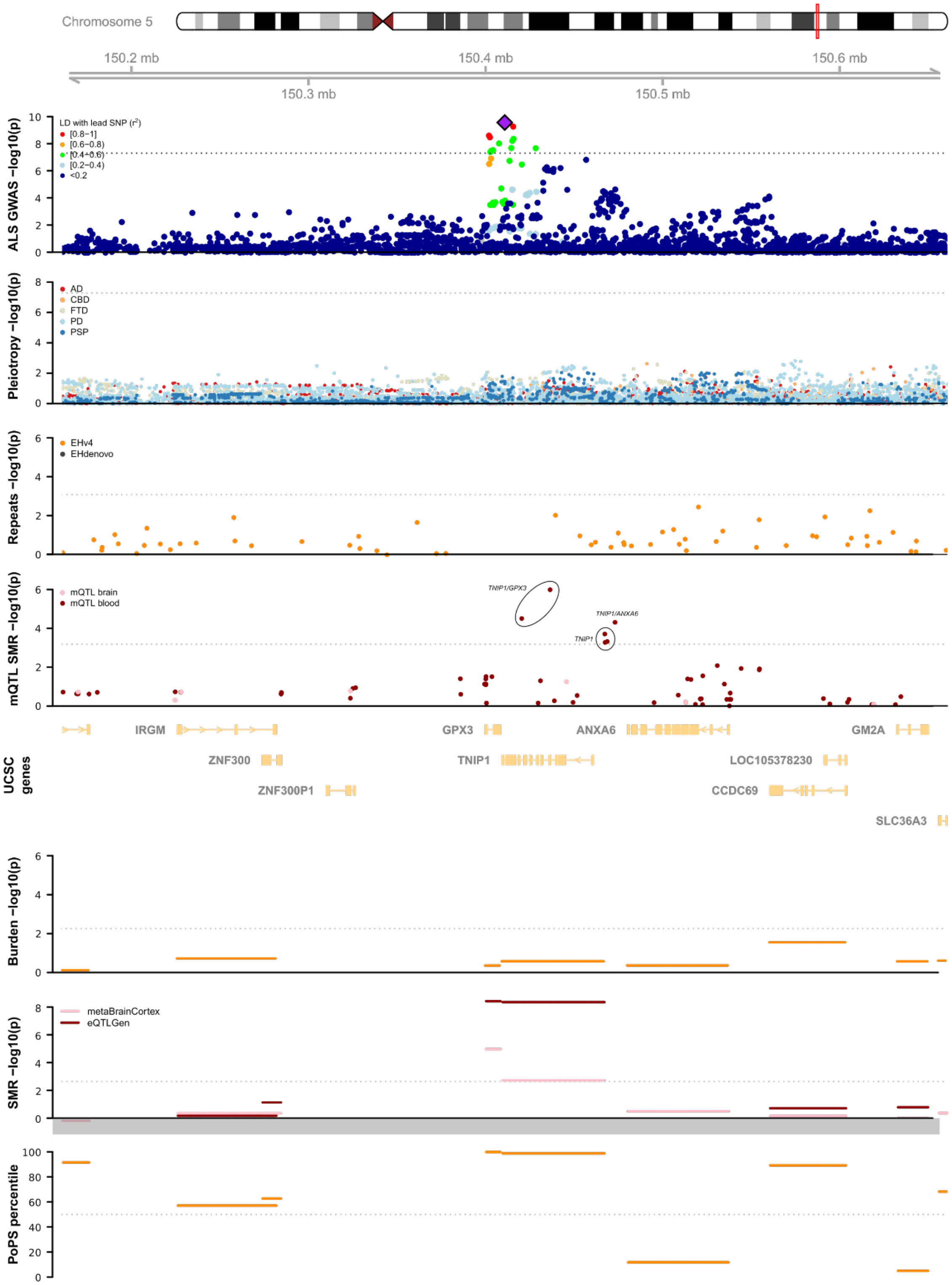
Chromosome 5
150.2 mb
150.4 mb
150.6 mb
150.3 mb
150.5 mb
ALS GWAS −log10(p)
LD with lead SNP (r²)
[0.8−1]
[0.6−0.8)
[0.4−0.6)
[0.2−0.4)
<0.2
Pleiotropy −log10(p)
AD
CBD
FTD
PD
PSP
Repeats −log10(p)
EHv4
EHdenovo
mQTL SMR −log10(p)
mQTL brain
mQTL blood
TNIP1/GPX3
TNIP1/ANXA6
TNIP1
UCSC genes
IRGM
GPX3
ANXA6
GM2A
ZNF300
TNIP1
LOC105378230
ZNF300P1
CCDC69
SLC36A3
Burden −log10(p)
SMR −log10(p)
metaBrainCortex
eQTLGen
PoPS percentile

**Supplementary figure 36. Trackplot for rs517339, *ERGIC1.*** Within the rs517339 locus we found no genes where rare variants were associated with an increased risk of ALS. The mQTL analysis, however, prioritized *ERGIC1* and eQTL analysis nominates *ERGIC1, CREBRF* and *SNIP1*. Finally, the gene feature prioritization nominates *ATP6V0E1.* No ALS-associated repeats were found.
[figure on next page]

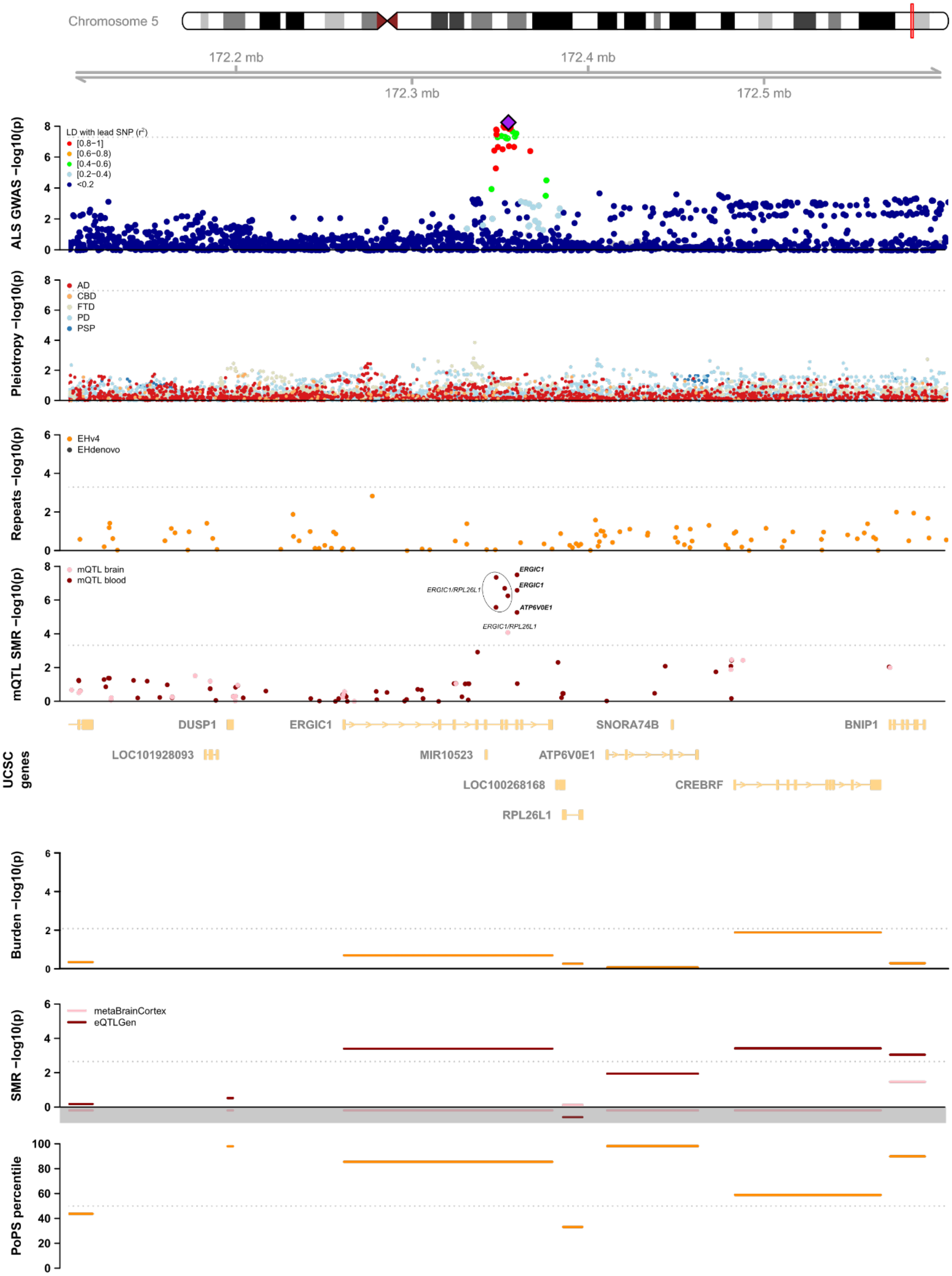
Chromosome 5
172.2 mb
172.3 mb
172.4 mb
172.5 mb
ALS GWAS −log10(p)
LD with lead SNP (r²)
[0.8−1]
[0.6−0.8)
[0.4−0.6)
[0.2−0.4)
<0.2
Pleiotropy −log10(p)
AD
CBD
FTD
PD
PSP
Repeats −log10(p)
EHv4
EHdenovo
mQTL SMR −log10(p)
mQTL brain
mQTL blood
ERGIC1
ERGIC1
ERGIC1/RPL26L1
ATP6V0E1
ERGIC1/RPL26L1
UCSC genes
DUSP1
ERGIC1
SNORA74B
BNIP1
LOC101928093
MIR10523
ATP6V0E1
LOC100268168
CREBRF
RPL26L1
Burden −log10(p)
SMR −log10(p)
metaBrainCortex
eQTLGen
PoPS percentile

**Supplementary figure 37. Trackplot for rs9275477, *HLA.*** The HLA locus, with rs9275477 as lead SNP contains many genes, and long-range LD. Neither rare variants, repeat expansions, nor regulatory effects through eQTL and mQTL prioritize a gene within this locus.
[figure on next page]

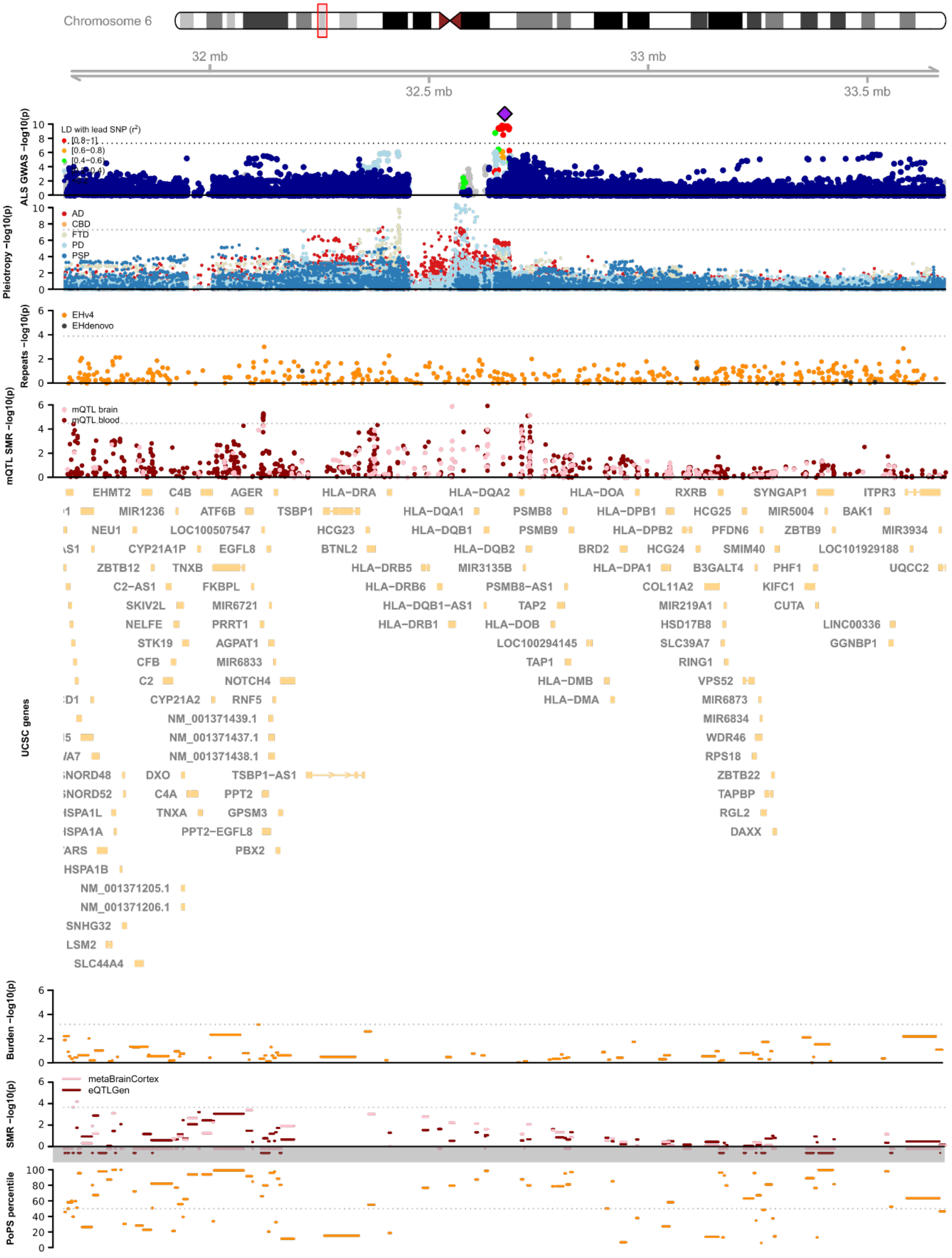

Chromosome 6
32 mb
33 mb
32.5 mb
33.5 mb
ALS GWAS −log10(p)
LD with lead SNP (r²)
[0.8−1]
[0.6−0.8)
[0.4−0.6)
Pleiotropy −log10(p)
AD
CBD
FTD
PD
PSP
Repeats −log10(p)
EHv4
EHdenovo
mQTL SMR −log10(p)
mQTL brain
mQTL blood
UCSC genes
EHMT2
C4B
AGER
HLA−DRA
HLA−DQA2
HLA−DOA
RXRB
SYNGAP1
ITPR3
MIR1236
ATF6B
TSBP1
HLA−DQA1
PSMB8
HLA−DPB1
HCG25
MIR5004
BAK1
NEU1
LOC100507547
HCG23
HLA−DQB1
PSMB9
HLA−DPB2
PFDN6
ZBTB9
MIR3934
CYP21A1P
EGFL8
BTNL2
HLA−DQB2
BRD2
HCG24
SMIM40
LOC101929188
ZBTB12
TNXB
HLA−DRB5
MIR3135B
HLA−DPA1
B3GALT4
PHF1
UQCC2
C2−AS1
FKBPL
HLA−DRB6
PSMB8−AS1
COL11A2
KIFC1
SKIV2L
MIR6721
HLA−DQB1−AS1
TAP2
MIR219A1
CUTA
NELFE
PRRT1
HLA−DRB1
HLA−DOB
HSD17B8
LINC00336
STK19
AGPAT1
LOC100294145
SLC39A7
GGNBP1
CFB
MIR6833
TAP1
RING1
C2
NOTCH4
HLA−DMB
VPS52
CYP21A2
RNF5
HLA−DMA
MIR6873
NM_001371439.1
MIR6834
NM_001371437.1
WDR46
NM_001371438.1
RPS18
DXO
TSBP1−AS1
ZBTB22
C4A
PPT2
TAPBP
TNXA
GPSM3
RGL2
PPT2−EGFL8
DAXX
PBX2
HSPA1B
NM_001371205.1
NM_001371206.1
SNHG32
LSM2
SLC44A4
Burden −log10(p)
SMR −log10(p)
metaBrainCortex
eQTLGen
PoPS percentile

**Supplementary figure 38. Trackplot for rs10280711, *PTPRN2.*** The rs10280711 only contains a single protein-coding gene, *PTPRN2*. Fort *PTPRN2* we observe a significant eQTL effect Evidence is only available for *PTPRN2*, which has a significant eQTL SMR effect in blood. The rare variant burden for missense variants at MAF < 0.005 was nomainally significant ($P_{RVB}$ = 0.038). Previous biomarker screens in FTD and AD have identified lower levels of PTPRN2 in cerebrospinal fluid of patients with FTD and *GRN* mutations and AD. An epigenome-wide methylation study in Parkinson's disease identified hypomethylation of CpG sites in *PTPRN2* as a marker for motor decline.

[figure on next page]

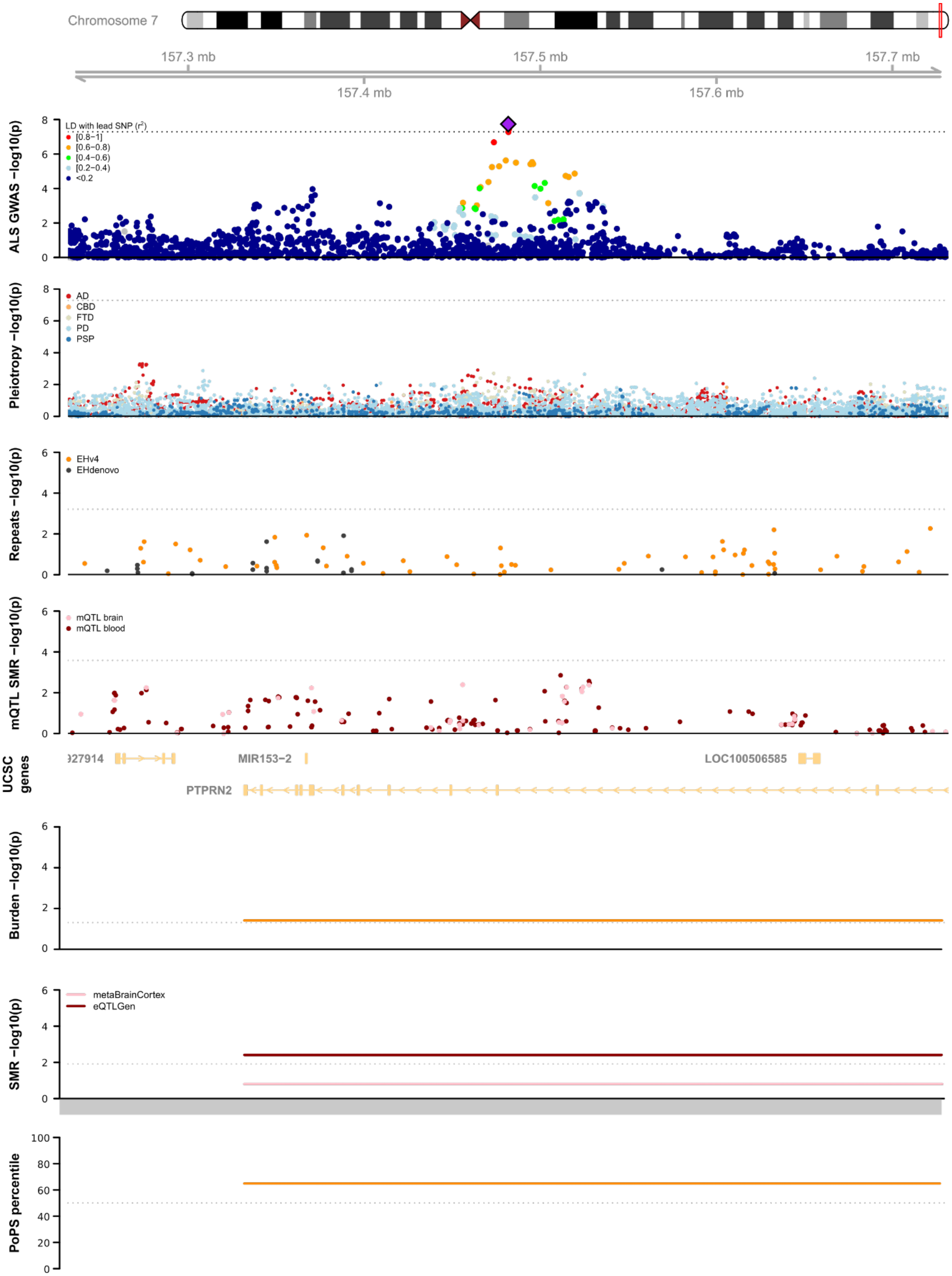
Chromosome 7
157.3 mb
157.5 mb
157.7 mb
157.4 mb
157.6 mb
ALS GWAS −log10(p)
LD with lead SNP (r²)
[0.8−1]
[0.6−0.8)
[0.4−0.6)
[0.2−0.4)
<0.2
Pleiotropy −log10(p)
AD
CBD
FTD
PD
PSP
Repeats −log10(p)
EHv4
EHdenovo
mQTL SMR −log10(p)
mQTL brain
mQTL blood
UCSC genes
MIR153−2
LOC100506585
PTPRN2
Burden −log10(p)
SMR −log10(p)
metaBrainCortex
eQTLGen
PoPS percentile

**Supplementary figure 39. Trackplot for rs2453555, *C9orf72*.** The rs2453555 SNP tags the pathogenic hexanucleotide repeat expansion in *C9orf72* which is the most common cause of ALS with or without frontotemporal dementia. This repeat expansion is detected by ExpansionHunter v4 since it is in the used catalogue, but found through de-novo assembly of unmapped reads with ExpansionHunter De-novo. The repeat expansion is in high LD with the top SNP ($r^2$ = 0.14, |D'| = 0.99) and after conditioning on the repeat expansions, there is no residual association signal within the whole-genome sequencing cohort for the top SNP ($P_{SNP}$ = 0.005, $P_{SNP|Repeat}$ = 0.86). Additionally, eQTL SMR analysis highlighted a potential effect of *C9orf72* gene expression, but the HEIDI test indicated strong evidence for heterogeneity ($P_{HEIDI} = 3.72 \times 10^{-23}$) suggesting horizontal pleiotropy and that the causal effect is not mediated through the SNP eQTL effect. An identical pattern was seen for mQTL SMR (minimal $P_{HEIDI} = 4.1 \times 10^{-7}$).
[figure on next page]

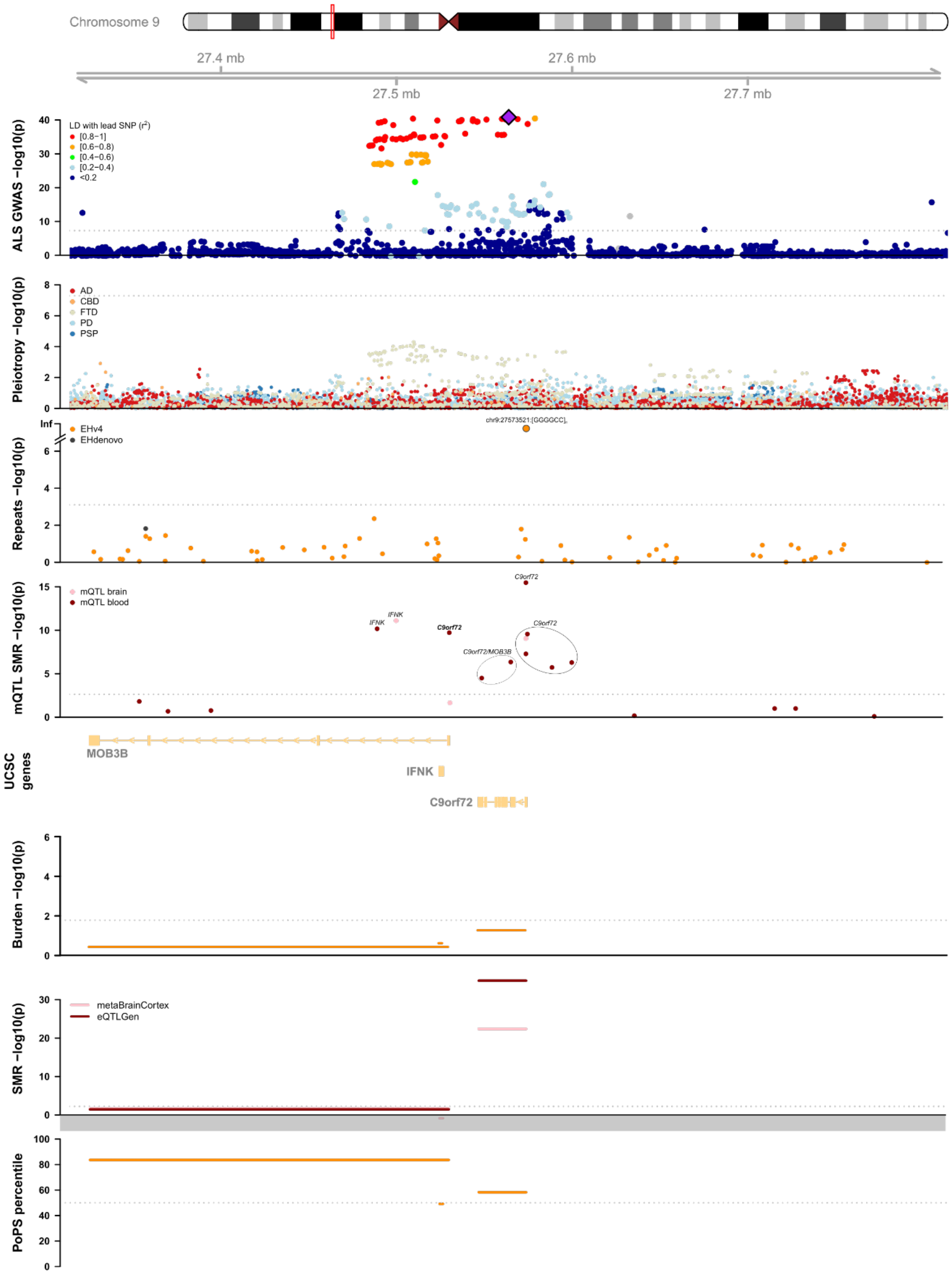

Chromosome 9
27.4 mb
27.6 mb
27.5 mb
27.7 mb
ALS GWAS −log10(p)
LD with lead SNP (r2)
[0.8−1]
[0.6−0.8)
[0.4−0.6)
[0.2−0.4)
<0.2
Pleiotropy −log10(p)
AD
CBD
FTD
PD
PSP
Repeats −log10(p)
Inf
EHv4
EHdenovo
chr9:27573521:[GGGGCC]
mQTL SMR −log10(p)
mQTL brain
mQTL blood
IFNK
IFNK
C9orf72
C9orf72
C9orf72
C9orf72/MOB3B
UCSC genes
MOB3B
IFNK
C9orf72
Burden −log10(p)
SMR −log10(p)
metaBrainCortex
eQTLGen
PoPS percentile

**Supplementary figure 40. Trackplot for rs113247976, *KIF5A.*** The rs113247976 SNP is a coding low-frequency variant in *KIF5A* (P986L, MAF = 0.0128 in HRC). There are few associated low-frequency SNPs in LD. Loss-of-function mutations in the cargo-binding tail domain of KIF5A have previously been identified in familial ALS, which together with the associated SNP identified KIF5A as ALS risk gene (Nicolas et al, Neuron 2019). We do not find an overall increased burden of rare variants in *KIF5A* in patients in our sequencing cohort, which consists of mostly apparently sporadic ALS patients. We find no evidence for repeat expansions or regulatory effects contributing to disease risk.

[figure on next page]

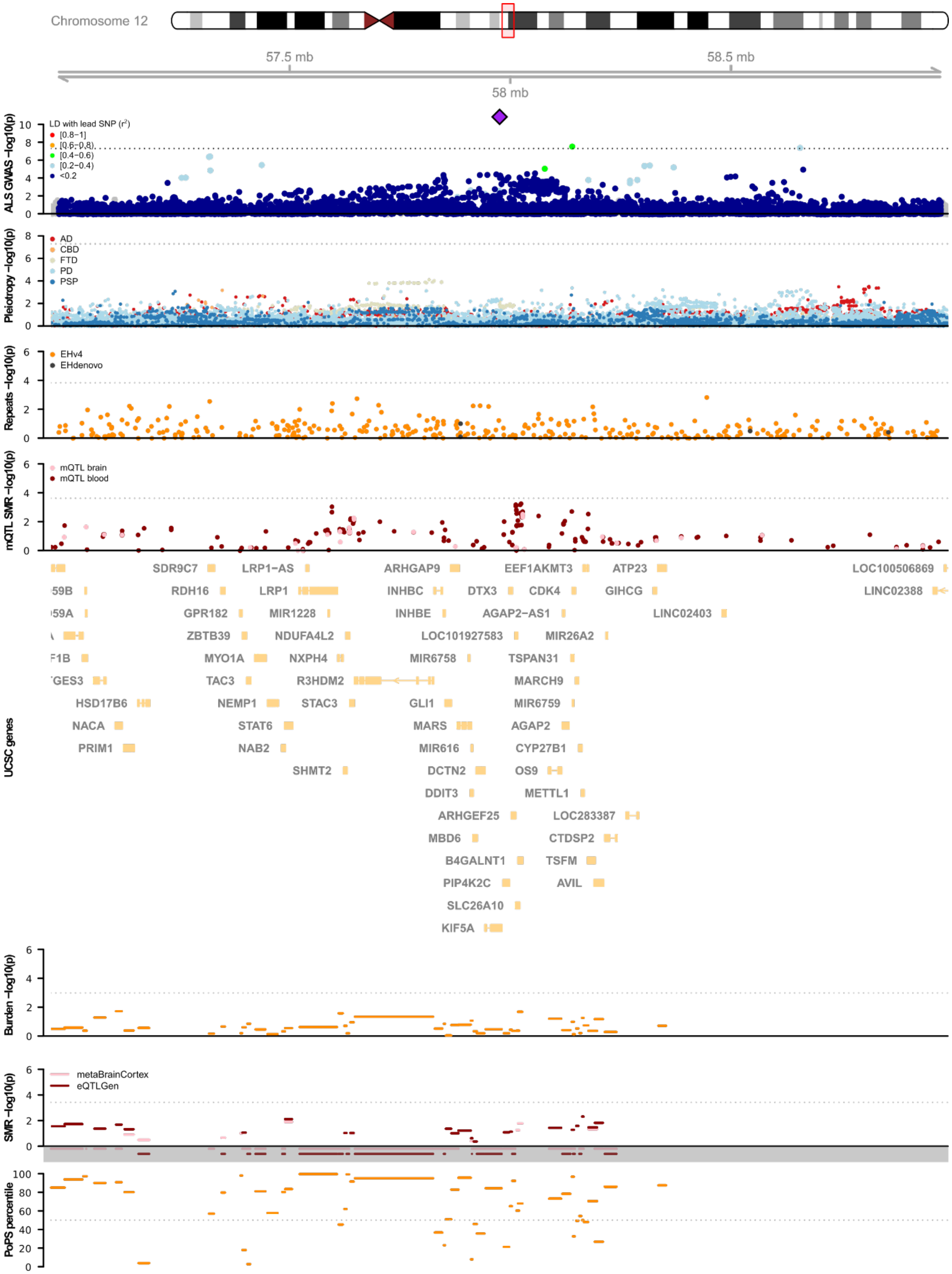

Chromosome 12
57.5 mb
58 mb
58.5 mb
ALS GWAS −log10(p)
LD with lead SNP (r²)
[0.8−1]
[0.6−0.8)
[0.4−0.6)
[0.2−0.4)
<0.2
Pleiotropy −log10(p)
AD
CBD
FTD
PD
PSP
Repeats −log10(p)
EHv4
EHdenovo
mQTL SMR −log10(p)
mQTL brain
mQTL blood
UCSC genes
SDR9C7
LRP1-AS
ARHGAP9
EEF1AKMT3
ATP23
LOC100506869
RDH16
LRP1
INHBC
DTX3
CDK4
GIHCG
LINC02388
GPR182
MIR1228
INHBE
AGAP2-AS1
LINC02403
ZBTB39
NDUFA4L2
LOC101927583
MIR26A2
MYO1A
NXPH4
MIR6758
TSPAN31
TAC3
R3HDM2
MARCH9
HSD17B6
NEMP1
STAC3
GLI1
MIR6759
NACA
STAT6
MARS
AGAP2
PRIM1
NAB2
MIR616
CYP27B1
SHMT2
DCTN2
OS9
DDIT3
METTL1
ARHGEF25
LOC283387
MBD6
CTDSP2
B4GALNT1
TSFM
PIP4K2C
AVIL
SLC26A10
KIF5A
Burden −log10(p)
SMR −log10(p)
metaBrainCortex
eQTLGen
PoPS percentile

**Supplementary figure 41. Trackplot for rs4075094, *TBK1*.** The *TBK1* gene is a known ALS gene within this locus. It was identified in a whole-exome sequencing case-control study in ALS and loss-of-function mutations are associated with familial ALS. Here, we find the rare variant burden for *TBK1* for missense mutations with MAF < 0.01 to be associated with ALS risk ($P_{RVB} = 5.4 \times 10^{-3}$) just passing the threshold for multiple testing across all genes within this locus ($0.05/8 = 6.25 \times 10^{-3}$). The conditional analysis in all individuals with whole-genome sequencing and GWAS data indicated that the rare-variant burden is conditionally independent from the GWAS signal ($P_{RVB} = 0.021$, $P_{RVB|SNP} = 0.021$). SMR identified CpG sites that were associated with *TBK1* expression suggesting that the regulatory effects on *TBK1* contribute to ALS risk independent from the known ALS associated rare variants. The HEIDI pleiotropy test showed nominal evidence for pleiotropy of the SMR signal (P < 0.05, but P > 0.05/number of genes).
[figure on next page]

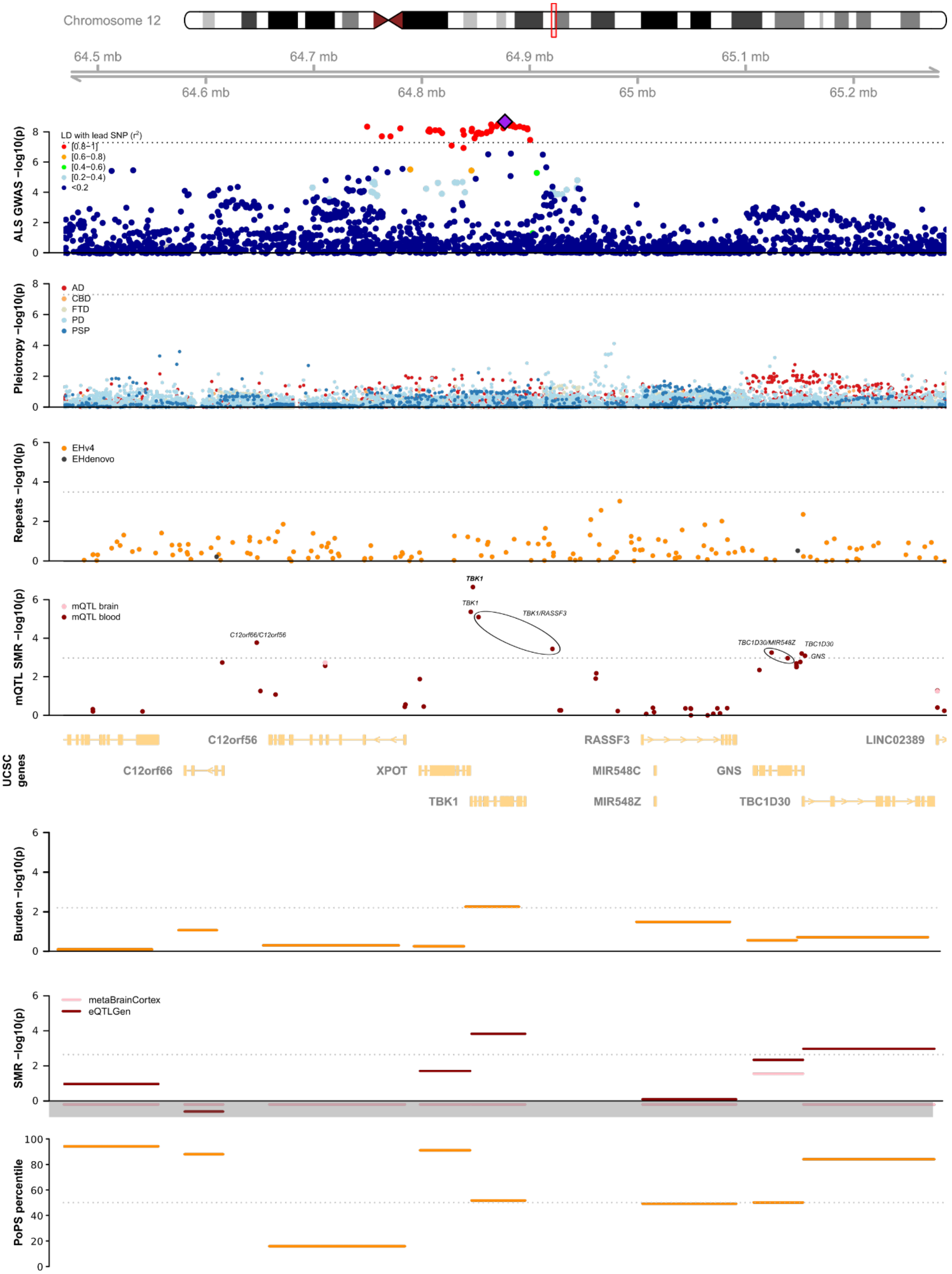

Chromosome 12
64.5 mb
64.7 mb
64.9 mb
65.1 mb
64.6 mb
64.8 mb
65 mb
65.2 mb
ALS GWAS −log10(p)
LD with lead SNP (r²)
[0.8−1]
[0.6−0.8)
[0.4−0.6)
[0.2−0.4)
<0.2
Pleiotropy −log10(p)
AD
CBD
FTD
PD
PSP
Repeats −log10(p)
EHv4
EHdenovo
mQTL SMR −log10(p)
mQTL brain
mQTL blood
TBK1
TBK1/RASSF3
C12orf66/C12orf56
TBC1D30/MIR548Z
TBC1D30
GNS
UCSC genes
C12orf56
RASSF3
LINC02389
C12orf66
XPOT
MIR548C
MIR548Z
TBC1D30
Burden −log10(p)
SMR −log10(p)
metaBrainCortex
eQTLGen
PoPS percentile

**Supplementary figure 42. Trackplot for rs2985994, *COG3*.** Within the rs2985994 locus, no genes exhibit a rare variant burden signal and we found no ALS-associated repeats. eQTL SMR in cortical tissues identified *COG3* as the most likely causal gene, with reduced expression leading to an increased risk of ALS ($b_{xy}$ = -0.239 ± 0.070 [s.e.], P = 5.15 × $10^{-4}$). No evidence for heterogeneity was found by the HEIDI test (P = 0.28). The two CpG site identified through SMR in blood methylation profiles were predicted to regulate *COG3* and/or *ERICH6B* and the gene feature selection analysis scored *COG3* (99.8 percentile) higher than *ERICH6B* (73.9 percentile). These analysis combined, prioritize *COG3* within the rs2985994 locus.

[figure on next page]

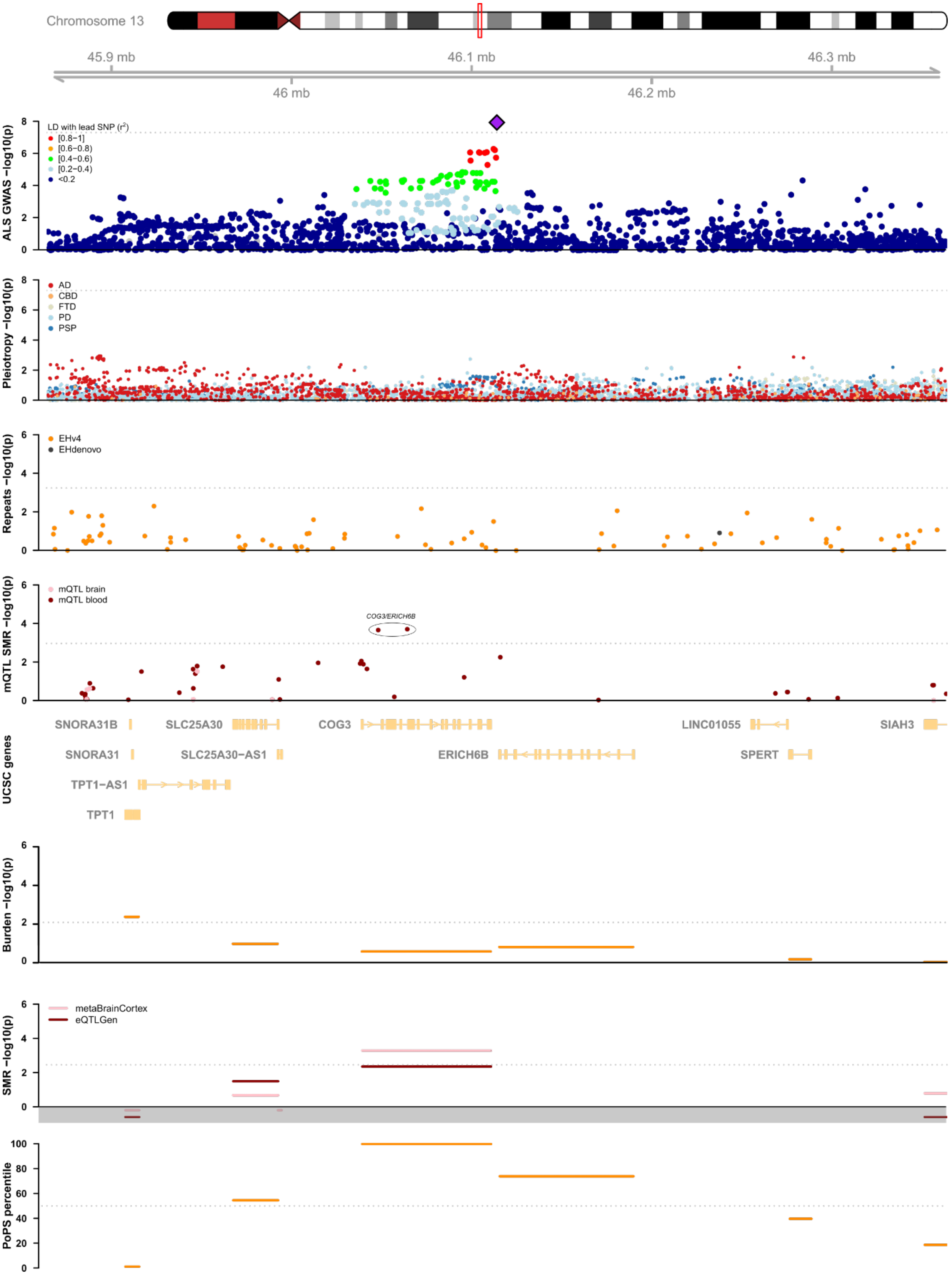
Chromosome 13
45.9 mb
46.1 mb
46.3 mb
46 mb
46.2 mb
ALS GWAS −log10(p)
LD with lead SNP (r²)
[0.8−1]
[0.6−0.8)
[0.4−0.6)
[0.2−0.4)
<0.2
Pleiotropy −log10(p)
AD
CBD
FTD
PD
PSP
Repeats −log10(p)
EHv4
EHdenovo
mQTL SMR −log10(p)
mQTL brain
mQTL blood
COG3/ERICH6B
UCSC genes
SNORA31B
SLC25A30
COG3
LINC01055
SIAH3
SNORA31
SLC25A30−AS1
ERICH6B
SPERT
TPT1−AS1
TPT1
Burden −log10(p)
SMR −log10(p)
metaBrainCortex
eQTLGen
PoPS percentile

**Supplementary figure 43. Trackplot for rs229243, *SCFD1.*** Within the rs229195 locus the associated SNPs span both *G2E3* and *SCFD1*. Rare variant burden analyses show no evidence for a rare variant signal in either two of these genes. There are no repeat expansions associated with ALS within this locus. mQTL SMR identified expression-altering CpG sites for both *G2E3* and *SCFD1*, with the strongest (brain-specific) signals for *SCFD1*. Similarly, the eQTL SMR signal for *SCFD1* is stronger than that for *G2E3* as well. Finally, gene feature analysis scores *SCFD1* (99.2 percentile) higher than *G2E3* (89.5 percentile). Taken together, these analyses prioritize *SCFD1* over *G2E3.*

[figure on next page]

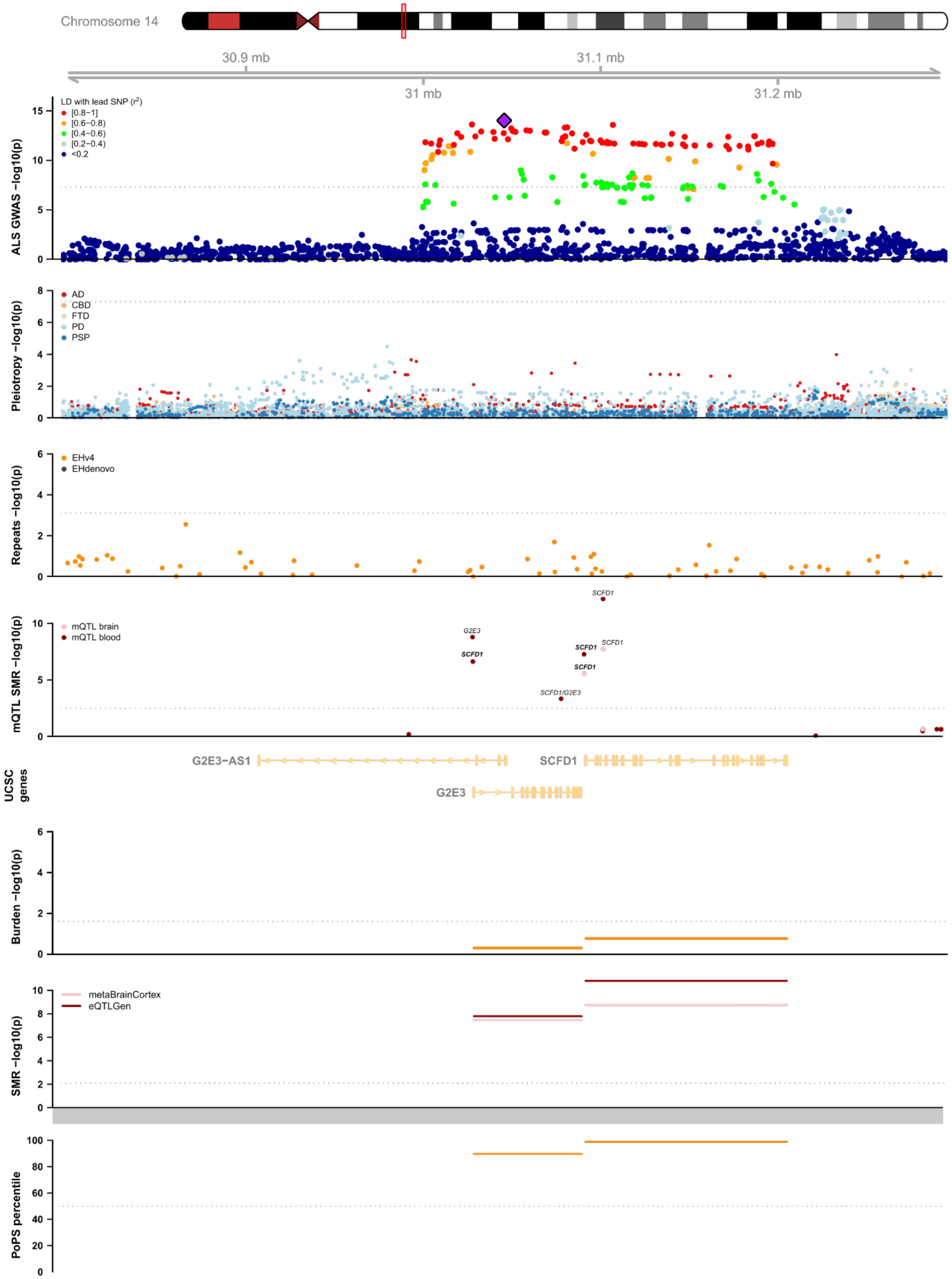
Chromosome 14
30.9 mb
31.1 mb
31 mb
31.2 mb
LD with lead SNP (r2)
[0.8–1]
[0.6–0.8)
[0.4–0.6)
[0.2–0.4)
<0.2
ALS GWAS −log10(p)
AD
CBD
FTD
PD
PSP
Pleiotropy −log10(p)
EHv4
EHdenovo
Repeats −log10(p)
mQTL brain
mQTL blood
mQTL SMR −log10(p)
SCFD1
G2E3
SCFD1/G2E3
UCSC genes
G2E3−AS1
G2E3
Burden −log10(p)
metaBrainCortex
eQTLGen
SMR −log10(p)
PoPS percentile

**Supplementary figure 44. Trackplot for rs12608932, *UNC13A*.** The rs12608932 locus was one of the first robustly associated GWAS loci in ALS together with the *C9orf72* signal[26] and has been identified in FTD as well[27]. There is little LD with the top SNP in this region and the associated SNPs are all intronic variants within the *UNC13A* gene. We found no evidence of ALS-associated rare variants or repeat expansions within this region. The ALS-associated SNPs do not act as strong eQTL or mQTL for any of the genes or CgG sites in this region.
[figure on next page]

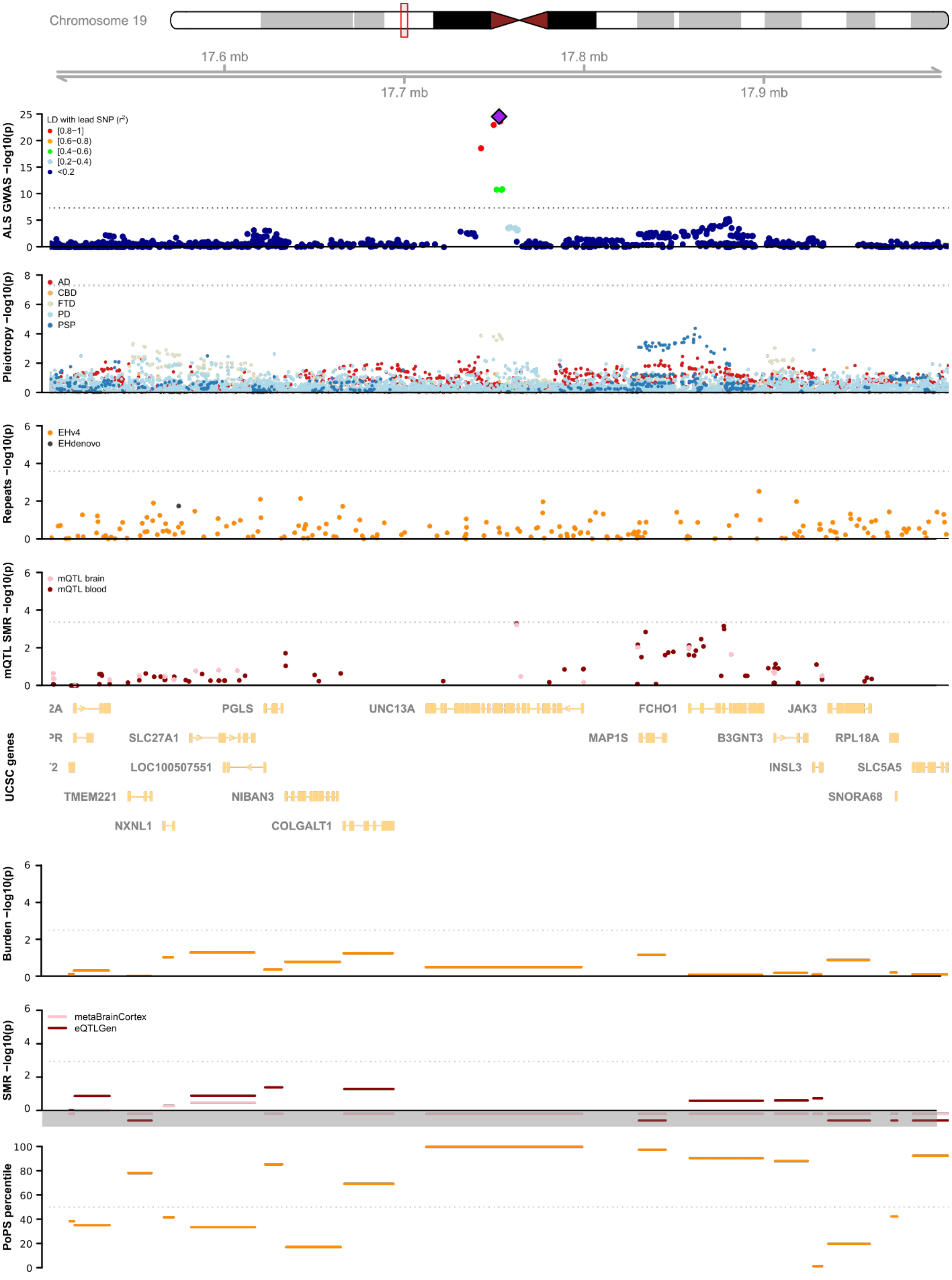

Chromosome 19
17.6 mb
17.7 mb
17.8 mb
17.9 mb
ALS GWAS −log10(p)
LD with lead SNP ($r^2$)
[0.8−1]
[0.6−0.8)
[0.4−0.6)
[0.2−0.4)
<0.2
Pleiotropy −log10(p)
AD
CBD
FTD
PD
PSP
Repeats −log10(p)
EHv4
EHdenovo
mQTL SMR −log10(p)
mQTL brain
mQTL blood
UCSC genes
PGLS
UNC13A
FCHO1
JAK3
SLC27A1
MAP1S
B3GNT3
RPL18A
LOC100507551
INSL3
SLC5A5
TMEM221
NIBAN3
SNORA68
NXNL1
COLGALT1
Burden −log10(p)
SMR −log10(p)
metaBrainCortex
eQTLGen
PoPS percentile

**Supplementary figure 45. Trackplot for rs17785991, *SLC9A8*/*SPATA2*.** The rs17785991 locus consists of a long LD block spanning multiple genes. The strongest brain-mQTL identified CpG sites that were predicted to change expression of *SPATA2*, but SMR revealed multiple CpG sites spanning this region. eQTL SMR on the other hand highlighted *SLC9A8*, with a strong brain-specific signal with no evidence for heterogeneity ($b_{xy}$ = 0.195 ± 0.042, $P_{SMR}$ = 2.50 × $10^{-6}$, $P_{HEIDI}$ = 0.38).
[figure on next page]

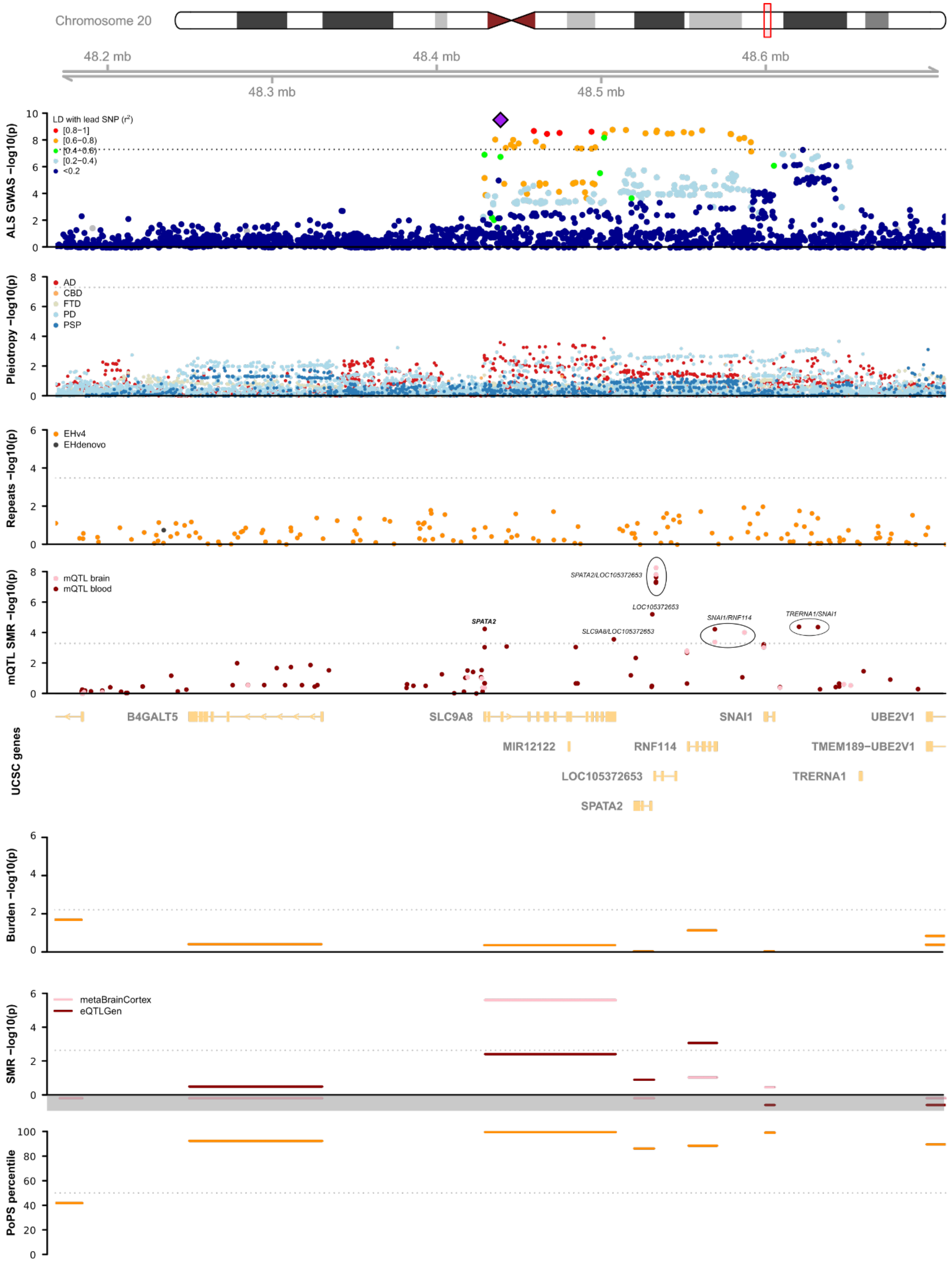
Chromosome 20
48.2 mb
48.4 mb
48.6 mb
48.3 mb
48.5 mb
ALS GWAS −log10(p)
LD with lead SNP (r2)
[0.8−1]
[0.6−0.8)
[0.4−0.6)
[0.2−0.4)
<0.2
Pleiotropy −log10(p)
AD
CBD
FTD
PD
PSP
Repeats −log10(p)
EHv4
EHdenovo
mQTL SMR −log10(p)
mQTL brain
mQTL blood
SPATA2/LOC105372653
LOC105372653
SNAI1/RNF114
TRERNA1/SNAI1
SPATA2
SLC9A8/LOC105372653
UCSC genes
B4GALT5
SLC9A8
SNAI1
UBE2V1
MIR12122
RNF114
TMEM189−UBE2V1
LOC105372653
TRERNA1
SPATA2
Burden −log10(p)
SMR −log10(p)
metaBrainCortex
eQTLGen
PoPS percentile

**Supplementary figure 46. Trackplot for rs80265967, *SOD1.*** The lead SNP rs80265967 is the coding (D90A) variant in *SOD1*. *SOD1* was the first gene described to cause ALS with a predominant autosomal dominant mode of inheritance. The D90A variant has been associated with ALS with an autosomal recessive mode of inheritance and is more common in the Finish population. The rare variant burden of *SOD1* was associated with ALS ($P_{RVB} = 2.4 \times 10^{-4}$ for all disruptive, damaging and non-synonymous variants with MAF < 0.01) which passed the threshold after correcting for multiple testing within this locus (0.05/16 = $3.1 \times 10^{-3}$). This association was not driven by the D90A variant due to the fact that no Finnish ALS cases were included in the WGS cohort. Conditioning on the D90A did not change the association statistics for rare variants within *SOD1* in the individuals overlapping the WGS and GWAS cohort ($P_{RVB} = 4.5 \times 10^{-4}$, $P_{RVB|SNP} = 4.5 \times 10^{-4}$). A dinucleotide repeat expansion intronic of *TIAM1* was associated with ALS (threshold > 24 dinucleotides, $P = 7.0 \times 10^{-5}$, FDR = $1.3 \times 10^{-3}$), which passed the threshold after correcting for multiple testing within this locus. This repeat is in LD with the D90A variant in *SOD1* ($r^2 = 2.7 \times 10^{-4}$, $|D'| = 0.39$).
[figure on next page]

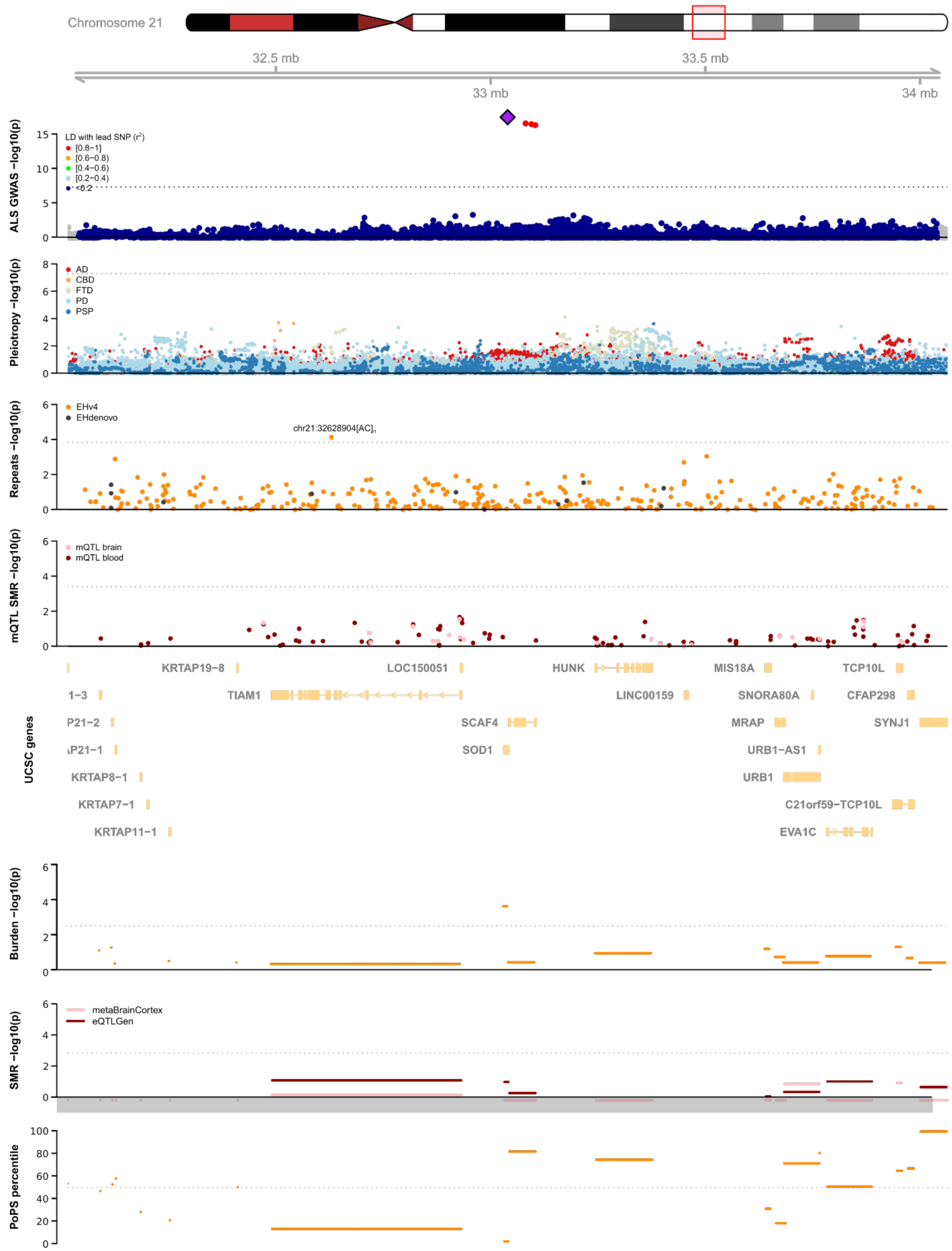
Chromosome 21
32.5 mb
33 mb
33.5 mb
34 mb
ALS GWAS −log10(p)
LD with lead SNP (r²)
[0.8−1]
[0.6−0.8)
[0.4−0.6)
[0.2−0.4)
<0.2
Pleiotropy −log10(p)
AD
CBD
FTD
PD
PSP
Repeats −log10(p)
EHv4
EHdenovo
chr21:32628904[AC]n
mQTL SMR −log10(p)
mQTL brain
mQTL blood
UCSC genes
KRTAP19−8
LOC150051
HUNK
MIS18A
TCP10L
TIAM1
LINC00159
SNORA80A
CFAP298
SCAF4
MRAP
SYNJ1
SOD1
URB1−AS1
KRTAP8−1
URB1
KRTAP7−1
C21orf59−TCP10L
KRTAP11−1
EVA1C
Burden −log10(p)
SMR −log10(p)
metaBrainCortex
eQTLGen
PoPS percentile

**Supplementary figure 47. Trackplot for rs75087725, *CFAP410*.** The rs75087725 has previously been described in the discovery of the novel ALS gene *C21orf2* (ref [28]), later named *CFAP410*. This is a known coding variant of *CFAP410* (p.V58L). The rare variant burden association of *CFAP410* was nominally significant ($P_{RVB} = 7.7 \times 10^{-3}$ for all disruptive, damaging and non-synonymous variants with MAF < 0.01), which did not meet the threshold for multiple testing correction within this locus ($0.05/40 = 1.25 \times 10^{-3}$), but was independent from rs75087725.

[figure on next page]

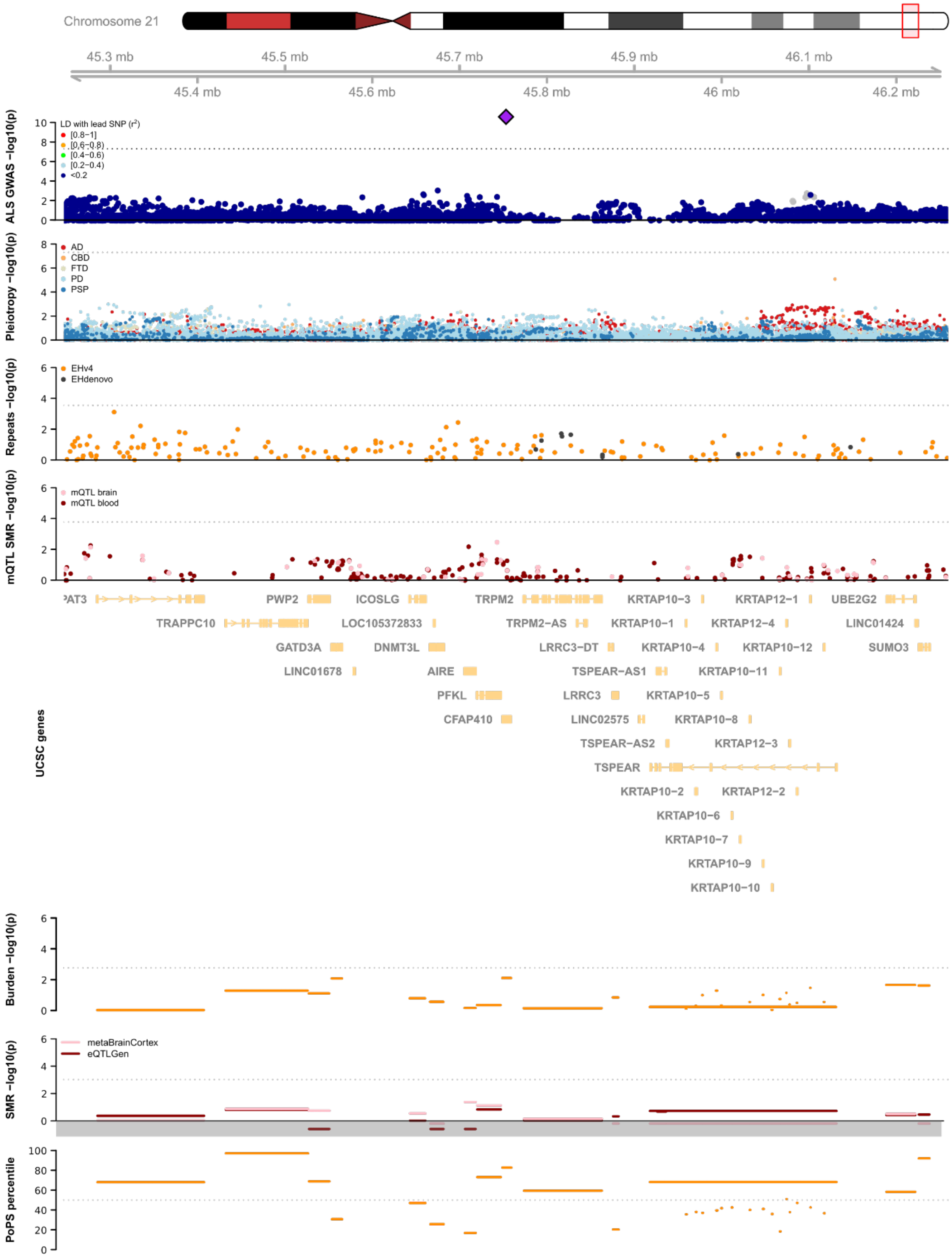

Chromosome 21
45.3 mb
45.5 mb
45.7 mb
45.9 mb
46.1 mb
45.4 mb
45.6 mb
45.8 mb
46 mb
46.2 mb
ALS GWAS −log10(p)
LD with lead SNP (r²)
[0.8−1]
[0.6−0.8)
[0.4−0.6)
[0.2−0.4)
<0.2
Pleiotropy −log10(p)
AD
CBD
FTD
PD
PSP
Repeats −log10(p)
EHv4
EHdenovo
mQTL SMR −log10(p)
mQTL brain
mQTL blood
UCSC genes
PAT3
TRAPPC10
PWP2
GATD3A
LINC01678
ICOSLG
LOC105372833
DNMT3L
AIRE
PFKL
CFAP410
TRPM2
TRPM2−AS
LRRC3−DT
TSPEAR−AS1
LRRC3
LINC02575
TSPEAR−AS2
TSPEAR
KRTAP10−3
KRTAP10−1
KRTAP10−4
KRTAP10−5
KRTAP10−8
KRTAP10−2
KRTAP10−6
KRTAP10−7
KRTAP10−9
KRTAP10−10
KRTAP12−1
KRTAP12−4
KRTAP10−12
KRTAP10−11
KRTAP12−3
KRTAP12−2
UBE2G2
LINC01424
SUMO3
Burden −log10(p)
SMR −log10(p)
metaBrainCortex
eQTLGen
PoPS percentile

**Supplementary figure 48. *NEK1* repeat distribution.** The frequency of various repeat lengths among ALS cases and controls are shown. A repeat length of 11 and longer was used as threshold for disease-associated genotype. The P-value was calculated by Firth logistic regression. Repeat position on GRCh37, and repeat motif are shown.

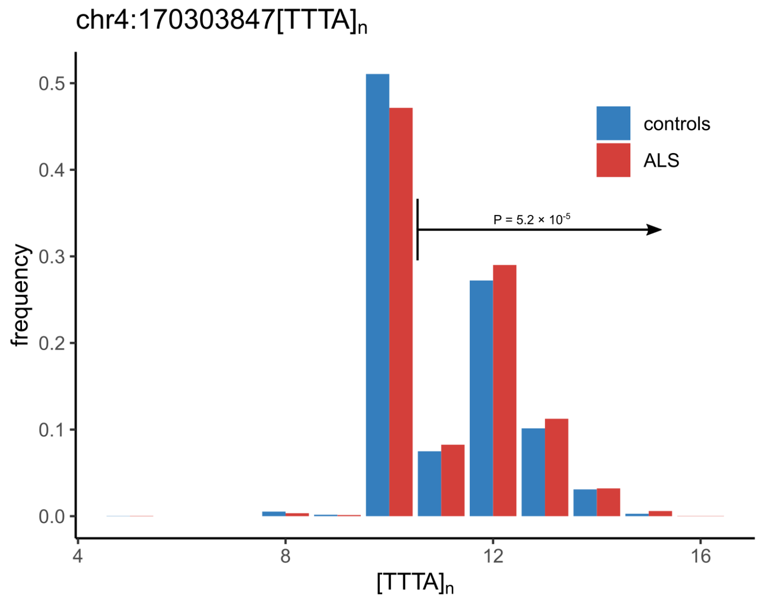


**Supplementary figure 49. Genetic correlation power calculation.** Power calculations to detect a statistically significant genetic correlation ($P < 0.05$) between ALS and a secondary neurodegenerative trait, given the current sample size and observed h2 estimate using LD-score regression. Power is defined as 1 - type II error rate at $P = 0.05$. Power calculations were based on ref 29.

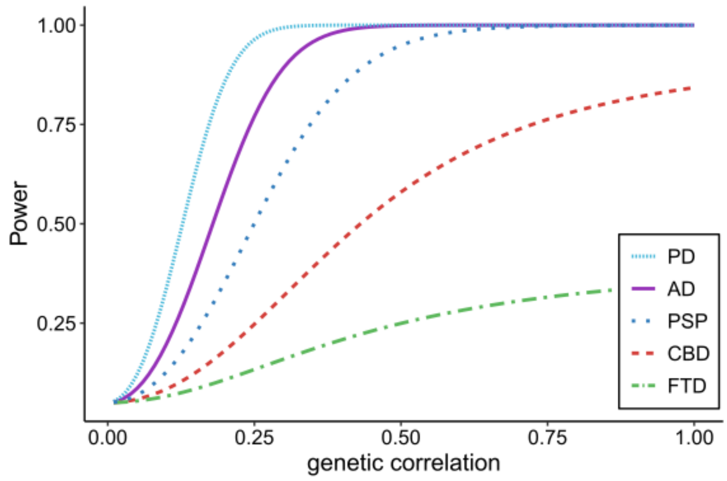

**Supplementary figure 50. Colocalization signals.** Loci were selected for colocalization analysis with COLOC if a SNP was present with a genome-wide significant association ($P < 5 \times 10^{-8}$) in one trait and a P-value below $5 \times 10^{-5}$ in the other. For ALS, the European ancestry-only meta-analysis was used. Posterior probabilities of the same variant driving the both traits are reported below locus names.

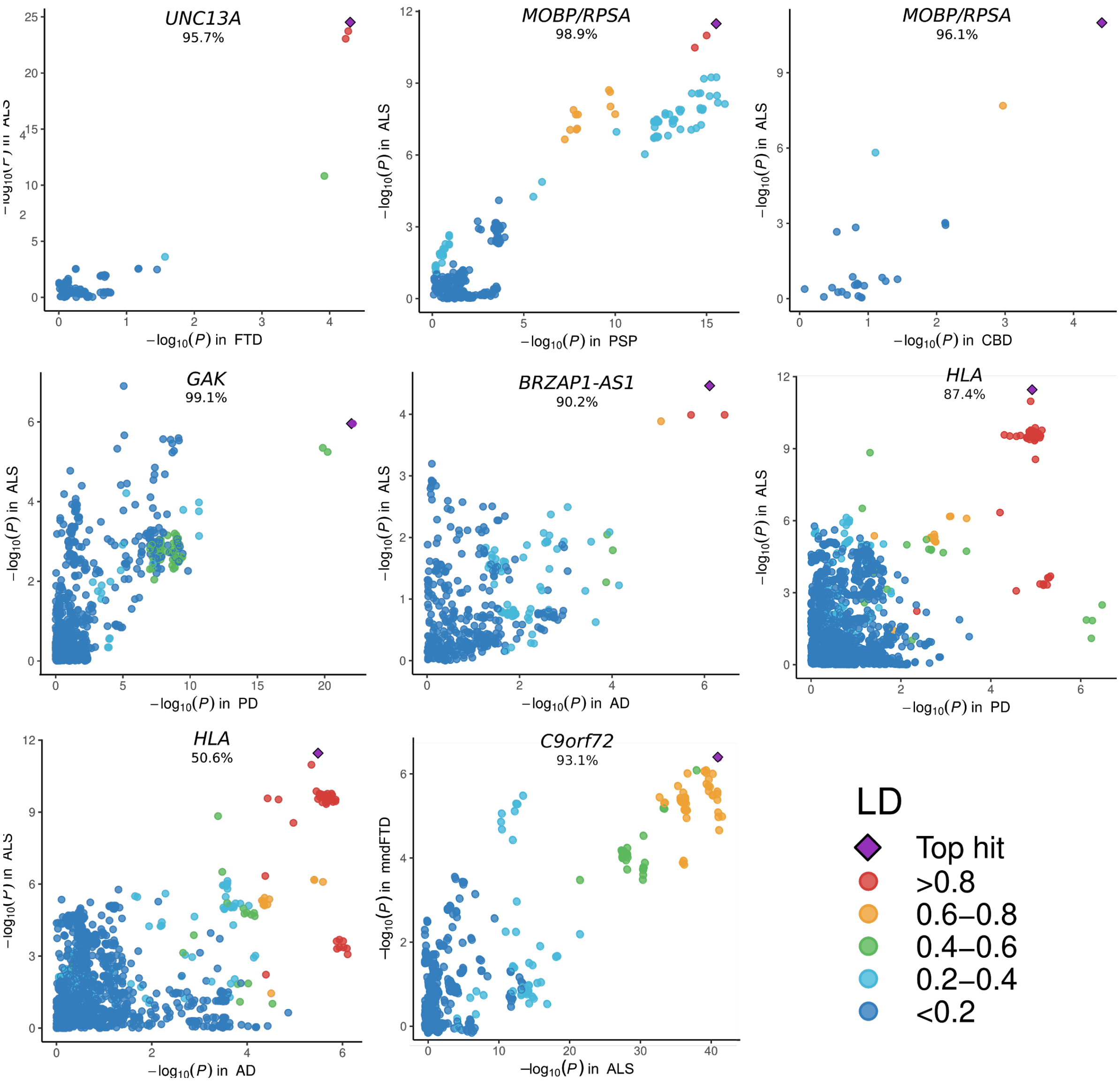

**Supplementary figure 51. Colocalization analysis with FTD subtypes.** SNP association P-values of the lead SNPs in the ALS GWAS were selected for colocalization analysis between ALS and FTD subtypes using COLOC. In the top panel, point height is -log$_{10}$(P) of the lead SNP in the European ancestry ALS GWAS. In the bottom panel, association P-values of these SNPs with FTD subtypes are shown by color. The posterior probability of both traits driven by the same variant are depicted by a connection between points.

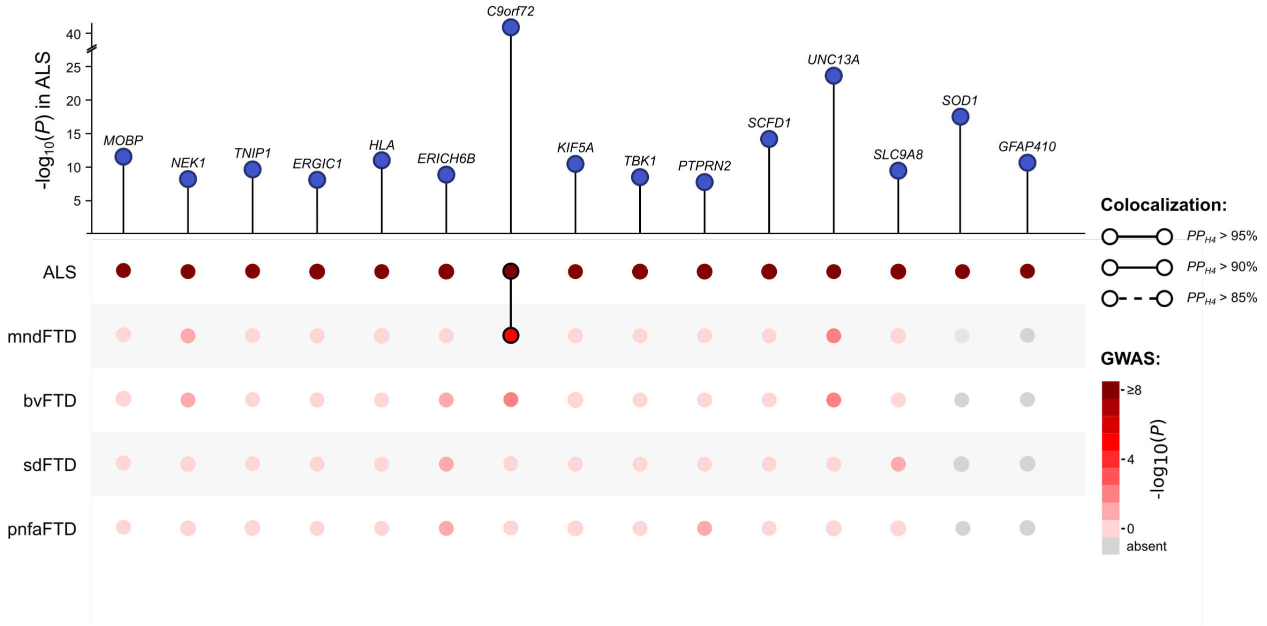


**Supplementary figure 52. Cell-type enrichment analysis in mice.** Cell-type enrichment analysis using the DropViz single-cell RNA sequencing dataset obtained from mice. Similar to the cell-type enrichment analyses there is neuron-specific enrichment in ALS and Parkinson's disease. In Alzheimer's disease microglia are the most enriched celltypes. Statistically significant enrichment after correction for multiple testing with a false discovery rate (FDR) < 0.05 are marked with an asterisk.

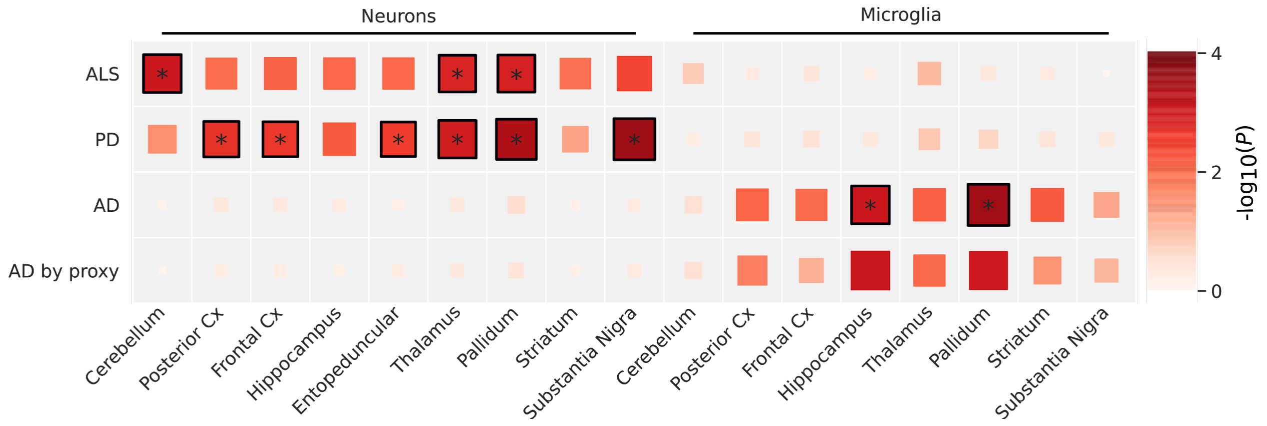

**Supplementary figure 53. Human phenotype ontology term enrichment.** Downstreamer enrichment analyses were performed using the multi-tissue and brain-specific co-expression matrix to identify co-regulated ALS-genes. The distribution of enrichment statistics (Z-scores) for all HPO terms are plotted per HPO parent branch. The multi-tissue analysis indicates enrichment for the neurology parent branch "*abnormality of the nervous system"* (dark-red), although no term passes the Bonferroni threshold for multiple testing. The brain-specific analysis illustrates stronger enrichment for the neurology parent branch where 58 HPO terms pass the threshold for multiple testing of which 42 are defined within *"abnormality of the nervous system"* branch.

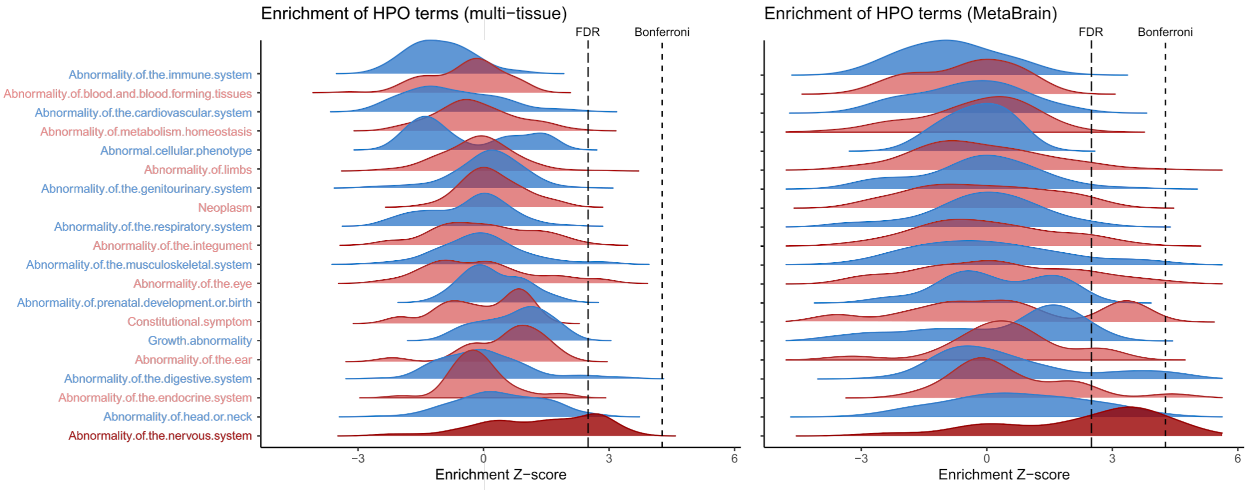

**Supplementary figures 54-59. Principal components plots per stratum including HapMap3 reference populations.**

**Supplementary figure 54 - PCA plot for stratum 1**

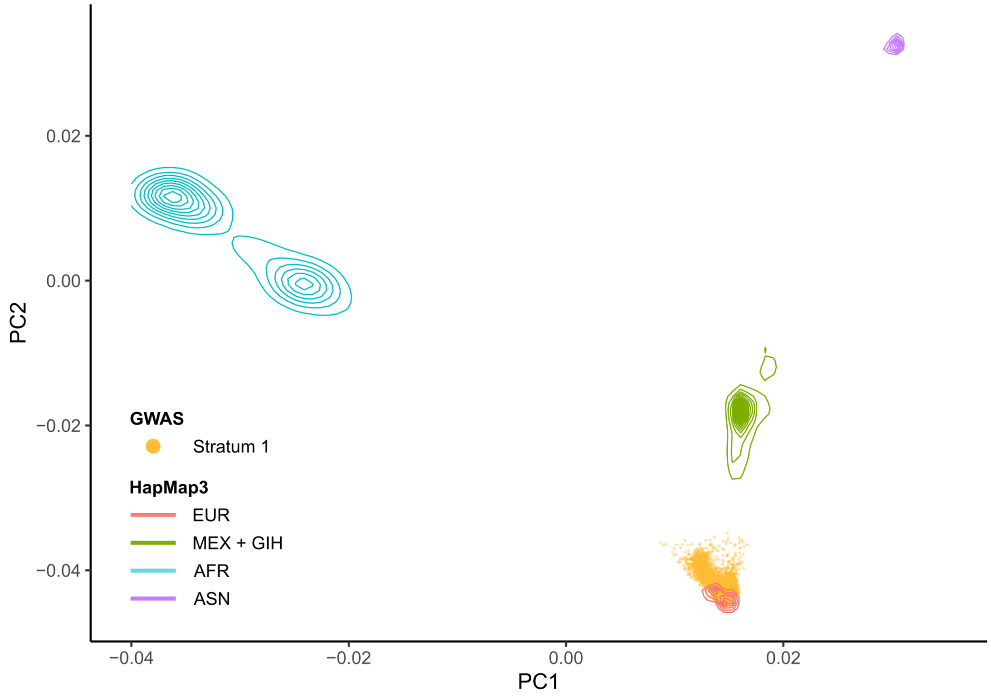


**Supplementary figure 55 - PCA plot for stratum 2**

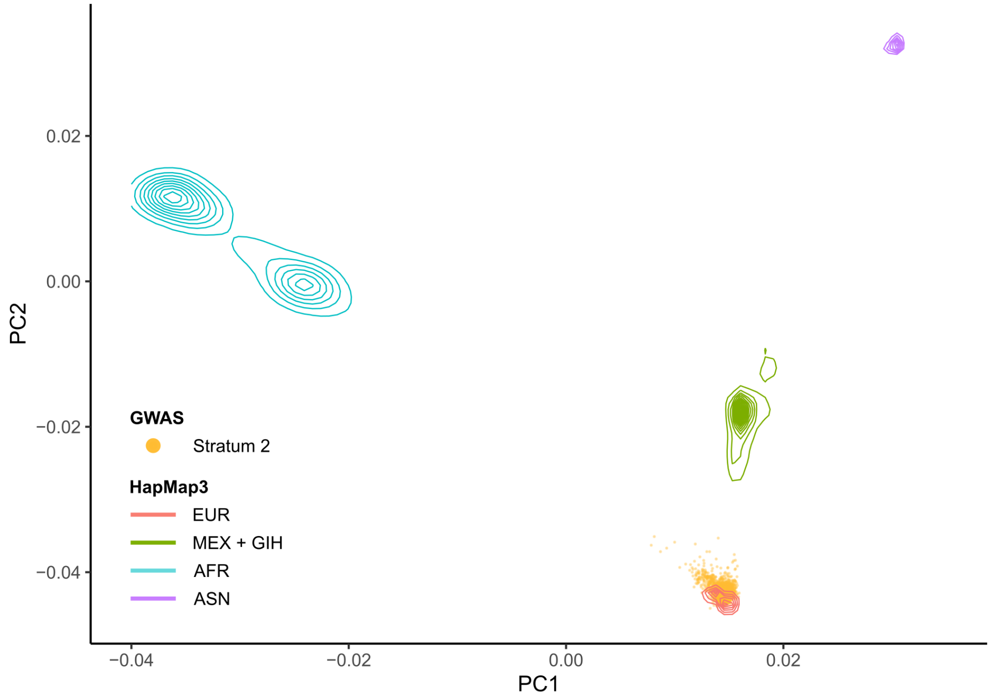

**Supplementary figure 56 - PCA plot for stratum 3**

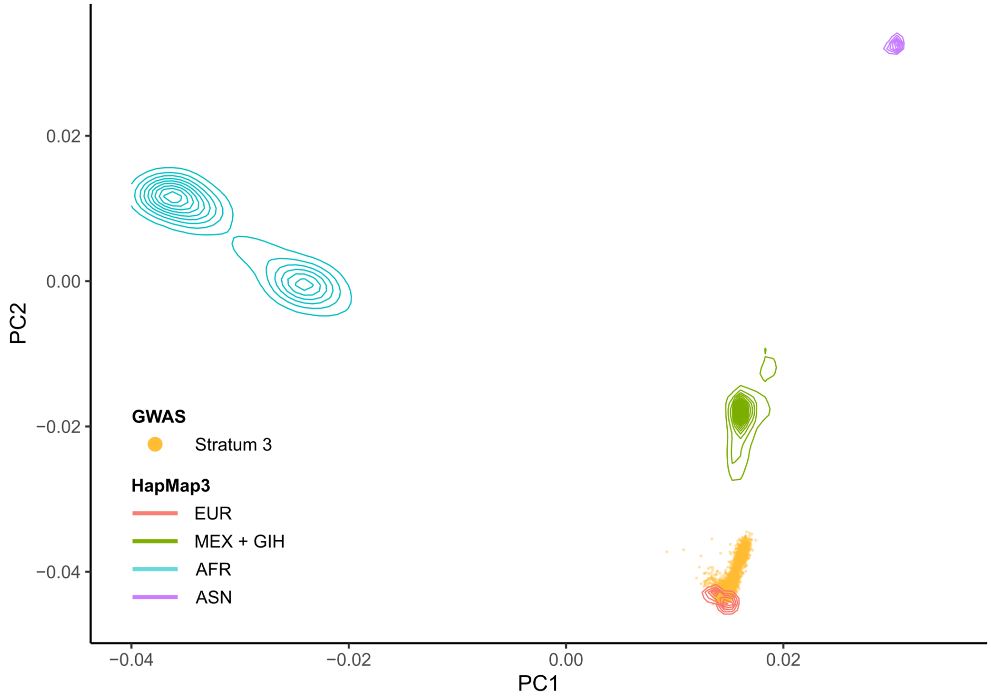


**Supplementary figure 57 - PCA plot for stratum 4**

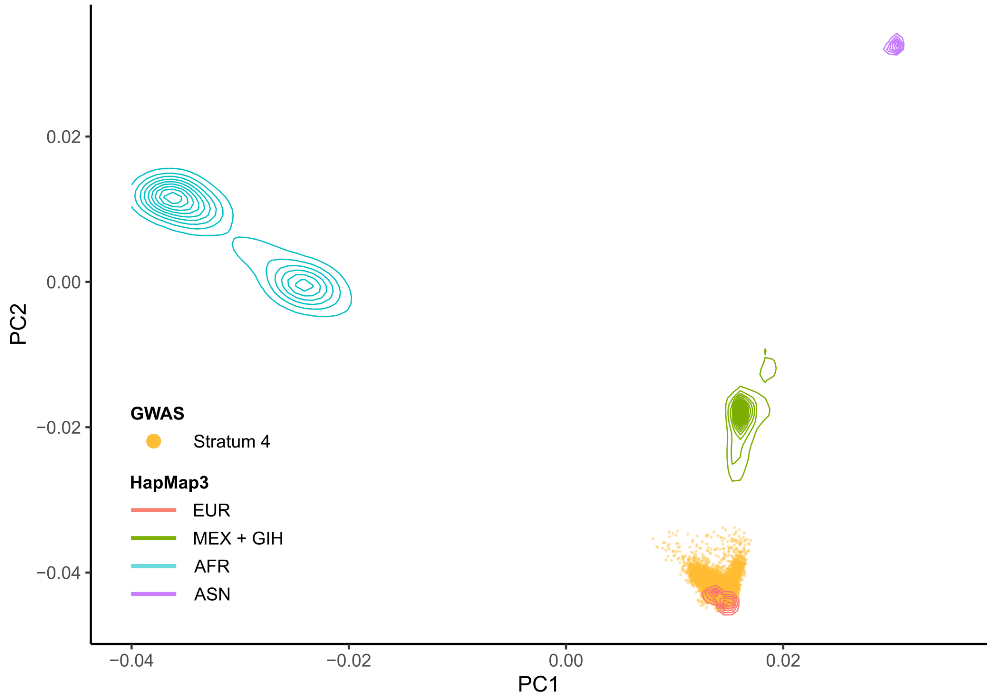

**Supplementary figure 58 - PCA plot for stratum 6**

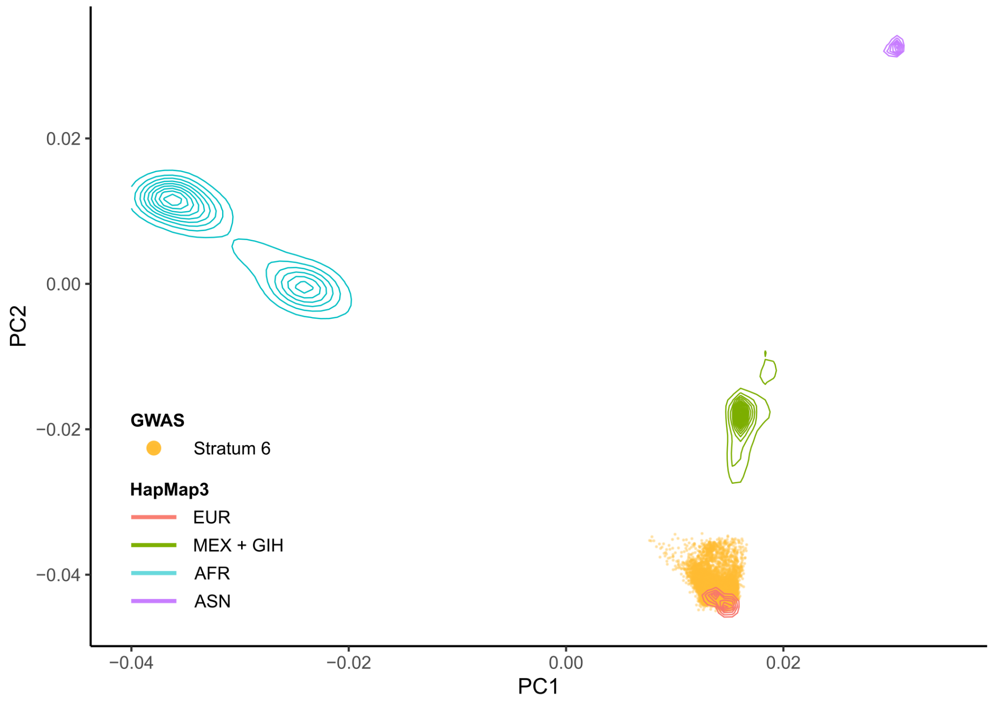


**Supplementary figure 59 - PCA plot for stratum 7**

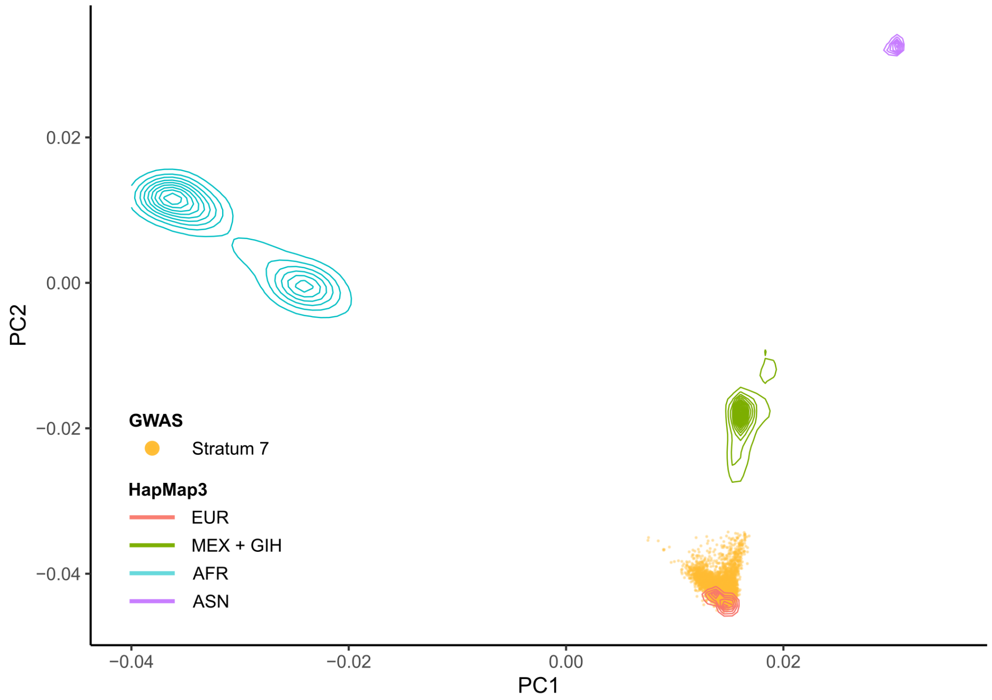

### Supplementary Tables

**Supplementary table 1. GWAS Cohort description**
[table in Excel file]

**Supplementary table 2. GWAS Quality control details**
[table in Excel file]

**Supplementary table 3. WGS Quality control details**
[table in Excel file]

**Supplementary tables 4-18. Details for genome-wide significant loci.** SNP ID, effect allele, and annotated genes are reported above each Table. European ancestries is the meta-analysis of strata s1, s2, s3, s4, s6, and s7. Cross-ancestry is the meta-analysis of European ancestries, China and Japan. Freq = effect allele frequency, $N_{eff}$ = effective sample size.

**Supplementary table 4. Details for rs631312 (G), MOBP/*RPSA***

| Stratum | Freq | INFO | beta | SE | P | $N_{eff}$ |
|---|---|---|---|---|---|---|
| s1 | 0.28 | 1.00 | 0.12 | 0.038 | $2.07\times10^{-3}$ | 7,500 |
| s2 | 0.29 | 0.98 | 0.08 | 0.055 | 0.122 | 3,403 |
| s3 | 0.30 | 0.98 | 0.12 | 0.052 | 0.024 | 4,085 |
| s4 | 0.28 | 0.98 | 0.11 | 0.033 | $9.57\times10^{-4}$ | 12,570 |
| s6 | 0.29 | 1.00 | 0.05 | 0.017 | 0.006 | 39,764 |
| s7 | 0.30 | 0.95 | 0.12 | 0.031 | $6.29\times10^{-5}$ | 13,391 |
| European ancestries | 0.29 | - | 0.08 | 0.012 | $5.24\times10^{-11}$ | 80,713 |
| China | - | - | 0.04 | 0.049 | 0.405 | 3,445 |
| Japan | - | - | 0.13 | 0.053 | 0.012 | 4,147 |
| Cross-ancestry | - | - | 0.08 | 0.011 | $3.28\times10^{-12}$ | 88,305 |

**Supplementary table 5. Details for rs62333164 (A), *NEK1***

| Stratum | Freq | INFO | beta | SE | P | $N_{eff}$ |
|---|---|---|---|---|---|---|
| s1 | 0.33 | 0.95 | 0.06 | 0.038 | 0.09 | 7,500 |
| s2 | 0.34 | 0.99 | 0.01 | 0.054 | 0.82 | 3,403 |
| s3 | 0.34 | 1.00 | 0.08 | 0.050 | 0.12 | 4,085 |
| s4 | 0.33 | 1.00 | 0.03 | 0.032 | 0.27 | 12,570 |
| s6 | 0.34 | 0.98 | 0.08 | 0.016 | $4.7\times10^{-6}$ | 39,764 |
| s7 | 0.33 | 0.96 | 0.06 | 0.030 | 0.05 | 13,391 |
| European ancestries | 0.33 | - | 0.06 | 0.012 | $7.0\times10^{-8}$ | 80,713 |
| China | - | - | 0.19 | 0.094 | 0.04 | 3,445 |
| Japan | - | - | 0.22 | 0.106 | 0.04 | 4,147 |
| Cross-ancestry | - | - | 0.07 | 0.012 | $6.9\times10^{-9}$ | 88,305 |

**Supplementary table 6. Details for rs10463311 (C), *GPX3/TNIP1***

| Stratum | Freq | INFO | beta | SE | P | $N_{eff}$ |
|---|---|---|---|---|---|---|
| s1 | 0.25 | 0.990 | 0.12 | 0.04 | 0.003 | 7,500 |
| s2 | 0.24 | 0.987 | 0.10 | 0.06 | 0.083 | 3,403 |
| s3 | 0.25 | 0.997 | 0.08 | 0.05 | 0.150 | 4,085 |
| s4 | 0.25 | 0.996 | 0.11 | 0.03 | $6.30\times10^{-4}$ | 12,570 |
| s6 | 0.26 | 0.993 | 0.07 | 0.02 | $8.30\times10^{-5}$ | 39,764 |
| s7 | 0.25 | 0.950 | 0.05 | 0.03 | 0.156 | 13,391 |
| European ancestries | 0.25 | - | 0.08 | 0.01 | $3.46\times10^{-10}$ | 80,713 |
| China | - | - | 0.13 | 0.05 | 0.009 | 3,445 |
| Japan | - | - | -0.06 | 0.05 | 0.284 | 4,147 |
| Cross-ancestry | - | - | 0.08 | 0.01 | $2.73\times10^{-10}$ | 88,305 |

**Supplementary table 7. Details for rs517339 (C), *ERGIC1***

| Stratum | Freq | INFO | beta | SE | P | $N_{eff}$ |
|---|---|---|---|---|---|---|
| s1 | 0.40 | 1.000 | -0.06 | 0.035 | 0.11 | 7,500 |
| s2 | 0.40 | 0.998 | -0.06 | 0.051 | 0.23 | 3,403 |
| s3 | 0.38 | 0.999 | -0.07 | 0.048 | 0.14 | 4,085 |
| s4 | 0.40 | 1.000 | -0.07 | 0.030 | 0.02 | 12,570 |
| s6 | 0.40 | 0.998 | -0.07 | 0.016 | $2.5\times10^{-6}$ | 39,764 |
| s7 | 0.40 | 0.976 | -0.03 | 0.029 | 0.29 | 13,391 |
| European ancestries | 0.40 | - | -0.06 | 0.011 | $8.5\times10^{-9}$ | 80,713 |
| China | - | - | -0.07 | 0.074 | 0.38 | 3,445 |
| Japan | - | - | - | - | - | |
| Cross-ancestry | - | - | -0.06 | 0.011 | $5.6\times10^{-9}$ | 84,158 |

**Supplementary table 8. Details for rs9275477 (C), *HLA***

| Stratum | Freq | INFO | beta | SE | P | $N_{eff}$ |
|---|---|---|---|---|---|---|
| s1 | 0.10 | 0.96 | -0.230 | 0.060 | $1.1\times10^{-4}$ | 7,500 |
| s2 | 0.09 | 0.95 | -0.062 | 0.087 | 0.47 | 3,403 |
| s3 | 0.11 | 0.97 | 0.005 | 0.076 | 0.95 | 4,085 |
| s4 | 0.09 | 0.96 | -0.091 | 0.057 | 0.11 | 12,570 |
| s6 | 0.09 | 0.74 | -0.195 | 0.031 | $2.6\times10^{-10}$ | 39,764 |
| s7 | 0.09 | 0.99 | -0.072 | 0.051 | 0.15 | 13,391 |
| European ancestries | 0.10 | - | -0.143 | 0.021 | $5.5\times10^{-12}$ | 80,713 |
| China | - | - | -0.110 | 0.111 | 0.34 | 3,445 |
| Japan | - | - | - | - | - | - |
| Cross-ancestry | - | - | -0.142 | 0.020 | $3.5\times10^{-12}$ | 84,158 |

**Supplementary table 9. Details for rs10280711 (G), *PTPRN2***

| Stratum | Freq | INFO | beta | SE | P | $N_{eff}$ |
|---|---|---|---|---|---|---|
| s1 | 0.13 | 0.80 | 0.082 | 0.058 | 0.156 | 7,500 |
| s2 | 0.12 | 0.95 | 0.065 | 0.077 | 0.398 | 3,403 |
| s3 | 0.13 | 0.99 | 0.048 | 0.072 | 0.507 | 4,085 |
| s4 | 0.12 | 0.99 | 0.155 | 0.044 | $4.40\times10^{-4}$ | 12,570 |
| s6 | 0.12 | 0.98 | 0.076 | 0.023 | $1.13\times10^{-3}$ | 39,764 |
| s7 | 0.13 | 0.91 | 0.012 | 0.043 | 0.78 | 13,391 |
| European ancestries | 0.12 | - | 0.076 | 0.017 | $5.76\times10^{-6}$ | 80,713 |
| China | - | - | 0.139 | 0.048 | 0.005 | 3,445 |
| Japan | - | - | 0.122 | 0.056 | 0.029 | 4,147 |
| Cross-ancestry | - | - | 0.086 | 0.015 | $1.78\times10^{-8}$ | 88,305 |

**Supplementary table 10. Details for rs2453555 (A), *C9orf72***

| Stratum | Freq | INFO | beta | SE | P | $N_{eff}$ |
|---|---|---|---|---|---|---|
| s1 | 0.24 | 0.93 | 0.16 | 0.04 | $1.84\times10^{-4}$ | 7,500 |
| s2 | 0.24 | 0.99 | 0.17 | 0.06 | 0.004 | 3,403 |
| s3 | 0.24 | 1.00 | 0.30 | 0.05 | $3.26\times10^{-8}$ | 4,085 |
| s4 | 0.24 | 0.99 | 0.17 | 0.03 | $5.67\times10^{-7}$ | 12,570 |
| s6 | 0.25 | 1.00 | 0.18 | 0.02 | $7.50\times10^{-25}$ | 39,764 |
| s7 | 0.24 | 0.98 | 0.12 | 0.03 | $3.42\times10^{-4}$ | 13,391 |
| European ancestries | 0.25 | - | 0.17 | 0.01 | $9.98\times10^{-43}$ | 80,713 |
| China | - | - | -0.07 | 0.10 | 0.514 | 3,445 |
| Japan | - | - | 0.08 | 0.09 | 0.350 | 4,147 |
| Cross-ancestry | - | - | 0.17 | 0.01 | $1.48\times10^{-41}$ | 88,305 |

**Supplementary table 11. Details for rs113247976 (T), *KIF5A***

| Stratum | Freq | INFO | beta | SE | P | $N_{eff}$ |
|---|---|---|---|---|---|---|
| s1 | 0.014 | 0.82 | 0.26 | 0.16 | 0.105 | 7,500 |
| s2 | 0.016 | 0.89 | 0.47 | 0.21 | 0.023 | 3,403 |
| s3 | 0.025 | 0.93 | 0.27 | 0.15 | 0.081 | 4,085 |
| s4 | 0.014 | 0.88 | 0.13 | 0.15 | 0.385 | 12,570 |
| s6 | 0.015 | 0.83 | 0.38 | 0.07 | $3.48\times10^{-8}$ | 39,764 |
| s7 | 0.012 | 0.85 | 0.36 | 0.14 | 0.008 | 13,391 |
| European ancestries | 0.016 | - | 0.33 | 0.05 | $1.42\times10^{-11}$ | 80,713 |
| China | - | - | - | - | - | - |
| Japan | - | - | - | - | - | - |
| Cross-ancestry | - | - | 0.33 | 0.05 | $1.42\times10^{-11}$ | 80,713 |

**Supplementary table 12. Details for rs4075094 (A), *TBK1***

| Stratum | Freq | INFO | beta | SE | P | $N_{eff}$ |
|---|---|---|---|---|---|---|
| s1 | 0.11 | 0.97 | -0.08 | 0.06 | 0.14 | 7,500 |
| s2 | 0.11 | 0.98 | -0.03 | 0.08 | 0.75 | 3,403 |
| s3 | 0.11 | 0.99 | -0.16 | 0.07 | 0.03 | 4,085 |
| s4 | 0.11 | 0.98 | -0.07 | 0.04 | 0.14 | 12,570 |
| s6 | 0.11 | 0.94 | -0.08 | 0.02 | $6.9\times10^{-4}$ | 39,764 |
| s7 | 0.10 | 0.97 | -0.19 | 0.04 | $2.7\times10^{-5}$ | 13,391 |
| European ancestries | 0.11 | - | -0.10 | 0.02 | $1.7\times10^{-8}$ | 80,713 |
| China | - | - | -0.28 | 0.11 | 0.02 | 3,445 |
| Japan | - | - | -0.10 | 0.15 | 0.52 | 4,147 |
| Cross-ancestry | - | - | -0.10 | 0.02 | $2.1\times10^{-9}$ | 88,305 |

**Supplementary table 13. Details for rs2985994 (C), *COG3***

| Stratum | Freq | INFO | beta | SE | P | $N_{eff}$ |
|---|---|---|---|---|---|---|
| s1 | 0.26 | 0.97 | 0.11 | 0.040 | 0.004 | 7,500 |
| s2 | 0.26 | 0.98 | 0.07 | 0.057 | 0.225 | 3,403 |
| s3 | 0.27 | 0.99 | 0.05 | 0.053 | 0.344 | 4,085 |
| s4 | 0.26 | 0.99 | 0.06 | 0.035 | 0.078 | 12,570 |
| s6 | 0.26 | 0.98 | 0.06 | 0.018 | 0.0012 | 39,764 |
| s7 | 0.26 | 0.95 | 0.07 | 0.033 | 0.029 | 13,391 |
| European ancestries | 0.26 | - | 0.07 | 0.013 | $1.87\times10^{-7}$ | 80,713 |
| China | - | - | 0.07 | 0.054 | 0.188 | 3,445 |
| Japan | - | - | 0.14 | 0.062 | 0.026 | 4,147 |
| Cross-ancestry | - | - | 0.07 | 0.012 | $1.16\times10^{-8}$ | 88,305 |

**Supplementary table 14. Details for rs229195 (A), *SCFD1***

| Stratum | Freq | INFO | beta | SE | P | $N_{eff}$ |
|---|---|---|---|---|---|---|
| s1 | 0.33 | 0.981 | 0.14 | 0.037 | $1.70\times10^{-4}$ | 7,500 |
| s2 | 0.34 | 0.997 | 0.02 | 0.053 | 0.673 | 3,403 |
| s3 | 0.34 | 0.999 | 0.10 | 0.049 | 0.037 | 4,085 |
| s4 | 0.34 | 0.999 | 0.10 | 0.031 | 0.0010 | 12,570 |
| s6 | 0.34 | 0.950 | 0.11 | 0.017 | $2.70\times10^{-10}$ | 39,764 |
| s7 | 0.33 | 0.939 | 0.02 | 0.031 | 0.522 | 13,391 |
| European ancestries | 0.34 | - | 0.09 | 0.012 | $9.20\times10^{-15}$ | 80,713 |
| China | - | - | - | - | - | |
| Japan | - | - | - | - | - | |
| Cross-ancestry | - | - | 0.09 | 0.012 | $9.20\times10^{-15}$ | 80,713 |

**Supplementary table 15. Details for rs12608932 (C), *UNC13A***

| Stratum | Freq | INFO | beta | SE | P | $N_{eff}$ |
|---|---|---|---|---|---|---|
| s1 | - | 0.514 | - | - | - | - |
| s2 | 0.35 | 0.997 | 0.12 | 0.05 | 0.02 | 3,403 |
| s3 | 0.39 | 0.998 | 0.25 | 0.05 | $1.6\times10^{-7}$ | 4,085 |
| s4 | 0.34 | 0.997 | 0.07 | 0.03 | 0.03 | 12,570 |
| s6 | 0.34 | 0.997 | 0.12 | 0.02 | $1.3\times10^{-13}$ | 39,764 |
| s7 | 0.35 | 0.988 | 0.14 | 0.03 | $9.1\times10^{-7}$ | 13,391 |
| European ancestries | 0.35 | - | 0.12 | 0.01 | $8.8\times10^{-25}$ | 73,213 |
| China | - | - | 0.05 | 0.05 | 0.29 | 3,445 |
| Japan | - | - | 0.11 | 0.06 | 0.08 | 4,147 |
| Cross-ancestry | - | - | 0.12 | 0.01 | $3.0\times10^{-25}$ | 80,805 |

**Supplementary table 16. Details for rs17785991 (A), *SLC9A8/SPATA2***

| Stratum | Freq | INFO | beta | SE | P | $N_{eff}$ |
|---|---|---|---|---|---|---|
| s1 | 0.34 | 0.97 | 0.09 | 0.037 | 0.01 | 7,500 |
| s2 | 0.36 | 0.90 | 0.09 | 0.055 | 0.10 | 3,403 |
| s3 | 0.31 | 0.93 | 0.12 | 0.053 | 0.02 | 4,085 |
| s4 | 0.35 | 0.95 | 0.05 | 0.032 | 0.11 | 12,570 |
| s6 | 0.36 | 0.93 | 0.06 | 0.017 | $1.1\times10^{-4}$ | 39,764 |
| s7 | 0.34 | 0.97 | 0.10 | 0.030 | $1.2\times10^{-3}$ | 13,391 |
| European ancestries | 0.35 | - | 0.07 | 0.012 | $3.5\times10^{-10}$ | 80,713 |
| China | - | - | - | - | - | - |
| Japan | - | - | 0.04 | 0.076 | 0.55 | 4,147 |
| Cross-ancestry | - | - | 0.07 | 0.012 | $3.2\times10^{-10}$ | 84,860 |

**Supplementary table 17. Details for rs80265967 (C), *SOD1***

| Stratum | Freq | INFO | beta | SE | P | $N_{eff}$ |
|---|---|---|---|---|---|---|
| s1 | - | 0.61 | - | - | - | - |
| s2 | 0.0006 | 0.79 | 0.08 | 1.21 | 0.95 | 3,403 |
| s3 | 0.0105 | 0.96 | 0.91 | 0.17 | $1.0\times10^{-7}$ | 4,085 |
| s4 | 0.0007 | 0.82 | 1.07 | 0.48 | 0.03 | 12,570 |
| s6 | 0.0009 | 0.80 | 1.27 | 0.23 | $2.3\times10^{-8}$ | 39,764 |
| s7 | 0.0008 | 0.98 | 1.56 | 0.40 | $1.2\times10^{-4}$ | 13,391 |
| European ancestries | 0.0060 | - | 1.08 | 0.12 | $3.5\times10^{-18}$ | 73,213 |
| China | - | - | - | - | - | - |
| Japan | - | - | - | - | - | - |
| Cross-ancestry | - | - | 1.08 | 0.12 | $3.5\times10^{-18}$ | 73,123 |

**Supplementary table 18. Details for rs75087725 (A), *CFAP410***

| Stratum | Freq | INFO | beta | SE | P | $N_{eff}$ |
|---|---|---|---|---|---|---|
| s1 | - | - | - | - | - | - |
| s2 | - | - | - | - | - | - |
| s3 | 0.015 | 0.88 | 0.37 | 0.20 | 0.07 | 4,085 |
| s4 | 0.012 | 0.83 | 0.34 | 0.16 | 0.03 | 12,570 |
| s6 | 0.012 | 0.72 | 0.41 | 0.08 | $8.9\times10^{-7}$ | 39,764 |
| s7 | 0.011 | 0.79 | 0.55 | 0.15 | $2.8\times10^{-4}$ | 13,391 |
| European ancestries | 0.012 | - | 0.42 | 0.06 | $2.7\times10^{-11}$ | 69,810 |
| China | - | - | - | - | - | - |
| Japan | - | - | - | - | - | - |
| Cross-ancestry | - | - | 0.42 | 0.06 | $2.7\times10^{-11}$ | 69,810 |

**Supplementary table 19. Details for rs58854276 (A), *ACSL5/ZDHHC6***

| Stratum | Freq | INFO | beta | SE | P | $N_{eff}$ |
|---|---|---|---|---|---|---|
| s1 | 0.66 | 1.0 | 0.09 | 0.04 | 0.01 | 7,500 |
| s2 | 0.66 | 0.94 | 0.14 | 0.05 | 0.01 | 3,403 |
| s3 | 0.65 | 0.97 | 0.00 | 0.05 | 0.97 | 4,085 |
| s4 | 0.65 | 1.0 | 0.10 | 0.03 | $1.7\times10^{-3}$ | 12,570 |
| s6 | 0.66 | 0.99 | 0.02 | 0.02 | 0.24 | 39,764 |
| s7 | 0.66 | 0.95 | 0.06 | 0.03 | 0.05 | 13,391 |
| European ancestries | 0.66 | - | 0.05 | 0.01 | $5.4\times10^{-4}$ | 80,713 |
| China | - | - | 0.19 | 0.05 | $1.8\times10^{-4}$ | 3,445 |
| Japan | - | - | 0.17 | 0.05 | $1.6\times10^{-3}$ | 4,147 |
| Cross-ancestry | - | - | 0.06 | 0.01 | $6.5\times10^{-8}$ | 88,305 |

**Supplementary table 20. Association signals in neurodegenerative diseases**. Association beta (b) and P-values (p) of ALS lead SNPs for Cross-ancestry ALS, Alzheimer's disease (AD), Parkinson's disease (PD), progressive supranuclear palsy (PSP), frontotemporal dementia (FTD), and corticobasal degeneration (CBD) are shown. EA: effect allele. Name: annotated gene names.

| SNP | EA | Name | $b_{ALS}$ | $p_{ALS}$ | $b_{AD}$ | $p_{AD}$ | $b_{PD}$ | $p_{PD}$ | $b_{PSP}$ | $p_{PSP}$ | $b_{FTD}$ | $p_{FTD}$ | $b_{CBD}$ | $p_{CBD}$ |
|---|---|---|---|---|---|---|---|---|---|---|---|---|---|---|
| rs2453555 | A | *C9orf72* | 0.17 | 1.5E-41 | -0.004 | 0.82 | 0.009 | 0.67 | 0.003 | 0.56 | 0.149 | 6.5E-04 | *NA* | *NA* |
| rs12608932 | C | *UNC13A* | 0.12 | 3.0E-25 | 0 | 1.00 | 0.010 | 0.60 | *NA* | *NA* | 0.159 | 0.00011 | -0.18 | 0.17 |
| rs80265967 | C | *SOD1* | 1.08 | 3.5E-18 | *NA* | *NA* | -1.014 | 0.13 | *NA* | *NA* | *NA* | *NA* | *NA* | *NA* |
| rs229195 | A | *SCFD1* | 0.09 | 9.2E-15 | 0.002 | 0.91 | 0.003 | 0.89 | 0.003 | 0.47 | -0.007 | 0.88 | *NA* | *NA* |
| rs631312 | G | *MOBP/RPSA* | 0.08 | 3.3E-12 | 0.019 | 0.24 | 0.032 | 0.09 | 0.039 | 3.0E-16 | 0.129 | 0.02 | *NA* | *NA* |
| rs9275477 | C | *HLA* | -0.14 | 3.5E-12 | -0.120 | 3.3E-06 | -0.152 | 1.2E-05 | -0.015 | 0.03 | -0.099 | 0.21 | *NA* | *NA* |
| rs113247976 | T | *KIF5A* | 0.33 | 1.4E-11 | 0.002 | 0.98 | 0.121 | 0.13 | 0.007 | 0.69 | *NA* | *NA* | *NA* | *NA* |
| rs75087725 | A | *CFAP410* | 0.42 | 2.7E-11 | *NA* | *NA* | -0.059 | 0.48 | *NA* | *NA* | *NA* | *NA* | *NA* | *NA* |
| rs10463311 | C | *GPX3/TNIP1* | 0.08 | 2.7E-10 | 0.013 | 0.43 | 0.004 | 0.85 | -0.010 | 0.05 | -0.033 | 0.47 | *NA* | *NA* |
| rs17785991 | A | *SLC9A8/SPATA2* | 0.07 | 3.2E-10 | 0.047 | 0.002 | 0.016 | 0.44 | 0.006 | 0.19 | 0.062 | 0.18 | *NA* | *NA* |
| rs4075094 | A | *TBK1* | -0.10 | 2.1E-09 | -0.019 | 0.40 | 0.051 | 0.16 | 0.010 | 0.12 | -0.004 | 0.95 | *NA* | *NA* |
| rs517339 | C | *ERGIC1/CREBRF* | -0.06 | 5.6E-09 | 0.007 | 0.62 | 0.002 | 0.92 | -0.0004 | 0.93 | -0.069 | 0.09 | *NA* | *NA* |
| rs62333164 | A | *NEK1* | 0.07 | 6.9E-09 | -0.025 | 0.11 | 0.058 | 0.0014 | -0.004 | 0.38 | -0.074 | 0.08 | *NA* | *NA* |
| rs2985994 | C | *COG3* | 0.07 | 1.2E-08 | -0.002 | 0.89 | -0.002 | 0.91 | -0.008 | 0.12 | -0.003 | 0.94 | *NA* | *NA* |
| rs10280711 | G | *PTPRN2* | 0.09 | 1.8E-08 | 0.003 | 0.89 | -0.031 | 0.38 | 0.001 | 0.90 | 0.167 | 0.005 | *NA* | *NA* |
| rs34311866 | C | *GAK* | 0.07 | 1.1E-06 | 0.015 | 0.46 | 0.227 | 8.0E-23 | *NA* | *NA* | 0.093 | 0.13 | *NA* | *NA* |
| rs2632516 | C | *BZRAP1-AS1* | -0.04 | 0.00010 | -0.075 | 3.7E-07 | 0.001 | 0.95 | -0.001 | 0.74 | -0.011 | 0.79 | *NA* | *NA* |

**Supplementary tables 21-24. Downstreamer enrichment analyses.**

**Supplementary table 21. Downstreamer HPO term enrichment.**

| HPO term | Description | Enrichment Z-score | Enrichment P-value |
|---|---|---|---|
| HP:0002120 | Cerebral cortical atrophy | 5.63 | $1.8\times10^{-8}$ |
| HP:0007367 | Atrophy/Degeneration affecting the central nervous system | 5.47 | $4.6\times10^{-8}$ |
| HP:0001288 | Gait disturbance | 5.38 | $7.6\times10^{-8}$ |
| HP:0001311 | Abnormal nervous system electrophysiology | 5.06 | $4.1\times10^{-7}$ |
| HP:0007369 | Atrophy/Degeneration affecting the cerebrum | 5.06 | $4.2\times10^{-7}$ |
| HP:0031466 | Impairment in personality functioning | 5.00 | $5.7\times10^{-7}$ |
| HP:0012444 | Brain atrophy | 4.96 | $6.9\times10^{-7}$ |
| HP:0002059 | Cerebral atrophy | 4.96 | $7.1\times10^{-7}$ |
| HP:0002465 | Poor speech | 4.94 | $8.0\times10\text{-}^{7}$ |
| HP:0003679 | Pace of progression | 4.93 | $8.3\times10^{-7}$ |
| HP:0003693 | Distal amyotrophy | 4.92 | $8.6\times10^{-7}$ |
| HP:0000750 | Delayed speech and language development | 4.80 | $1.6\times10^{-6}$ |
| HP:0030178 | Abnormality of central nervous system electrophysiology | 4.79 | $1.7\times10_{-6}$ |
| HP:0000713 | Agitation | 4.79 | $1.7\times10^{-6}$ |
| HP:0100852 | Abnormal fear/anxiety-related behavior | 4.78 | $1.7\times10^{-6}$ |
| HP:0012757 | Abnormal neuron morphology | 4.76 | $2.0\times10^{-6}$ |
| HP:0002071 | Abnormality of extrapyramidal motor function | 4.74 | $2.1\times10^{-6}$ |
| HP:0100022 | Abnormality of movement | 4.73 | $2.3\times10^{-6}$ |
| HP:0003674 | Onset | 4.73 | $2.3\times10^{-6}$ |
| HP:0002067 | Bradykinesia | 4.72 | $2.4\times10^{-6}$ |
| HP:0002353 | EEG abnormality | 4.72 | $2.4\times10^{-6}$ |
| HP:0025270 | Abnormality of esophagus physiology | 4.7 | $2.6\times10^{-6}$ |
| HP:0000739 | Anxiety | 4.7 | $2.6\times10^{-6}$ |
| HP:0002021 | Pyloric stenosis | 4.68 | $2.9\times10^{-6}$ |
| HP:0003676 | Progressive | 4.68 | $2.9\times10^{-6}$ |
| HP:0002015 | Dysphagia | 4.67 | $3.0\times10^{-6}$ |
| HP:0000711 | Restlessness | 4.63 | $3.7\times10^{-6}$ |
| HP:0000764 | Peripheral axonal degeneration | 4.62 | $3.9\times10^{-6}$ |
| HP:0003593 | Infantile onset | 4.61 | $4.0\times10^{-6}$ |
| HP:0031826 | Abnormal reflex | 4.58 | $4.6\times10^{-6}$ |
| HP:0100704 | Cortical visual impairment | 4.57 | $4.8\times10^{-6}$ |

| | | | |
|---|---|---|---|
| HP:0002450 | Abnormal motor neuron morphology | 4.55 | $5.3\times10^{-6}$ |
| HP:0031797 | Clinical course | 4.52 | $6.1\times10^{-6}$ |
| HP:0011799 | Abnormality of facial soft tissue | 4.52 | $6.2\times10^{-6}$ |
| HP:0002167 | Neurological speech impairment | 4.49 | $7.1\times10^{-6}$ |
| HP:0000716 | Depressivity | 4.48 | $7.6\times10^{-6}$ |
| HP:0002311 | Incoordination | 4.47 | $8.0\times10^{-6}$ |
| HP:0001344 | Absent speech | 4.46 | $8.3\times10^{-6}$ |
| HP:0000194 | Open mouth | 4.45 | $8.7\times10^{-6}$ |
| HP:0001257 | Spasticity | 4.44 | $9.2\times10^{-6}$ |
| HP:0001276 | Hypertonia | 4.43 | $9.4\times10^{-6}$ |
| HP:0004305 | Involuntary movements | 4.43 | $9.5\times10^{-6}$ |
| HP:0002540 | Inability to walk | 4.42 | $9.7\times10^{-6}$ |
| HP:0008207 | Primary adrenal insufficiency | 4.42 | $9.8\times10^{-6}$ |
| HP:0000301 | Abnormality of facial musculature | 4.41 | $1.0\times10^{-5}$ |
| HP:0007373 | Motor neuron atrophy | 4.40 | $1.1\times10^{-5}$ |
| HP:0000738 | Hallucinations | 4.39 | $1.2\times10^{-5}$ |
| HP:0010993 | Abnormality of the cerebral subcortex | 4.37 | $1.2\times10^{-5}$ |
| HP:0031828 | Abnormal superficial reflex | 4.37 | $1.2\times10^{-5}$ |
| HP:0003487 | Babinski sign | 4.37 | $1.2\times10^{-5}$ |
| HP:0004400 | Abnormality of the pylorus | 4.34 | $1.4\times10^{-5}$ |
| HP:0001336 | Myoclonus | 4.33 | $1.5\times10^{-5}$ |
| HP:0002072 | Chorea | 4.32 | $1.6\times10^{-5}$ |
| HP:0000708 | Behavioral abnormality | 4.31 | $1.6\times10^{-5}$ |
| HP:0001761 | Pes cavus | 4.30 | $1.7\times10^{-5}$ |
| HP:0001272 | Cerebellar atrophy | 4.30 | $1.7\times10^{-5}$ |
| HP:0003477 | Peripheral axonal neuropathy | 4.28 | $1.9\times10^{-0}$ |
| HP:0002577 | Abnormality of the stomach | 4.27 | $2.0\times10^{-5}$ |

**Supplementary table 22. Downstreamer Reactome enrichment.**

| Annotation | Parent branch | Enrichment Z-score | Enrichment P-value | Enrichment Q-value |
|---|---|---|---|---|
| Membrane Trafficking | Vesicle mediated transport | 4.60 | $4.2\times10^{-6}$ | 0.0020 |
| Intra-Golgi and retrograde Golgi-to-ER traffic | Vesicle mediated transport | 4.34 | $1.4\times10^{-5}$ | 0.0022 |
| Macroautophagy | Autophagy | 4.16 | $3.2\times10^{-5}$ | 0.0022 |

**Supplementary table 23. Downstreamer GO Biological Processes enrichment.**

| Annotation | Enrichment Z-score | Enrichment P-value | Enrichment Q-value |
|---|---|---|---|
| exocytosis | 5.12 | $3.1\times10^{-7}$ | 0.0000 |
| cytoplasmic microtubule organization | 4.96 | $7.0\times10^{-7}$ | 0.0000 |
| regulation of phosphoprotein phosphatase activity | 4.66 | $3.2\times10^{-6}$ | 0.0000 |
| protein targeting to vacuole | 4.59 | $4.5\times10^{-6}$ | 0.0000 |
| endosomal transport | 4.54 | $5.5\times10^{-6}$ | 0.0000 |
| protein K48-linked deubiquitination | 4.50 | $6.8\times10^{-6}$ | 0.0000 |
| microtubule nucleation | 4.42 | $1.0\times10^{-5}$ | $7.7\times10^{-4}$ |
| ubiquitin-dependent protein catabolic process | 4.38 | $1.2\times10^{-5}$ | $7.7\times10^{-4}$ |
| intracellular protein transport | 4.33 | $1.5\times10^{-5}$ | $7.7\times10^{-4}$ |
| retrograde transport, endosome to Golgi | 4.31 | $1.6\times10^{-5}$ | $7.7\times10^{-4}$ |
| clathrin-dependent endocytosis | 4.25 | $2.1\times10^{-5}$ | $7.7\times10^{-4}$ |
| vesicle-mediated transport | 4.19 | $2.8\times10^{-5}$ | $7.7\times10^{-4}$ |
| protein localization to phagophore assembly site | 4.13 | $3.6\times10^{-5}$ | $9.7\times10^{-4}$ |
| macroautophagy | 4.12 | $3.8\times10^{-5}$ | $9.7\times10^{-4}$ |

**Supplementary table 24. Downstreamer GO Cellular Components enrichment.**

| Annotation | Enrichment Z-score | Enrichment P-value | Enrichment Q-value |
|---|---|---|---|
| microtubule associated complex | 4.14 | $3.5\times10^{-5}$ | 0.003 |
| phagophore assembly site | 3.99 | $6.6\times10^{-5}$ | 0.006 |

**Supplementary table 25-33. Mendelian randomization details.**

**Supplementary table 25. Included GWAS for instrument selection.**
[Table in Excel file]

**Supplementary table 26. Full MR analyses.** Life-style traits.

| | | Instrument cut-off: $P < 5\times10^{-8}$ | | | | Instrument cut-off: $P < 5\times10^{-5}$ | | | |
|---|---|---|---|---|---|---|---|---|---|
| Exposure | Method | SNPs | beta | se | P | SNPs | beta | se | P |
| Age Of Smoking Initiation | Weighted median | 6 | 0.15 | 0.35 | 0.67 | 103 | -0.2 | 0.13 | 0.12 |
| | Weighted mode | 6 | 0.25 | 0.53 | 0.66 | 103 | -0.37 | 0.34 | 0.27 |
| | Simple mode | 6 | 0.27 | 0.58 | 0.66 | 103 | -0.4 | 0.38 | 0.29 |
| | MR Egger | 6 | 0.92 | 1.81 | 0.64 | 103 | -0.41 | 0.38 | 0.29 |
| | IVW | 6 | 0.11 | 0.38 | 0.78 | 103 | -0.07 | 0.1 | 0.50 |
| Cigarettes per Day | Weighted median | 22 | -0.07 | 0.06 | 0.28 | 140 | -0.07 | 0.06 | 0.21 |
| | Weighted mode | 22 | -0.06 | 0.06 | 0.28 | 140 | -0.05 | 0.06 | 0.41 |
| | Simple mode | 22 | -0.02 | 0.11 | 0.83 | 140 | 0.02 | 0.12 | 0.84 |
| | MR Egger | 22 | -0.06 | 0.09 | 0.52 | 140 | -0.06 | 0.07 | 0.36 |
| | IVW | 22 | -0.05 | 0.05 | 0.33 | 140 | -0.03 | 0.03 | 0.44 |
| Alcoholic drinks per week | Weighted median | 35 | -0.18 | 0.15 | 0.24 | 252 | -0.15 | 0.15 | 0.3 |
| | Weighted mode | 35 | -0.23 | 0.15 | 0.14 | 252 | -0.21 | 0.13 | 0.12 |
| | Simple mode | 35 | -0.45 | 0.36 | 0.22 | 252 | -0.21 | 0.38 | 0.59 |
| | MR Egger | 35 | -0.2 | 0.24 | 0.40 | 252 | -0.12 | 0.17 | 0.47 |
| | IVW | 35 | 0 | 0.15 | 1 | 252 | 0.04 | 0.08 | 0.61 |
| Body Mass Index | Weighted median | 506 | -0.06 | 0.07 | 0.35 | 526 | -0.01 | 0.07 | 0.88 |
| | Weighted mode | 506 | -0.04 | 0.12 | 0.74 | 526 | 0.01 | 0.15 | 0.96 |
| | Simple mode | 506 | 0.35 | 0.19 | 0.07 | 526 | 0.07 | 0.22 | 0.76 |
| | MR Egger | 506 | -0.12 | 0.11 | 0.26 | 526 | 0.01 | 0.12 | 0.91 |
| | IVW | 506 | -0.03 | 0.04 | 0.53 | 526 | -0.02 | 0.05 | 0.61 |
| Moderate activity | Weighted median | 8 | -0.11 | 0.17 | 0.50 | 96 | 0.03 | 0.07 | 0.71 |
| | Weighted mode | 8 | -0.17 | 0.23 | 0.48 | 96 | -0.18 | 0.18 | 0.32 |
| | Simple mode | 8 | -0.17 | 0.23 | 0.50 | 96 | -0.18 | 0.2 | 0.37 |
| | MR Egger | 8 | 0.08 | 0.89 | 0.93 | 96 | 0.16 | 0.23 | 0.48 |
| | IVW | 8 | 0.02 | 0.12 | 0.89 | 96 | 0.03 | 0.06 | 0.54 |
| Vigorous activity | Weighted median | 8 | 0.31 | 0.21 | 0.14 | 79 | 0.16 | 0.09 | 0.09 |
| | Weighted mode | 8 | 0.53 | 0.34 | 0.16 | 79 | 0.24 | 0.22 | 0.28 |
| | Simple mode | 8 | 0.59 | 0.34 | 0.12 | 79 | 0.24 | 0.25 | 0.33 |
| | MR Egger | 8 | -1.64 | 2.4 | 0.52 | 79 | -0.27 | 0.25 | 0.29 |
| | IVW | 8 | 0.24 | 0.18 | 0.19 | 79 | 0.07 | 0.07 | 0.31 |

**Supplementary table 26. Full MR analyses (cont.)** Cardiovascular traits and Educational attainment.

| | | Instrument cut-off: $P < 5×10^{-8}$ | | | | Instrument cut-off: $P < 5×10^{-5}$ | | | |
|---|---|---|---|---|---|---|---|---|---|
| Exposure | Method | SNPs | beta | se | P | SNPs | beta | se | P |
| Diastolic blood pressure | Weighted median | 448 | 0 | 0.01 | 0.48 | 463 | 0 | 0.01 | 0.86 |
| | Weighted mode | 448 | 0.01 | 0.01 | 0.53 | 463 | 0 | 0.01 | 0.99 |
| | Simple mode | 448 | 0 | 0.02 | 0.82 | 463 | 0 | 0.02 | 0.82 |
| | MR Egger | 448 | 0.01 | 0.01 | 0.22 | 463 | 0.01 | 0.01 | 0.36 |
| | IVW | 448 | 0 | 0 | 0.58 | 463 | 0 | 0 | 0.49 |
| Systolic blood pressure | Weighted median | 450 | 0.01 | 0.003 | 0.06 | 514 | 0.01 | 0.004 | 0.06 |
| | Weighted mode | 450 | 0.01 | 0.01 | 0.3 | 514 | 0.01 | 0.01 | 0.32 |
| | Simple mode | 450 | 0.01 | 0.01 | 0.54 | 514 | 0.01 | 0.01 | 0.6 |
| | MR Egger | 450 | 0.01 | 0.01 | 0.09 | 514 | 0.01 | 0.01 | 0.07 |
| | IVW | 450 | 0.01 | 0.002 | 0.06 | 514 | 0.01 | 0.002 | 0.003 |
| Coronary artery disease | Weighted median | 144 | 0.03 | 0.03 | 0.44 | 388 | 0.02 | 0.03 | 0.45 |
| | Weighted mode | 144 | 0.04 | 0.04 | 0.35 | 388 | 0.03 | 0.04 | 0.43 |
| | Simple mode | 144 | -0.03 | 0.08 | 0.74 | 388 | -0.03 | 0.08 | 0.69 |
| | MR Egger | 144 | 0.08 | 0.05 | 0.09 | 388 | 0.04 | 0.04 | 0.34 |
| | IVW | 144 | 0.03 | 0.02 | 0.15 | 388 | 0.03 | 0.02 | 0.11 |
| Stroke | Weighted median | 8 | 0.19 | 0.1 | 0.06 | 42 | 0.07 | 0.06 | 0.23 |
| | Weighted mode | 8 | 0.22 | 0.12 | 0.1 | 42 | -0.03 | 0.13 | 0.81 |
| | Simple mode | 8 | 0.22 | 0.14 | 0.16 | 42 | -0.03 | 0.14 | 0.85 |
| | MR Egger | 8 | 0.84 | 0.63 | 0.23 | 42 | 0.01 | 0.13 | 0.97 |
| | IVW | 8 | 0.08 | 0.1 | 0.44 | 42 | 0.07 | 0.04 | 0.11 |
| Years of schooling | Weighted median | 306 | -0.18 | 0.09 | 0.04 | 681 | -0.16 | 0.07 | 0.02 |
| | Weighted mode | 306 | -0.15 | 0.26 | 0.56 | 681 | -0.2 | 0.24 | 0.41 |
| | Simple mode | 306 | -0.19 | 0.28 | 0.51 | 681 | -0.22 | 0.28 | 0.43 |
| | MR Egger | 306 | 0.05 | 0.27 | 0.85 | 681 | 0.03 | 0.17 | 0.85 |
| | IVW | 306 | -0.18 | 0.07 | 0.01 | 681 | -0.19 | 0.05 | 0.0002 |

**Supplementary table 26. Full MR analyses (cont.)** Lipids.

| | | Instrument cut-off: $P < 5\times10^{-8}$ | | | | Instrument cut-off: $P < 5\times10^{-5}$ | | | |
|---|---|---|---|---|---|---|---|---|---|
| Exposure | Method | SNPs | beta | se | P | SNPs | beta | se | P |
| Triglycerides | Weighted median | 54 | 0.01 | 0.06 | 0.8 | 135 | 0.01 | 0.06 | 0.81 |
| | Weighted mode | 54 | 0.05 | 0.05 | 0.4 | 135 | 0.06 | 0.05 | 0.27 |
| | Simple mode | 54 | 0.18 | 0.11 | 0.1 | 135 | 0.13 | 0.1 | 0.2 |
| | MR Egger | 54 | 0.1 | 0.07 | 0.19 | 135 | 0.07 | 0.07 | 0.35 |
| | IVW | 54 | 0.03 | 0.04 | 0.43 | 135 | 0.03 | 0.04 | 0.53 |
| Total cholesterol | Weighted median | 83 | 0.15 | 0.04 | 0.0003 | 178 | 0.15 | 0.04 | 0.0007 |
| | Weighted mode | 83 | 0.13 | 0.04 | 0.004 | 178 | 0.11 | 0.04 | 0.007 |
| | Simple mode | 83 | 0.02 | 0.1 | 0.86 | 178 | 0.03 | 0.11 | 0.75 |
| | MR Egger | 83 | 0.13 | 0.05 | 0.01 | 178 | 0.15 | 0.05 | 0.005 |
| | IVW | 83 | 0.09 | 0.03 | 0.005 | 178 | 0.08 | 0.03 | 0.02 |
| HDL cholesterol | Weighted median | 87 | 0.01 | 0.05 | 0.86 | 158 | 0 | 0.05 | 0.93 |
| | Weighted mode | 87 | 0.01 | 0.05 | 0.9 | 158 | -0.02 | 0.05 | 0.73 |
| | Simple mode | 87 | 0.01 | 0.09 | 0.94 | 158 | 0.03 | 0.1 | 0.79 |
| | MR Egger | 87 | -0.06 | 0.07 | 0.34 | 158 | -0.08 | 0.07 | 0.27 |
| | IVW | 87 | 0.04 | 0.04 | 0.32 | 158 | 0.03 | 0.04 | 0.49 |
| LDL cholesterol | Weighted median | 75 | 0.08 | 0.04 | 0.06 | 147 | 0.08 | 0.05 | 0.07 |
| | Weighted mode | 75 | 0.06 | 0.04 | 0.16 | 147 | 0.07 | 0.04 | 0.04 |
| | Simple mode | 75 | 0.2 | 0.09 | 0.03 | 147 | 0.18 | 0.09 | 0.05 |
| | MR Egger | 75 | 0.11 | 0.05 | 0.03 | 147 | 0.1 | 0.05 | 0.03 |
| | IVW | 75 | 0.07 | 0.03 | 0.03 | 147 | 0.06 | 0.03 | 0.08 |

**Supplementary table 26. Full MR analyses (cont.)** Inflammation markers.

| | | Instrument cut-off: $P < 5\times10^{-8}$ | | | | Instrument cut-off: $P < 5\times10^{-5}$ | | | |
|---|---|---|---|---|---|---|---|---|---|
| Exposure | Method | SNPs | beta | se | P | SNPs | beta | se | P |
| C-Reactive protein level | Weighted median | 55 | -0.06 | 0.05 | 0.25 | 62 | -0.08 | 0.06 | 0.19 |
| | Weighted mode | 55 | -0.04 | 0.04 | 0.27 | 62 | -0.08 | 0.06 | 0.17 |
| | Simple mode | 55 | -0.02 | 0.09 | 0.83 | 62 | 0.16 | 0.2 | 0.43 |
| | MR Egger | 55 | -0.05 | 0.05 | 0.28 | 62 | -0.12 | 0.08 | 0.13 |
| | IVW | 55 | -0.01 | 0.03 | 0.83 | 62 | -0.06 | 0.06 | 0.27 |
| white blood cell count | Weighted median | 480 | -0.06 | 0.05 | 0.23 | 744 | -0.05 | 0.04 | 0.23 |
| | Weighted mode | 480 | -0.02 | 0.07 | 0.76 | 744 | -0.02 | 0.07 | 0.78 |
| | Simple mode | 480 | -0.04 | 0.12 | 0.76 | 744 | -0.04 | 0.12 | 0.75 |
| | MR Egger | 480 | -0.13 | 0.06 | 0.04 | 744 | -0.08 | 0.06 | 0.15 |
| | IVW | 480 | -0.05 | 0.03 | 0.06 | 744 | -0.05 | 0.03 | 0.07 |
| lymphocyte cell count | Weighted median | 486 | -0.05 | 0.05 | 0.27 | 739 | -0.09 | 0.04 | 0.04 |
| | Weighted mode | 486 | -0.13 | 0.09 | 0.16 | 739 | -0.19 | 0.08 | 0.01 |
| | Simple mode | 486 | 0.03 | 0.14 | 0.84 | 739 | -0.01 | 0.13 | 0.92 |
| | MR Egger | 486 | -0.05 | 0.06 | 0.39 | 739 | -0.04 | 0.05 | 0.41 |
| | IVW | 486 | -0.03 | 0.03 | 0.36 | 739 | -0.03 | 0.03 | 0.26 |
| neutrophil cell count | Weighted median | 404 | -0.05 | 0.05 | 0.3 | 656 | -0.05 | 0.05 | 0.28 |
| | Weighted mode | 404 | -0.03 | 0.06 | 0.65 | 656 | -0.03 | 0.07 | 0.61 |
| | Simple mode | 404 | -0.01 | 0.11 | 0.92 | 656 | -0.01 | 0.12 | 0.9 |
| | MR Egger | 404 | -0.13 | 0.07 | 0.06 | 656 | -0.09 | 0.06 | 0.13 |
| | IVW | 404 | -0.03 | 0.03 | 0.32 | 656 | -0.03 | 0.03 | 0.27 |
| monocyte cell count | Weighted median | 472 | 0.06 | 0.04 | 0.16 | 716 | 0.05 | 0.04 | 0.17 |
| | Weighted mode | 472 | 0.05 | 0.05 | 0.25 | 716 | 0.04 | 0.04 | 0.41 |
| | Simple mode | 472 | 0 | 0.09 | 0.96 | 716 | -0.03 | 0.1 | 0.78 |
| | MR Egger | 472 | 0.05 | 0.04 | 0.32 | 716 | 0.05 | 0.04 | 0.24 |
| | IVW | 472 | -0.01 | 0.03 | 0.82 | 716 | -0.01 | 0.02 | 0.76 |
| eosinophil cell count | Weighted median | 422 | -0.02 | 0.04 | 0.62 | 645 | -0.03 | 0.04 | 0.53 |
| | Weighted mode | 422 | -0.04 | 0.06 | 0.48 | 645 | -0.04 | 0.05 | 0.5 |
| | Simple mode | 422 | -0.16 | 0.1 | 0.13 | 645 | -0.06 | 0.1 | 0.6 |
| | MR Egger | 422 | -0.03 | 0.05 | 0.58 | 645 | -0.03 | 0.05 | 0.54 |
| | IVW | 422 | -0.03 | 0.03 | 0.27 | 645 | -0.03 | 0.03 | 0.28 |

**Supplementary table 27. MR results after outlier removal for total cholesterol.**

| | | Instrument cut-off: $P < 5\times10^{-8}$ | | | | Instrument cut-off: $P < 5\times10^{-5}$ | | | |
|---|---|---|---|---|---|---|---|---|---|
| Method | Outliers removed | SNPs | beta | se | P | SNPs | beta | se | P |
| Weighted median | none | 83 | 0.154 | 0.044 | 0.0004 | 178 | 0.149 | 0.044 | 0.0008 |
| Weighted mode | none | 83 | 0.132 | 0.041 | 0.002 | 178 | 0.109 | 0.04 | 0.008 |
| Simple mode | none | 83 | 0.017 | 0.091 | 0.85 | 178 | 0.034 | 0.105 | 0.75 |
| MR Egger | none | 83 | 0.132 | 0.053 | 0.01 | 178 | 0.147 | 0.052 | 0.005 |
| IVW | none | 83 | 0.091 | 0.032 | 0.005 | 178 | 0.077 | 0.033 | 0.02 |
| Weighted median | IVW | 75 | 0.155 | 0.042 | 0.00023 | 156 | 0.148 | 0.043 | 0.0006 |
| Simple mode | IVW | 75 | -0.016 | 0.091 | 0.86 | 156 | 0.032 | 0.101 | 0.75 |
| Weighted mode | IVW | 75 | 0.161 | 0.042 | 0.0003 | 156 | 0.106 | 0.043 | 0.01 |
| MR Egger | IVW | 75 | 0.14 | 0.046 | 0.003 | 156 | 0.157 | 0.04 | 0.0001 |
| IVW | IVW | 75 | 0.092 | 0.029 | 0.001 | 156 | 0.071 | 0.026 | 0.007 |
| Weighted median | Egger | 76 | 0.166 | 0.044 | 0.0002 | 157 | 0.157 | 0.043 | 0.0003 |
| Weighted mode | Egger | 76 | 0.166 | 0.043 | 0.0003 | 157 | 0.145 | 0.043 | 0.001 |
| Simple mode | Egger | 76 | -0.002 | 0.091 | 0.98 | 157 | 0.047 | 0.1 | 0.64 |
| MR Egger | Egger | 76 | 0.184 | 0.049 | 0.0004 | 157 | 0.2 | 0.042 | $5.5\times10^{-6}$ |
| IVW | Egger | 76 | 0.098 | 0.03 | 0.001 | 157 | 0.078 | 0.028 | 0.005 |

**Supplementary table 28. MR results for reverse causality of genetic instrument for ALS on total cholesterol.**

| | | | Instrument cut-off: $P < 5\times10^{-8}$ | | | | Instrument cut-off: $P < 5\times10^{-5}$ | | | |
|---|---|---|---|---|---|---|---|---|---|---|
| Exposure | Outcome | Method | SNPs | beta | se | P | SNPs | beta | se | P |
| ALS | Total cholesterol | Weighted median | 9 | 0.02 | 0.03 | 0.4 | 75 | 0.02 | 0.02 | 0.2 |
| ALS | Total cholesterol | Weighted mode | 9 | 0.02 | 0.03 | 0.4 | 75 | 0.02 | 0.02 | 0.4 |
| ALS | Total cholesterol | Simple mode | 9 | 0.02 | 0.04 | 0.7 | 75 | 0.01 | 0.04 | 0.7 |
| ALS | Total cholesterol | MR Egger | 9 | 0.01 | 0.07 | 0.9 | 75 | -0.03 | 0.04 | 0.5 |
| ALS | Total cholesterol | IVW | 9 | 0.03 | 0.02 | 0.2 | 75 | 0.01 | 0.01 | 0.5 |

**Supplementary table 29. Multivariable MR results for total cholesterol, conditional on years of schooling.**

| | Instrument cut-off: $P < 5\times10^{-8}$ | | | Instrument cut-off: $P < 5\times10^{-5}$ | | |
|---|---|---|---|---|---|---|
| Method | beta | se | P | beta | se | P |
| IVW | 0.09 | 0.03 | 0.005 | 0.08 | 0.03 | 0.02 |
| Multivariable MR | 0.15 | 0.04 | $2\times10^{-4}$ | 0.11 | 0.04 | 0.005 |

**Supplementary table 30. MR instrument coverage**
[table in Excel file]

**Supplementary table 31. MR F-statistics**
[table in Excel file]

**Supplementary table 32. Wald ratios**
[table in Excel file]

**Supplementary table 33. Heterogeneity statistics**
[table in Excel file]
